# Shared multicellular injury programs of acute and chronic kidney disease enable mechanistic patient stratification

**DOI:** 10.64898/2026.03.05.26347522

**Authors:** Robin Fallegger, Sergio A. Gomez-Ochoa, Charlotte Boys, Ricardo Omar Ramirez Flores, Jovan Tanevski, Evanthia Pashos, Denis Feliers, Mary Piper, Jennifer A. Schaub, Zixiang Zhou, Weiguang Mao, Xi Chen, Rachel S. G. Sealfon, Rajasree Menon, Viji Nair, Sean Eddy, Fadhl M Alakwaa, Laura Pyle, Ye Ji Choi, Petter Bjornstad, Charles E. Alpers, Markus Bitzer, Andrew S. Bomback, M. Luiza Caramori, Dawit Demeke, Agnes B. Fogo, Leal C. Herlitz, Krzysztof Kiryluk, James P. Lash, Raghavan Murugan, John F. O’Toole, Paul M. Palevsky, Chirag R. Parikh, Sylvia E. Rosas, Avi Z Rosenberg, John R. Sedor, Miguel A. Vazquez, Sushrut S. Waikar, F. Perry Wilson, Jeffrey B. Hodgin, Laura Barisoni, Jonathan Himmelfarb, Sanjay Jain, Wenjun Ju, Olga G. Troyanskaya, Matthias Kretzler, Michael T. Eadon, Julio Saez-Rodriguez

## Abstract

**Abstract:** Acute kidney injury (AKI) and chronic kidney disease (CKD) are two interconnected clinical conditions, both defined by degree of functional impairment, but with heterogeneous clinical trajectories. Using new transcriptomic technologies, recent studies have described the cellular diversity in the healthy and injured kidney at the single cell level. Here, we used single nucleus transcriptomics to investigate the molecular diversity and commonalities in kidney biopsies from over 150 participants with AKI and CKD enrolled within the Kidney Precision Medicine Project (KPMP) and did so at the patient participant level. Using an unsupervised approach, we identified two multi-cellular programs associated with clinical and histopathological features of acute injury and chronic damage, respectively. We found that these programs are expressed across patients with AKI and CKD, supporting shared, rather than distinct, underlying molecular mechanisms. These programs capture tissue-level compositional changes towards adaptive and failed-repair states in tubular epithelial cells, as well as intra-cellular molecular changes characteristic of stress in all cell types. We identified subunits of the NFkB and AP-1 complexes, as well as members of the STAT family, as putative upstream regulators of the acute and chronic programs. We were able to map these continuous molecular measures of acute injury and chronic damage to urine and plasma protein profiles obtained at time of biopsy. These non-invasive protein signatures were predictive of renal outcomes in an independent cohort of 44 thousand participants from the UK biobank. In summary, unbiased identification of cellular programs in kidney disease biopsies defined molecular programs of injury cutting across conventional disease categorization and established a non-invasive molecular link to long term patient outcomes.

**Graphical Abstract:** 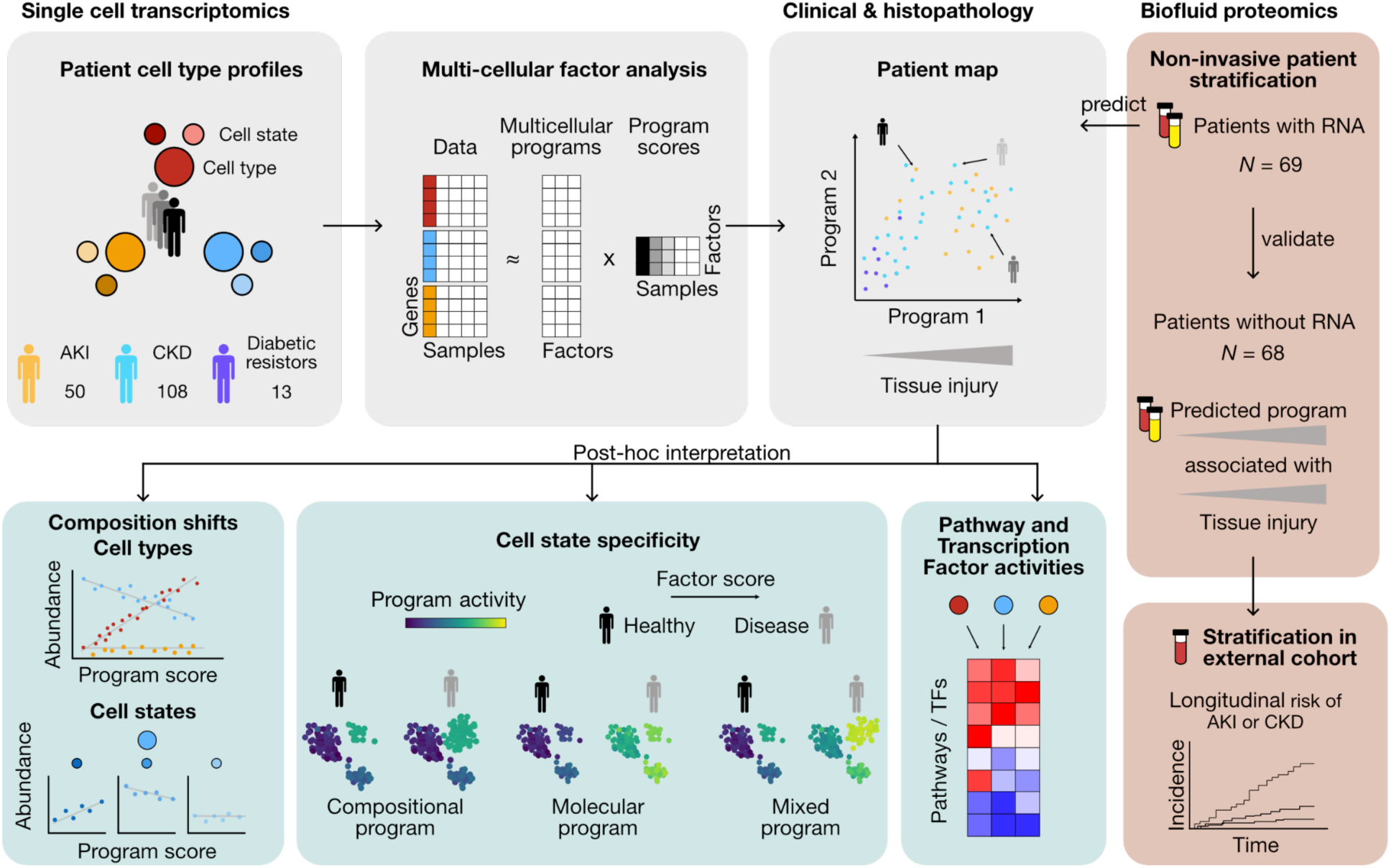

## Introduction

Acute kidney injury (AKI) and chronic kidney disease (CKD) are conditions with a high prevalence, severe impact on quality of life and increased mortality^1–4^. It is estimated that CKD affects more than 10% of the global adult population, whereas AKI is present in up to half of intensive care patients^3,5^. AKI and CKD are currently considered distinct clinical entities, but are interconnected: episodes of AKI confer a substantially higher risk of developing or accelerating CKD, while pre-existing CKD predisposes to AKI^6–8^. Despite this recognition, clinical trajectories within each condition remain highly heterogeneous and the reasons why some recover or remain stable, while others progress remain poorly understood, limiting effective clinical management and therapeutic development^9–11^.

A central challenge is that both AKI and CKD are defined by functional metrics, such as changes in serum creatinine, that capture the consequence of injury rather than its cause or cellular basis, and histopathological scoring requires tissue biopsy and then often identifies structural changes late and incompletely^12–14^. Consequently, patients with similar clinical profiles can harbor markedly different molecular landscapes^15,16^. Non-invasive biomarkers such as KIM-1 and NGAL have been associated with outcomes based on clinical categorisation^17^, but a framework linking biomarkers to tissue level molecular changes of injury and repair is currently missing. There is therefore a need for unbiased molecular characterization of patient heterogeneity that bridges cellular mechanisms and clinical diversity. New molecular tissue level profiling technologies, including single-cell resolution RNA sequencing, chromatin accessibility assays, and spatial sequencing technologies, have allowed unprecedented insight into the cellular diversity of both the healthy and diseased kidney, opening the door to molecular stratification of kidney health^18^. Recent atlas efforts have mapped the cellular diversity in the human and mouse kidney and identified mechanisms driving cell state changes in disease, including key regulators orchestrating the transition from injured (so-called adaptive) tubular epithelial cells back to healthy states or forward into failed repair^19–26^. However, these studies have generally relied on clinical definitions of disease, supervised quantification of prognostic clinical-pathologic metrics, or mouse disease models, none of which are designed to capture the intertwined nature of AKI and CKD. Consequently, it remains unclear how the identified cell states explain patient heterogeneity both in clinical trajectories and outcomes across these conditions.

In this study, we used unsupervised analysis of single-nucleus transcriptomic data to explore *patient* diversity and assess how the molecular profiles can inform our understanding of individual patient disease states. For this purpose, we leveraged data from 132 living study participants enrolled in the Kidney Precision Medicine Project (KPMP) cohort recently released in the Human Kidney Atlas v2^23^. These participants volunteered research kidney biopsies, which in the setting of AKI or CKD are usually not indicated for clinical care, and provide a representative cross-section of these kidney diseases as seen in clinical practice^27^. Extensive multi-center clinical and pathologic data from the individual participants was available for our analysis.

We identified two programs describing patient heterogeneity solely based on single cell molecular profiles. The two programs were categorized in terms of acute and chronic cellular injury, and individuals with AKI and CKD displayed components of each. We found that these multicellular programs capture increases in both proportions of cell states previously identified and general (cell-state agnostic) intracellular gene expression changes in epithelial cells in an injury-associated fashion. We were able to link these transcriptomically-defined spectra of acute and chronic injury to the plasma and urine proteome obtained at time of biopsy, establishing non-invasive surrogates of the intra-renal cellular programs. The non-invasive surrogates of the acute and chronic intrarenal cell states were shown to be associated with long-term clinical outcomes in an independent, population level study (UK biobank).

## Results

### Clinical and histopathologic features of an AKI and CKD cohort

To understand inter-individual variability in CKD and AKI, we used the single-nuclei (snRNA) and single cell RNA (scRNA) data from the integrated Human Kidney Atlas v2 of kidney samples obtained through percutaneous biopsy^23^. Analyses were restricted to samples obtained through percutaneous biopsy to avoid ischemic artefacts (Figure S1, Figure S2), which were previously characterized in this cohort^28^. We analyzed snRNA and scRNA separately as they capture different mRNA-populations. After sample-level quality control filtering for cell number and representation of major kidney cell types, the dataset consisted of 101 distinct study participants in the snRNA atlas, and 67 individuals in the scRNA atlas. 37 study participants were profiled in both modalities, with tissue coming from two separate cores obtained during the same biopsy procedure^21,29^.

Participants were enrolled in the KPMP study as part of three enrollment categories: a CKD group, defined clinically as diabetic kidney disease (DKD) or hypertension-associated CKD (HCKD); acute kidney injury (AKI); and diabetes mellitus-resistors (DM-R). The latter group comprises individuals with long history of type 1 diabetes (≥ 25 years duration) without clinical CKD in their clinical presentation, based on an estimated glomerular filtration rate (eGFR) > 60 mL/min per 1.73 m2 and absence of albuminuria at time of biopsy^27^. Diabetes mellitus-resistors exhibited the highest eGFR, and lowest levels of albuminuria, interstitial fibrosis and tubular atrophy and, as a group, can be considered as having lowest disease burden (Table 1, Table S1).

**Table 1:**
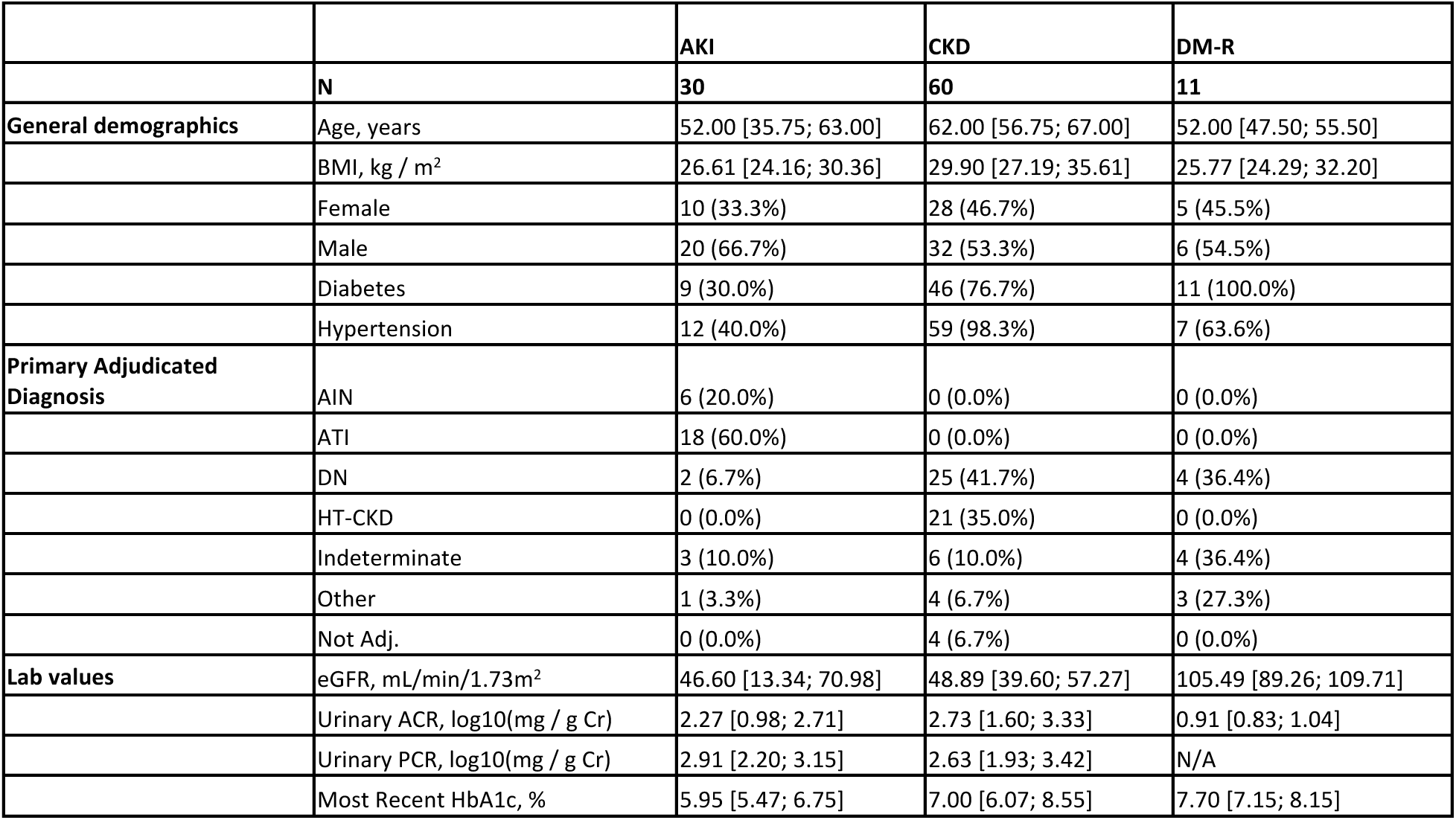
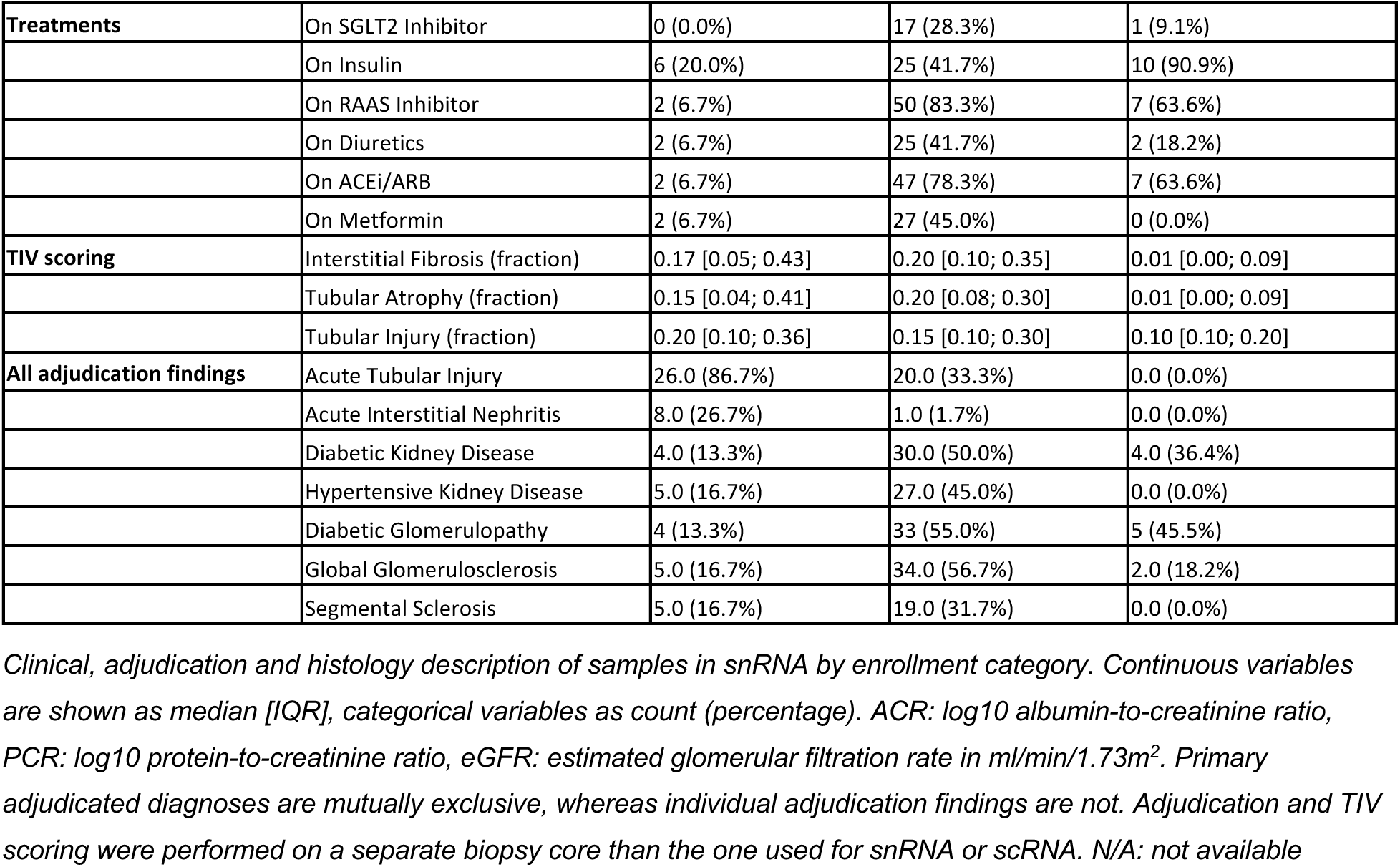
snRNA cohort characteristics.

|  |  | AKI | CKD | DM-R |
| --- | --- | --- | --- | --- |
|  | N | 30 | 60 | 11 |
| General demographics | Age, years | 52.00 [35.75; 63.00] | 62.00 [56.75; 67.00] | 52.00 [47.50; 55.50] |
|  | BMI, kg / m<sup>2</sup> | 26.61 [24.16; 30.36] | 29.90 [27.19; 35.61] | 25.77 [24.29; 32.20] |
|  | Female | 10 (33.3%) | 28 (46.7%) | 5 (45.5%) |
|  | Male | 20 (66.7%) | 32 (53.3%) | 6 (54.5%) |
|  | Diabetes | 9 (30.0%) | 46 (76.7%) | 11 (100.0%) |
|  | Hypertension | 12 (40.0%) | 59 (98.3%) | 7 (63.6%) |
| Primary Adjudicated Diagnosis | AIN | 6 (20.0%) | 0 (0.0%) | 0 (0.0%) |
|  | ATI | 18 (60.0%) | 0 (0.0%) | 0 (0.0%) |
|  | DN | 2 (6.7%) | 25 (41.7%) | 4 (36.4%) |
|  | HT-CKD | 0 (0.0%) | 21 (35.0%) | 0 (0.0%) |
|  | Indeterminate | 3 (10.0%) | 6 (10.0%) | 4 (36.4%) |
|  | Other | 1 (3.3%) | 4 (6.7%) | 3 (27.3%) |
|  | Not Adj. | 0 (0.0%) | 4 (6.7%) | 0 (0.0%) |
| Lab values | eGFR, mL/min/1.73m<sup>2</sup> | 46.60 [13.34; 70.98] | 48.89 [39.60; 57.27] | 105.49 [89.26; 109.71] |
|  | Urinary ACR, log10(mg / g Cr) | 2.27 [0.98; 2.71] | 2.73 [1.60; 3.33] | 0.91 [0.83; 1.04] |
|  | Urinary PCR, log10(mg / g Cr) | 2.91 [2.20; 3.15] | 2.63 [1.93; 3.42] | N/A |
|  | Most Recent HbA1c, % | 5.95 [5.47; 6.75] | 7.00 [6.07; 8.55] | 7.70 [7.15; 8.15] |
| **Treatments** | On SGLT2 Inhibitor | 0 (0.0%) | 17 (28.3%) | 1 (9.1%) |
|  | On Insulin | 6 (20.0%) | 25 (41.7%) | 10 (90.9%) |
|  | On RAAS Inhibitor | 2 (6.7%) | 50 (83.3%) | 7 (63.6%) |
|  | On Diuretics | 2 (6.7%) | 25 (41.7%) | 2 (18.2%) |
|  | On ACEi/ARB | 2 (6.7%) | 47 (78.3%) | 7 (63.6%) |
|  | On Metformin | 2 (6.7%) | 27 (45.0%) | 0 (0.0%) |
| **TIV scoring** | Interstitial Fibrosis (fraction) | 0.17 [0.05; 0.43] | 0.20 [0.10; 0.35] | 0.01 [0.00; 0.09] |
|  | Tubular Atrophy (fraction) | 0.15 [0.04; 0.41] | 0.20 [0.08; 0.30] | 0.01 [0.00; 0.09] |
|  | Tubular Injury (fraction) | 0.20 [0.10; 0.36] | 0.15 [0.10; 0.30] | 0.10 [0.10; 0.20] |
| **All adjudication findings** | Acute Tubular Injury | 26.0 (86.7%) | 20.0 (33.3%) | 0.0 (0.0%) |
|  | Acute Interstitial Nephritis | 8.0 (26.7%) | 1.0 (1.7%) | 0.0 (0.0%) |
|  | Diabetic Kidney Disease | 4.0 (13.3%) | 30.0 (50.0%) | 4.0 (36.4%) |
|  | Hypertensive Kidney Disease | 5.0 (16.7%) | 27.0 (45.0%) | 0.0 (0.0%) |
|  | Diabetic Glomerulopathy | 4 (13.3%) | 33 (55.0%) | 5 (45.5%) |
|  | Global Glomerulosclerosis | 5.0 (16.7%) | 34.0 (56.7%) | 2.0 (18.2%) |
|  | Segmental Sclerosis | 5.0 (16.7%) | 19.0 (31.7%) | 0.0 (0.0%) |

*Clinical, adjudication and histology description of samples in snRNA by enrollment category. Continuous variables are shown as median [IQR], categorical variables as count (percentage). ACR: log10 albumin-to-creatinine ratio, PCR: log10 protein-to-creatinine ratio, eGFR: estimated glomerular filtration rate in ml/min/1.73m<sup>2</sup>. Primary adjudicated diagnoses are mutually exclusive, whereas individual adjudication findings are not. Adjudication and TIV scoring were performed on a separate biopsy core than the one used for snRNA or scRNA. N/A: not available*

Participant cases were adjudicated based on clinical and histopathologic data (see Methods) into acute tubular injury (ATI), acute interstitial nephritis (AIN), diabetic nephropathy (DN), hypertension-associated CKD (HT-CKD), Other (i.e. other nephropathies), and Indeterminate, which includes cases without clear features of the aforementioned groups, nor support for any other cause of kidney disease. Differences between clinical enrollment category and tissue-based adjudicated diagnoses, including unsuspected findings, have been reported previously^30–32^. To fully contextualize molecular findings, we therefore considered both enrollment categories and adjudicated diagnoses.

### Identification of acute and chronic programs in tubular epithelial cells

To understand how molecular data can inform on inter-individual variability, we next used multicellular factor analysis^33^ to identify multicellular transcriptional programs that vary across samples in a coordinated way across cell types. We included cell types located in the renal corpuscle, epithelial cells of the proximal, distal and collecting tubules, as well as interstitial cell types such as fibroblasts, immune cells, endothelial cells and vascular smooth muscle cells (including pericytes). In an analogous way to principle component analysis, factor analysis decomposes the variance in an unsupervised fashion into a set of factors that capture multicellular gene programs, where the factor score signifies how high the activity of the program is in a given sample, and the factor loading shows how a given gene (in a given cell type) contributes to the program. The total variance explained (R^2^) by the factor analysis model across all cell types is shown in Figure S4A and S5A.

To understand what biological processes were captured by the factors, we associated the resulting factor scores with categorical and continuous participant metadata (illustrated in Figure 1A). This showed two factors (2, 5) associated with clinical and histopathology data in snRNA (Figure 1B). Factor 2 explained gene expression in epithelial cells of the proximal tubule (PT, 12%), thick ascending limb (TAL, 16%), distal convoluted tubule (DCT, 23%), and collecting tubule (CNT, 23%), whereas Factor 5 described changes predominantly in PT (13%) and to a lesser degree in TAL (6%) (Figure S4A). Top up-regulated genes in these epithelial cells in factor 2 included genes such as *SPP1* and *HIF1A* (Figure S4D). Amongst others, factor 5 captured upregulation of *MMP7*, *TPM1* or *VIM* in proximal tubule epithelial cells (Figure S4E). The fuller biological contextualization of the gene expression changes captured by these factors is explored later in the results. The other factors were either technical or not directly interpretable: factor 1 reflected differences in ribosomal gene content across samples (Figure S3), factor 6 captured dosage of sex-chromosome-linked genes (Figure S4B,F), whereas factors 3, 4 and 7 were not associated with participant variables. Although some associations were shared between both snRNA and scRNA (Figure S5), most of the associations were found in snRNA. Nevertheless, factor 2 and 5 in snRNA corresponded to factors 1 and 5 respectively in scRNA, based on correlations both at gene and sample level (Figure S6). Therefore, we will focus the description on the snRNA results.

**Figure 1:**
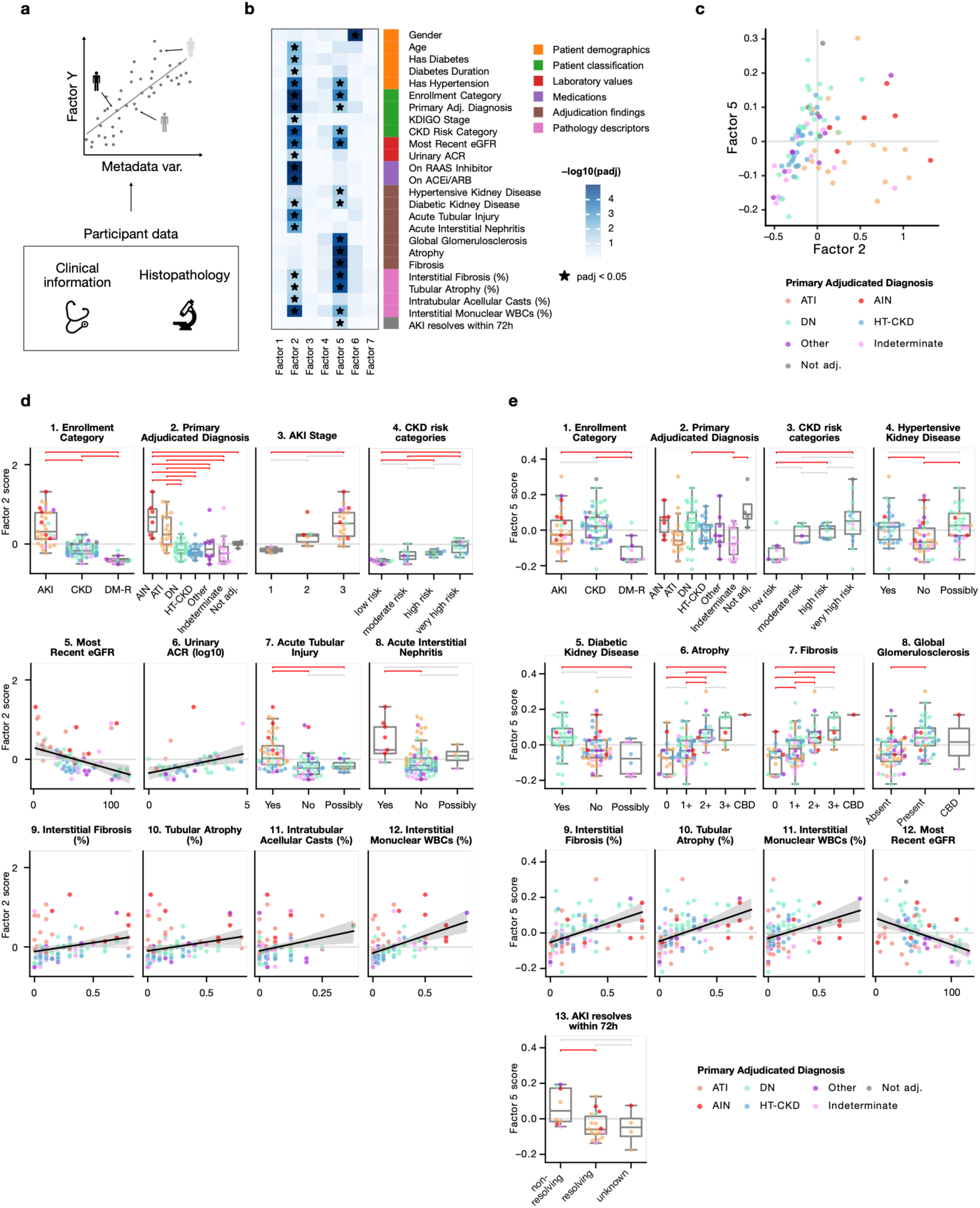
Identification of biologically interpretable factors using participant information. a. Factor scores are associated with histopathology, clinical and other available metadata b. Statistical significance of the association between sample metadata and the factor scores in snRNA. Adjusted p-values are shown based on analysis of variance or linear regression for categorical and continuous variables respectively. * marks significant associations (adj. p. < 0.05). c. Patient map based on biologically-meaningful factors from snRNA d-e. Sample metadata associated with Factor 2/5 in snRNA. Horizontal brackets show Tukey’s HSD tests in red if p < 0.05 and grey if not significant. CBD: cannot be determined.

In snRNA, factor 2 was associated with the enrollment categories (p = 1 x 10^-16^) with highest scores in AKI, followed by CKD and diabetes-mellitus resistors (DM-R) (Figure 1D1). This was consistent with adjudication diagnoses (p = 2 x 10^-8^), where acute interstitial nephritis (AIN) and acute tubular injury (ATI) had the highest scores (Figure 1D2). The factor also separated AKI (p = 0.03) and CKD (p = 5 x 10^-5^) by stage and risk categories, respectively – although the increase in score was more pronounced for AKI staging (Figure 1D3-4). These stage and risk categories based on the Kidney Disease: Improving Global Outcomes (KDIGO) guidelines quantify disease severity and are predictive of increased risk of long-term adverse outcomes^12,34^. Factor 2 was associated with eGFR (p = 0.0001) and urinary albumin-to-creatinine ratio (ACR) (p = 0.03) – measures which are used for CKD risk categorization (Figure 1D5-6). Histopathology descriptors associated with factor 2 included interstitial fibrosis (p = 0.02), interstitial mononuclear white blood cells (p = 2 x 10^-5^), tubular atrophy (p = 0.04), and intra-tubular acellular casts (p = 0.02). Given these associations and this program showing highest activity in AKI-enrolled participants, we will refer to this factor as the acute multicellular program (“acute program”).

We next investigated Factor 5 and how it is related to clinical and histopathology variables. Factor 5 also separated study participants based on enrollment categories (p = 0.0003), however participants with AKI and CKD had similar scores (Figure 1E1). Based on primary adjudicated diagnosis, participants with diabetic nephropathy (DN) had higher scores than Indeterminate (p = 0.02), which contains low-disease burden diabetes-mellitus resistors as noted above (Figure 1E2). Participants who were found to have hypertensive (p = 0.02) and diabetic (p = 0.02) changes upon adjudication also had higher factor 5 scores (Figure 1E4-5). Furthermore, factor 5 was associated with CKD risk categories (p = 0.04) (Figure 1E3), but not AKI stage (p = 0.68), and with lower eGFR (p = 0.0002) (Figure 1E12) but not urinary albumin-to-creatinine ratio (p = 0.14). Factor 5 was linked with global glomerulosclerosis (p = 5 x 10^-5^), tubular atrophy (p = 6 x 10^-6^, p = 4 x 10^-5^) and interstitial fibrosis (p = 1 x 10^-5^, p = 2 x 10^-5^), histologic features that are markers of chronic disease and predictors of progression^35–37^, both in semi-quantitative and continuous scale respectively (Figure 1E6-11). Given the association with these indicators of chronic kidney damage, we will refer to this factor as the chronic multicellular program (“chronic program”).

The chronic program separated AKI samples based on whether AKI resolved within 72 hours (p = 0.047; Figure 1E13) – assessed through serum creatinine – or not, with non-resolving AKI samples scoring higher. Correcting for the AKI stage (i.e. severity of AKI) reduced the significance of this association (p = 0.06). Since AKI resolution based on serum creatinine is linked to long-term outcomes^38^, our results suggest that the non-resolving AKI participants had a more chronic signature at the time of biopsy, which could increase their risk of progression.

Acute and chronic programs were associated with different clinical and histopathology variables, capturing changes mostly in tubular epithelial cell types. We found that the program activities spanned across all enrollment categories and adjudicated diagnoses, indicating that the acute and chronic transcriptional programs capture varying degrees of injury rather than being exclusive to one condition (Figure 1C). Therefore, the program scores were used in downstream analysis as a continuous measure of acute and chronic injury in each sample, regardless of enrollment or adjudicated categories. Importantly, while the programs were associated with glomerular and interstitial pathology findings, the program loadings best described gene expression variability in tubular epithelial cells rather than in the interstitial or glomerular compartments themselves.

### Tubular and fibroblast compositional changes in acute and chronic programs

As a next step, we examined whether the programs are related to overall changes in tissue composition. For that purpose, we investigated whether some cell types change in relative abundance based on the sample program scores (Figure 2A). In snRNA, we found that the activation of the acute program was associated with a proportional increase of immune cells and a proportional decrease of intercalated (IC), principal (PC), and vascular smooth muscle/pericyte cells (VSM/P) (Figure 2B). This decrease is likely the effect of the increase in immune cells, given the proportional nature of the data. In turn, the chronic program was associated with an increase of immune cells, but also of fibroblasts, and a decrease of PT, DCT, CNT, and IC. This is in concordance with the histopathology findings above that were scored on a different biopsy core from the same study participant^23,27,29^.

**Figure 2:**
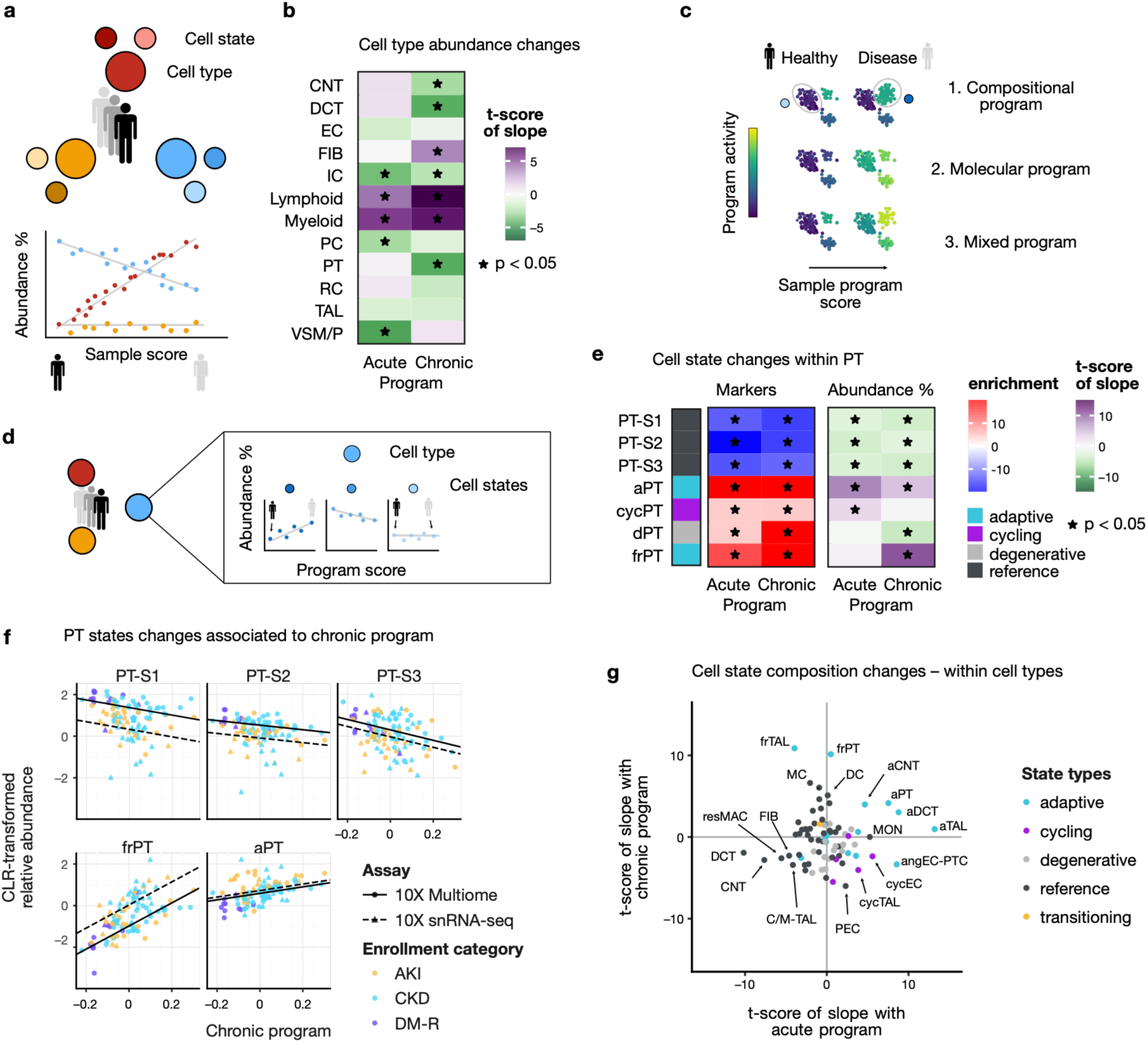
Multicellular programs capture cell state composition shifts. a. Relative abundance changes of major cell types relative to the program scores are identified with linear models b. Associations of acute and chronic programs to overall shifts in relative cell type abundance in snRNA * mark significant associations (adj. p. < 0.05) c. Multicellular programs can capture either a compositional change in terms of cell states (top), or molecular changes common to cell states (middle), or a mix of the two (bottom) d. Relative abundance changes of cell states within their respective cell type relative to the program scores are identified with linear models e. Cell state marker gene enrichment scores (left subpanel) in the program loadings and t-scores of the slopes associating (e.g., in c) within-cell type relative cell state abundance to sample program scores (right subpanel) for PT cell states in snRNA, f. CLR-transformed cell state proportion within PT relative to the chronic sample program score. Regression lines show slope of the associated mixed-effect model. g. t-scores of the slopes from mixed-effect model associating sample level acute (x-axis) and chronic (y-axis) program scores with relative cell state abundance within their respective cell types.

We next looked at which cellular processes were captured by the respective programs for each cell type. Because multicellular factor analysis uses pseudobulked data at the cell type level, it can result in three different types of programs (Figure 2C): (1) compositional programs that show high activity in specific cell states that are changing in relative abundance in disease (e.g., increase of “disease cell states”), (2) molecular programs that change in all cell states in a disease-dependent way, and (3) a mixed program where both compositional and molecular changes are captured.

To understand the compositional aspects of the programs, we examined whether marker genes of specific cell states were enriched in the program loadings. Additionally, we investigated whether cell state proportions change in relation to sample program scores (Figure 2D). In PT, marker enrichment showed that adaptive state markers were up-regulated and healthy state markers down-regulated in both programs (Figure 2E, left). Similarly, we found that the healthy states decreased as a proportion of the PT population with increasing acute and chronic scores, whereas cycling PT (cycPT) and adaptive PT (aPT) increased with the acute score, and aPT and failed repair PT (frPT) increased with the chronic score (Figure 2E, right). aPT and frPT are characterized by gene markers of early injury and unsuccessful repair respectively^20,23^. Although both aPT and frPT were found to increase together with chronic score, the increase of frPT was more pronounced (Figure 2F). Similar trends of increasing adaptive states were also seen in TAL, DCT, and CNT (Figure S7).

Even though other cell types had lower variance explained by the acute and chronic programs, we identified composition shifts correlated with program scores. Figure 2G shows the t-scores of the slopes associating the acute and chronic program scores to the relative cell state abundance within each cell type (e.g., slopes in Figure 2F, Figure S8C). In addition to the tubular changes described above, we found that the acute program was associated with an increase of cycling and angiogenic peritubular endothelial cells (cycEC, angEC-PTC) as well as classical monocytes (MON). Both programs were generally associated with decreasing healthy states of tubular epithelial cells, as well as other reference states such as fibroblasts (FIB) and resident macrophages (resMAC).

Given the association of the chronic program with fibrosis (Figure 1B, 1E9), we also looked more closely at how the program captures gene expression variability across study participants within the fibroblast population. Fibroblast cell state marker enrichment in the loadings showed that myofibroblast (MYOF) and inflammatory fibroblast (infFIB) markers were up-regulated in the acute program, and those of normal fibroblasts (FIB) were down-regulated (Figure S7A, left). Indeed, some of the genes with highest loadings were myofibroblast markers such as *COL1A1* and *FN1*^22,39^, or of activated fibroblasts such as *TNC*^40^ (Figure S4D). In contrast, the chronic program was associated with an increase of degenerative fibroblast markers (dFIB, dM-FIB), and a decrease of normal fibroblast markers. The association with the relative abundance of these states was consistent only for myofibroblasts and fibroblasts for the acute program scores (Figure S7A, right). This suggests that enrichment of marker genes might be more effective to understand fibroblast population changes at the patient level, rather than relying on detected cell state abundance. Overall, these results indicate that fibroblast reprogramming was captured in the acute program, and that fibroblast expansion – without clear changes within the fibroblast population – was co-occurring with the tubular changes in the chronic program.

In the cell population from the renal corpuscle (RC), we found an increase of parietal epithelial cell (PEC) marker genes in the acute program loadings, and an enrichment of marker genes of normal and adaptive glomerular capillary endothelial cells (EC-GC, aEC-GC) as well as of marker genes of mesangial cells (MC) in the chronic program loadings (Figure S7A). We also found that the relative abundance of mesangial cells in the renal corpuscle increased with higher chronic sample score (Figure 2G, Figure S7A). Mesangial expansion is a well-known structural feature of diabetic nephropathy^41,42^ and consistent with the association of the chronic program with global glomerulosclerosis (Figure 1D8).

### Molecular changes captured by acute and chronic programs

Having described the compositional aspects captured by the acute and chronic programs, we next asked whether there are molecular changes affecting all cell states of a cell type in a disease-dependent way. For that purpose, we quantified the activity of the program now at the *cell state-*level in each sample (Figure 3A). If a program captures only the compositional effect of one cell state increasing in abundance, one would expect a higher activity of the program in this cell state regardless of the patient disease status (as encoded by the sample program score). In the case of a purely molecular program, we would expect program activity to increase with acute or chronic program score in all cell states. A mixed program would be a combination of these two effects.

**Figure 3:**
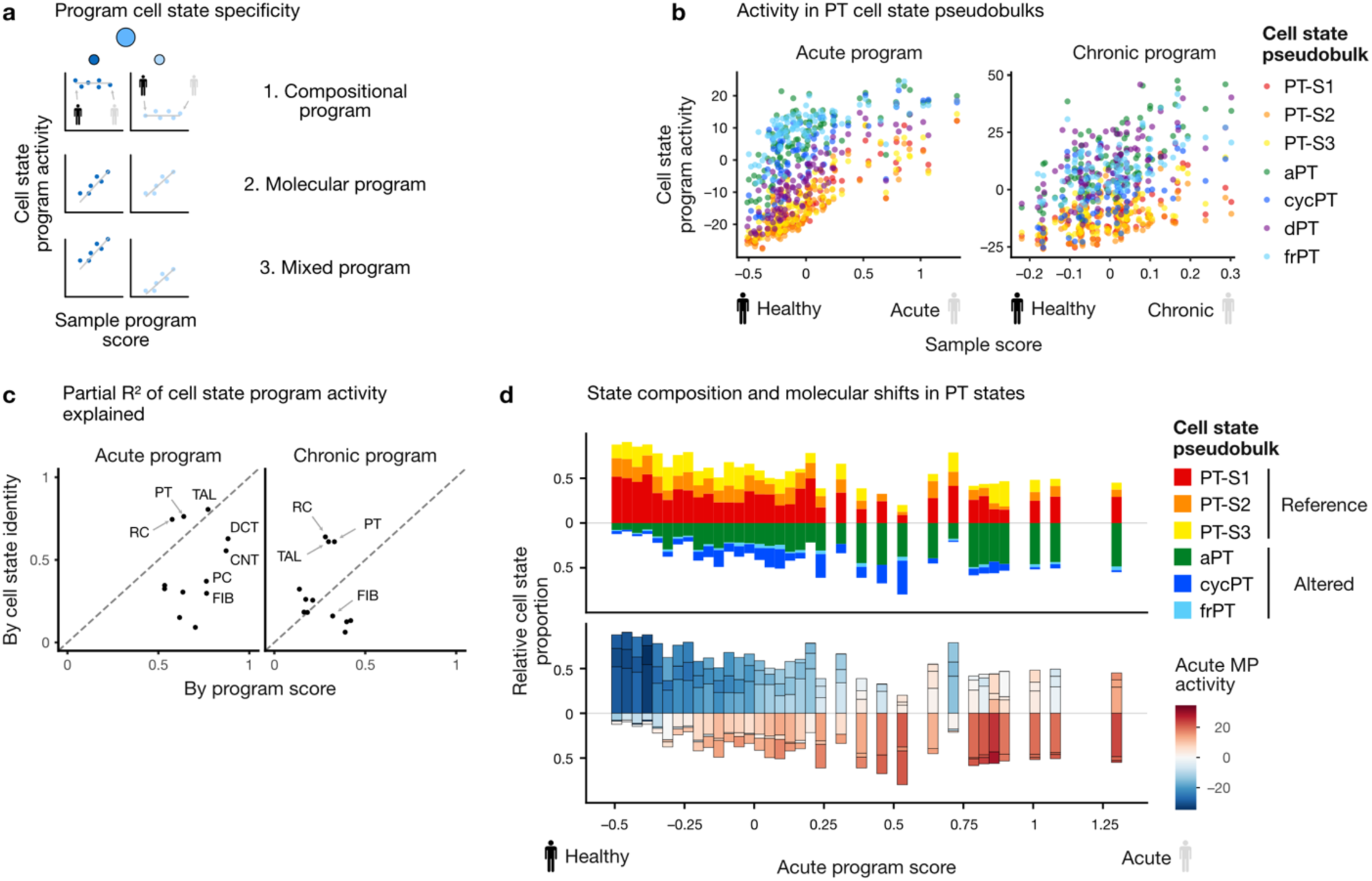
Programs capture compositional and general molecular changes in the proximal tubule. a. Programs may capture compositional changes of cell states, molecular changes common to all cell states in a disease-dependent way, or a mix of the two, b. Acute and chronic program activity in PT cell state-level pseudobulks relative to sample program score in snRNA c. Partial R^2^ contribution of cell state identity and sample program score to the prediction of program activity in cell state pseudobulks in snRNA (e.g., show in b) d. Histogram of relative PT cell state abundance (top) and acute program activity in PT cell state pseudobulks (bottom) with increasing sample acute program score in snRNA samples.

In epithelial cells of the proximal tubule, altered states (aPT, cycPT, frPT) generally showed higher activity of the acute and chronic programs than reference states (PT-S1,2,3), thus suggesting a compositional effect in both programs (Figure 3B). However, the program activity also increased in a disease-specific fashion indicating a general molecular response. In highly acute samples, reference cell states showed similar program activity to altered states in less acute samples. Similar trends were also observed in TAL cell states and in scRNA (Figure S7C, Figure S8).

To quantify the compositional versus molecular effects, we analyzed the partial R^2^ of the program activity explained by the cell state-specific intercepts and by the slope relating to the sample program score from linear models. For the acute program, the sample-level program score explained as much or more than the cell state identity in PT, TAL, DCT, and CNT (Figure 3C, left). These are the cell types whose gene expression was also best explained by the factor analysis model. Together, this suggests the acute program captured a mixture of compositional and more-widespread molecular processes (Figure 3D). In contrast, PT and TAL state identity was more important than the sample chronic program score (Figure 3C, right), thus indicating that it captured mostly compositional changes (results in previous section). For other cell types, partial R^2^ showed that the acute program captures molecular changes common to the respective cell states (Figure 3C). For instance, while myofibroblasts (MYOF) showed highest acute program activity, all fibroblast cell states had increased acute program activity in a more acute sample (Figure S7D), suggesting that this program captures a general fibroblast response.

Overall, the acute program captured a mixture of general transcriptomic changes related to acute injury as well as cell state composition shifts from healthy towards adaptive cell states predominantly within the proximal and distal tubule. Conversely, the chronic program captured compositional shifts towards failed-repair states of PT and TAL which is co-occurring with higher immune and fibroblast abundance and a lower tubular cell type population.

### Inflammatory signaling through Transcription Factor networks enhances chronic program

After describing how the acute and chronic programs relate to cell states and general molecular changes, we aimed to identify which specific biological processes they captured (Figure 4A). For that purpose, we looked at which pathways were enriched in the respective program loadings. We found that the acute program showed increased steady state levels of genes involved in epithelial to mesenchymal transition, angiogenesis, interferon alpha/gamma response, and apoptosis, as well as down-regulation of oxidative phosphorylation (Figure 4B). In the cell types best described by the chronic program (PT, TAL), we also observed higher levels of inflammatory signaling through interferon alpha/gamma and apoptosis (Figure 4C). In PT specifically, we also find increased oxidative phosphorylation, whereas it is downregulated in TAL.

**Figure 4:**
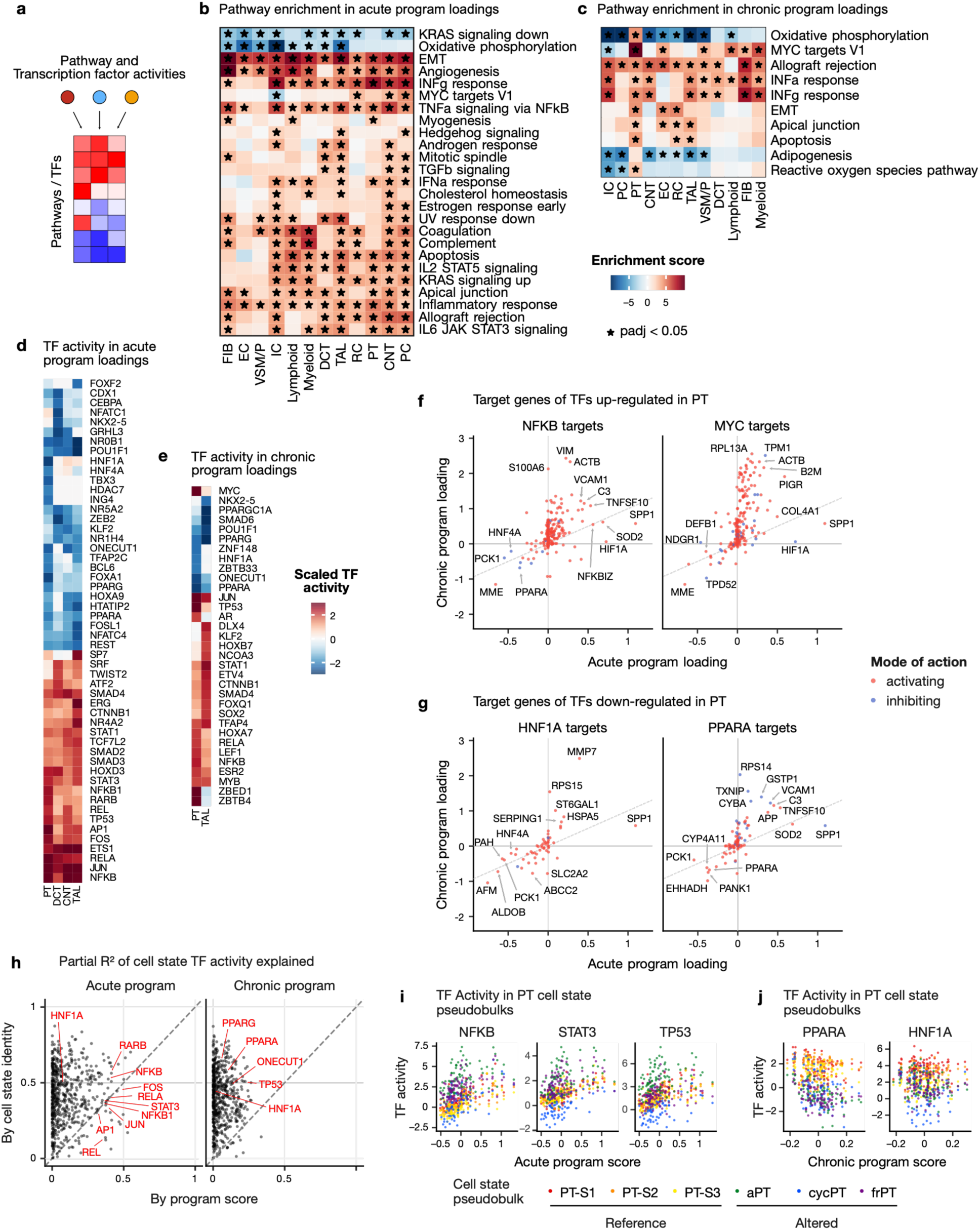
Transcriptional regulation captured by programs shows increased NFKB activity in snRNA. a. Pathways or TF activities are computed in program loadings b./c Enrichment of MSigDB hallmarks in the loadings of the acute and chronic programs respectively d./e. Top TFs in acute and chronic programs respectively in tubular epithelial cell types f./g. Loadings of target genes of NFKB, MYC, PPARA or HNF1A in acute and chronic programs. Color indicates mode of regulation where red: activated by TF; blue: inhibited. The sloped line indicates where the loadings are equal between acute and chronic loadings. h. Partial R2 contribution of cell state identity and sample program score to the prediction of TF activity in cell state pseudobulks in snRNA (e.g., shown in i-j) i./j. TF activity in PT cell state-level pseudobulks in relation to their respective sample acute/chronic program score

To get more granular insights of the putative regulatory mechanisms captured by the programs, we examined which transcription factors (TFs) could explain the observed gene expression changes by being differential active. In tubular cell types, the acute program was characterized by higher activity of inflammation-related transcription factors, including genes from the NFkB complex (*NFKB*, *NFKB1*, *RELA*, *REL*), the AP-1 complex (*JUN*, *AP-1*, *FOS*) and *STAT1/3* (Figure 4D). These transcription factors have been shown to be involved in PT and TAL cell state trajectories from healthy to adaptive and failed-repair states in the human kidney ^23,24^. Furthermore, inhibition of AP-1 has recently been shown to reduce inflammation and fibrosis in a mouse model of ischemia-reperfusion^25^. The chronic program showed higher activity of *MYC* and *TP53* (PT), as well as lower activity of *HNF1A* and *HNF4A* (PT), as well as *PPARA* and *PPARG* (PT and TAL) (Figure 4E). Additionally, we found that the transcription factor activities in the acute and chronic programs are positively correlated in PT and TAL loadings (Figure S9A). Specifically, we observed that TFs of the NFkB, AP-1 complex, the STAT family and *TP53* had positive activity, whereas *PPARA*, *PPARG*, *HNF1A* and *ONECUT1* had negative activity in both programs. A similar pattern was also observed in scRNA (Figure S9B). This suggests that similar mechanisms are at play in both programs, but that they have an additive effect at the patient level.

Amongst target genes of *NFKB* (Figure 4F, left), *SPP1* and *HIF1A*, as well as *PCK1* and *HNF4A* were more strongly up- and down-regulated respectively in the acute than the chronic program loadings. This is consistent with the known mode of regulation of *NFKB* on these targets^43^. However, many injury markers such as *VIM*, *ACTB*, *VCAM1,* and *C3* were strongly upregulated in the chronic program^23^ (Figure 4F). A similar pattern is also observed for *MYC* targets (Figure 4F, right). In turn, TFs with negative activity in both programs such as *PPARA* and *HNF1A* had targets that were more strongly downregulated in the chronic program (Figure 4G). This suggests that while the effects of acute injury – as captured by the acute program – are seen in the regulatory activity of TFs such as *NFKB* and *MYC*, chronic progression – as captured by the chronic program – has an additive effect by expanding the number of target genes affected. Furthermore, these results suggest a direct link between *NFKB* activity and *PPARA* down-regulation at patient level. Indeed *NFKB* is a transcriptional repressor of *PPARA* (Figure 4F, left) and these two TFs have antagonistic effects on several target genes (e.g., *VCAM1*, *SPP1*, *PCK1*) (Figure 4F-G). In such cases, the effect of *NFKB* activity could be further increased through the transcriptional inhibition of its antagonist *PPARA*. *PPARA* is known as a regulator of lipid metabolism and has been discussed as an important antagonist of *NFkB* and *AP-1* activity, as well as TGFꞵ-signaling in chronic kidney disease^44^.

After finding TFs that were putative regulators of the transcriptional changes captured by the programs, we again asked whether their activity is related to changes in cell state composition, or a shared molecular response across cell states. Consistent with the results in the previous section, we observed that top TFs in the acute program were both disease and cell state driven, whereas TFs in the chronic program –including *PPARA*– were mostly cell-state dependent (Figure 4H). We found that TFs with up-regulated activity in the acute program generally had higher activity in altered cell states of PT (aPT, cycPT, frPT) but that the activity also increased with acute program score (Figure 4I). In contrast, *PPARA*, and *HNF1A* had highest activity in reference states and lower in altered, with little or no effect of the sample program score (Figure 4J).

Taken together with the observations on cell state shifts in the previous section, these results suggest that the general transcriptomic changes captured by the acute program not only affect reference and adaptive states alike but might also directly enhance the transition towards adaptive and ultimately failed repair states, thereby linking acute and chronic trajectories. Furthermore, given the association of the chronic program with global glomerulosclerosis and tubular atrophy, this suggests that the transition towards failed repair states is related to irreversible chronic damage.

### Urine and plasma biomarkers correlate with acute and chronic program scores

To get an orthogonal validation of the transcriptomic findings described above, and to assess their potential translational value, we investigated whether the program scores are associated with plasma and urine protein abundances obtained at time of the research biopsy. To identify proteins associated with these processes of interest, we predicted the acute or chronic program scores of participants from the protein abundance data measured with the SomaScan aptamer-based proteomic platform^45–47^, which was available for a subset of participants with snRNA (n = 69 urine, n = 62 plasma; Table S2). In cross-validation, these models were able to order participants by acute and chronic program score based on protein abundance in plasma (mean spearman correlation of 0.59, and 0.47 respectively) or urine (0.78 and 0.40, respectively) (Figure S10A).

The plasma proteins predictive of the acute program scores included well-known markers of injury, such as SPP1 and HAVCR1 (KIM1) ^48–50^(Figure 5A). Assuming that plasma protein changes reflect tissue expression changes, we next examined the expression patterns of these genes in the transcriptomics data. *SPP1* was expressed in the majority of cell types (Figure S10B), and in PT cells showed highest expression in the degenerative state (dPT). We found that *SPP1* expression increased in all PT states together with sample program score (Figure 5C, top). *HAVCR1* expression was PT-specific and was highest in aPT and frPT while also increasing with acute program score (Figure 5C, bottom, Figure S10B). Other top plasma predictors included LRCH4 (involved in innate immune response^51^), ANG (involved in angiogenesis in kidney injury^52–54^), CSF1 (involved in macrophage reprogramming^55,56^), and CCL14 (a urinary biomarker associated with persistent AKI^57,58^). However, the snRNA data was too sparse to investigate their cell type-specific expression. The urine proteins predictive of the acute program included TNC/Cr, a marker of activated fibroblasts known to promote fibrogenesis^23,40,59,60^, and CCL2/Cr (MCP-1), a marker of inflammatory fibroblasts, inflammatory peritubular endothelial cells, and adaptive PT cells^23,61–64^ (Figure 5A, S11A). In fibroblasts, these genes exhibited both a cell state-specific expression and were increasing across states together with higher sample acute scores (Figure 5D). Other top increasing urinary proteins included LAMC2/Cr, an aTAL and frTAL marker, THBS1/Cr, expressed in inflammatory fibroblasts, and SERPINE2/Cr, which was most highly expressed in endothelial cells, degenerative and inflammatory fibroblasts, and has recently been hypothesized to promote collagen deposition by inhibiting its degradation in an in-vitro model of kidney fibrosis^65^ (Figure S11A). Proteins decreasing in abundance in the urine with higher acute scores included FLRT2/Cr, which is most highly expressed in endothelial cells (Figure S11A), where it prevents senescence^66^, and GNS/Cr, a lysosomal enzyme involved in heparan sulfate metabolism^67^. The expression of GNS was too sparse to derive any more insights into its expression patterns across cell types. Taken together, these results indicate that plasma and urine proteins associated with the acute program reflect tissue inflammation, fibrosis together with a loss of markers linked to both vascular and metabolic homeostasis.

**Figure 5:**
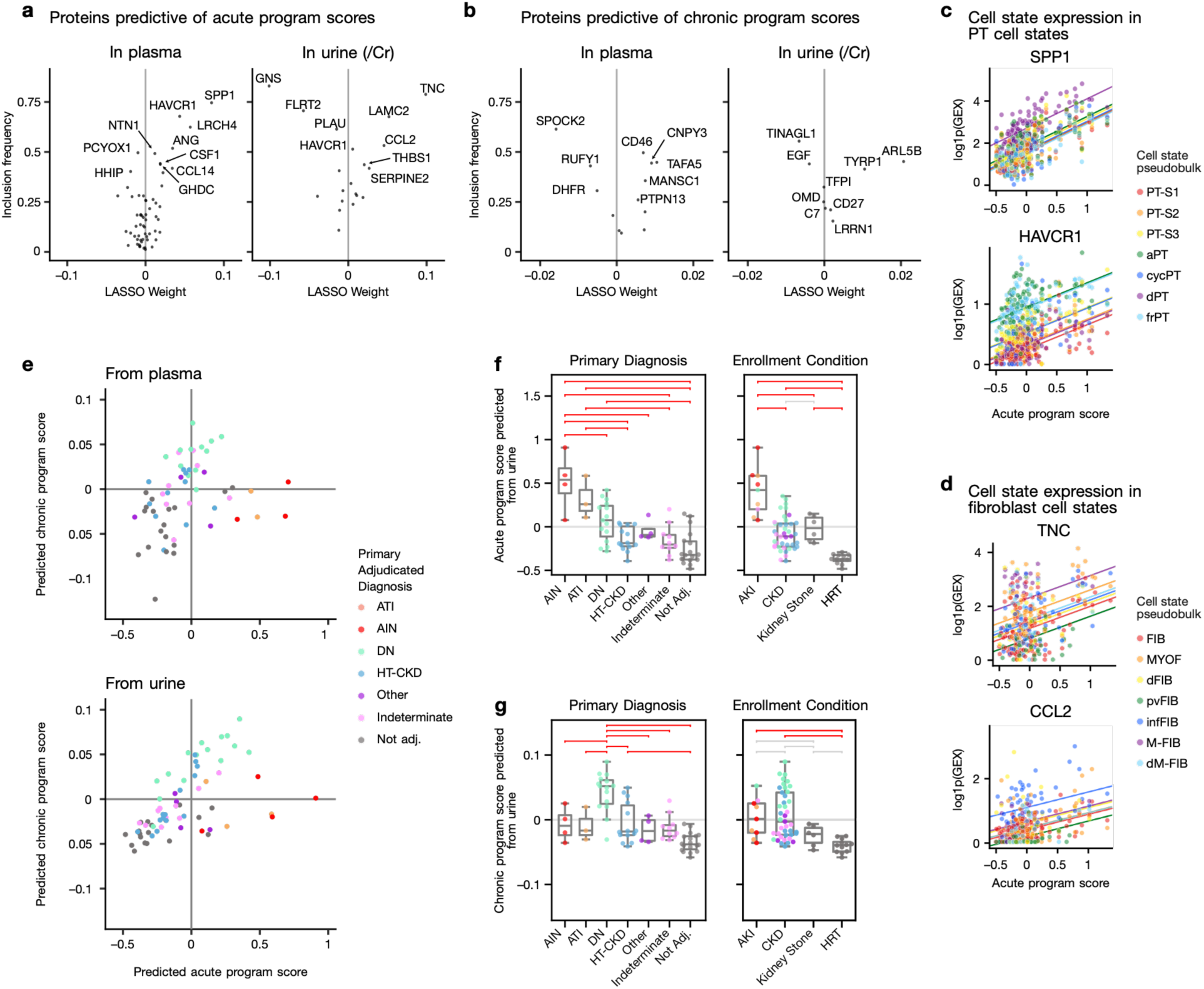
Plasma and urinary kidney damage protein reflect intra-renal acute and chronic program scores. a. Weights of plasma and urine proteins used to predict sample acute program scores and inclusion frequency in bootstrap. In all analyses urine proteins are normalized to creatinine (/Cr). Proteins are labeled by HGNC gene symbols. c. SPP1 and HAVCR1 expression in PT cell state pseudobulks relative to acute program score. Slopes show the relation between gene expression and acute program score with an intercept for each cell state, as determined by a mixed effect linear model, d. TNC and CCL2 expression in fibroblast cell state pseudobulks relative to chronic program score. Slopes show the relation between gene expression and acute program score with an intercept for each cell state, as determined by a mixed effect linear model e. Acute and chronic program scores predicted from plasma (top) and urine (bottom) for participants without transcriptomics data. f./g. Predicted score from urine with respect to primary adjudicated diagnosis and enrollment category, for acute and chronic programs respectively. Horizontal brackets show Tukey’s HSD tests in red if p < 0.05 and grey if not significant. CBD: cannot be determined.

For the chronic program, plasma proteins increasing with chronic scores included CNPY3, a marker gene for immune cells (Figure S10B) associated with Toll-like receptors and inflammasome activation^68,69^, and CD46, a complement inhibitor ubiquitously expressed in the kidney (Figure S10B) which has been associated with better transplant outcomes^70,71^. This observed increase in CD46 may be driven by increased STAT3 signaling^72^ captured in our transcription factor analysis of the acute program. Plasma proteins whose decrease was predictive of the chronic program included SPOCK2, a podocyte marker gene (Figure S10C) and marker of kidney health^73^, as well as RUFY1, which plays a role in endosome trafficking and intracellular trafficking of both SLC2A1 and activated EGFR^74–76^. In the urine, higher abundance of ARL5B/Cr, a protein involved in retrograde transport^77^, and lower abundance of EGF/Cr were predictive of higher chronic scores. *EGF* expression in the tissue was specific to DCT and TAL, where it was more highly expressed in healthy TAL states than in aTAL and frTAL (Figure S11B). Furthermore, *EGF* expression decreased across all TAL states at higher chronic program scores (Figure S11C). This decrease of urinary EGF/Cr has previously been shown to be associated with CKD progression, whereas its return to normal levels was predictive of recovery in IgA nephropathy patients^78,79^. Similar to the acute markers, plasma and urinary proteins associated with the chronic program capture a loss of healthy markers of the glomerulus and tubules together with increased inflammatory signaling.

To validate these programs within the KPMP cohort, we used the linear models to infer acute and chronic program scores for individuals that were not included in the snRNA, nor in the training of the linear models (plasma = 59, urine = 68, Figure 5E). This included healthy participants (HRT), as well as individuals with kidney stones. The inferred scores separated participants based on enrollment categories, adjudication and pathology descriptors for the models using urine or plasma proteins, in a manner consistent with our findings in the transcriptomics data (Figure S12-13). AKI-enrolled participants scored highest in the acute program, and DN-adjudicated participants scored highest in the chronic factor, with HRT samples scoring the lowest in both programs. Based on urinary proteins, participants with kidney stones scored higher in the acute program than HRT (Tukey’s HSD p = 0.002), to a similar level to CKD participants (Figure 5F), consistent with higher stress and immune response in the kidney, as has been reported previously^80^. The chronic score did not separate individuals with kidney stones from the HRT group (Tukey’s HSD p = 0.8; Figure 5G, S13C). Plasma and urine measurements were available in the same individuals for 59 participants, where the predicted scores were correlated for both the acute (Pearson’s r = 0.72, p-value = 1 x 10^-9^) and the chronic program (Pearson’s r = 0.66, p-value = 1.3 x 10^-8^), thus showing high concordance between independent measurements (Figure S14A-B).

### Tissue-derived programs are predictive of baseline kidney function and incident renal events in external cohort

To test whether kidney tissue-derived multicellular programs are detectable as circulating molecular signals beyond the KPMP biopsy cohort, we predicted acute and chronic scores, using the plasma proteins identified above, in the UK biobank where plasma proteins were measured using a different technology, the antibody-based Olink proximity extension assay platform^81^. The population in the UK biobank cohort with plasma measurements (n = 45,167) was older (mean age = 56.8 ± 8.2 years), had more female participants (54% female), and had better kidney function (baseline mean eGFR 94.2 ± 13.6 mL/min/1.73m^2^) than the KPMP cohort, and only 2% (N = 917) of participants met criteria for CKD (eGFR < 60 mL/min/1.73m^2^) at enrollment. These participants also had significantly higher rates of hypertension and cardiovascular disease (Table S3). The predicted acute and chronic scores showed continuous distributions across the cohort and were positively correlated (Pearson r = 0.542, p < 0.001, Figure S15), which is consistent with observations in the KPMP cohort.

Despite differences in proteomic platforms between the KPMP and UK biobank, both predicted program scores showed significant inverse associations with baseline eGFR in multivariable-adjusted models (Table S4). In the fully adjusted model including all participants, each standard deviation increase in the acute or chronic program score was associated with lower eGFR (β_acute_ = −2.92, p < 0.001; β_chronic_ = −3.45, p < 0.001). When both scores were included in the same model, both remained independently associated with eGFR (β_acute_ = −0.47, p < 0.001; β_chronic_ = −4.66, p < 0.001). Stratified analyses revealed differential associations by baseline CKD status. At baseline, the acute program showed stronger association among participants with CKD (eGFR < 60 60 mL/min/1.73m^2^) than without CKD (β_acute_ = −3.35 vs. −1.64, p-interaction < 0.001). In contrast, the chronic program maintained consistent associations across CKD strata (p-interaction = 0.462, Table S4). We also observed a steeper decline in eGFR relative to both acute and chronic scores at higher than average scores (p < 0.001 for deviation from linearity; Figure 6A), which is consistent with a loss of filtration in individuals with higher underlying tissue injury. Conversely, the slower decline at eGFR ≥ 90 mL/min/1.73m^2^ may partly reflect imprecisions in the eGFR measurement^82–84^.

**Figure 6:**
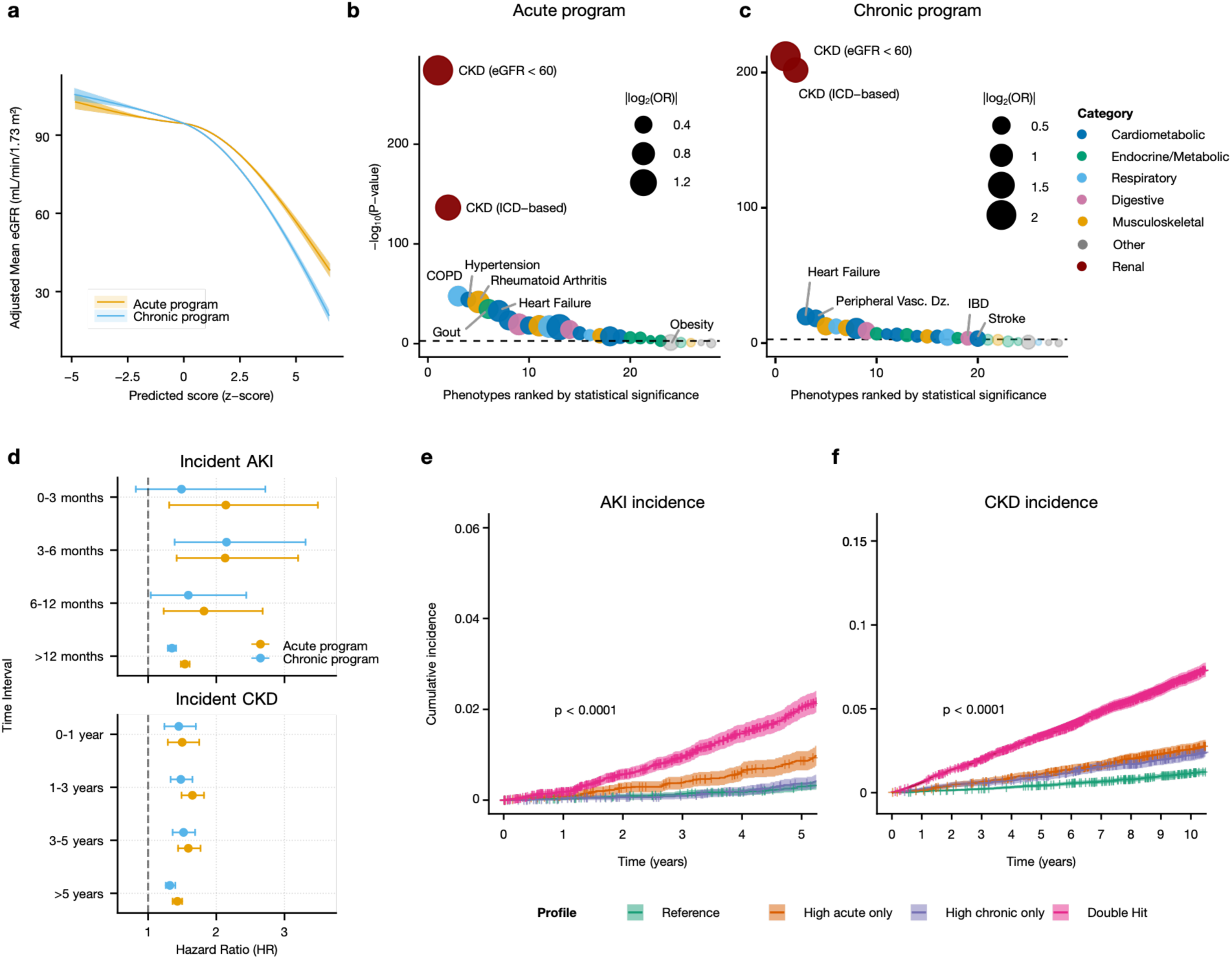
Circulating proteomic projections of kidney injury programs are associated with organ function and clinical outcomes in the UK Biobank. a. Restricted cubic spline curves showing adjusted mean eGFR as a function of predicted acute and chronic program scores (z-standardized) in UK Biobank participants with plasma Olink data (n = 45,167). Models adjusted for age, sex, BMI, smoking, baseline comorbidities, and RAS inhibitor use. b/c. Phenome-wide association of acute (b) and chronic (c) program scores with 28 prevalent conditions at baseline. Phenotypes are ranked by statistical significance (y-axis); dot size reflects log₂ odds ratio, color indicates disease category. Adjusted for age, sex, BMI, smoking, and RAS inhibitor use. d. Hazard ratios for incident AKI (top) and CKD (bottom) based on acute and chronic program scores (z-standardized). Cox models adjusted for age, sex, BMI, smoking, comorbidities, RAS inhibitor use and baseline eGFR. Brackets show 95% confidence intervals. e/f. Cumulative incidence of AKI (e) and CKD (f) among 44,250 participants without baseline CKD, stratified by median-split program scores into four profiles: Reference (low/low), High Acute Only, High Chronic Only, and Double Hit (high/high). Cox models adjusted for age, sex, BMI, smoking, comorbidities, RAS inhibitor use, and baseline eGFR.

To assess whether the predicted scores capture kidney-specific pathobiology or instead reflect systemic plasma protein alterations caused by other co-existing conditions, we examined their associations with baseline prevalence of 28 common chronic conditions spanning cardiometabolic, pulmonary, inflammatory, and degenerative disease categories (Figure 6B-C). The acute and chronic scores had the strongest association with chronic kidney disease (based on baseline eGFR; OR_acute_ = 3.04 per SD, p < 0.001; OR_chronic_ = 4.14, p < 0.001), showing that the plasma proteins preferentially captured renal signals. Among extra-renal conditions, the strongest associations were observed for atherosclerosis and rheumatoid arthritis, where the effect magnitude remained 30-45% lower than the CKD signal.

Having shown the cross-sectional associations of the programs with tissue injury and baseline kidney function, we next asked whether the predicted scores also translate to increased longitudinal risk of renal events. Among 44,250 participants in the UK biobank without CKD at baseline (eGFR ≥60 mL/min/1.73m^2^), 2,391 incident (after enrollment) AKI events and 2,315 incident CKD events accrued over a median follow-up of 13.81 years. In multivariable piecewise Cox models adjusted for age, sex, BMI, smoking, baseline diabetes, hypertension, heart failure, prior myocardial infarction, RAS inhibitor use, and baseline eGFR, higher predicted acute and chronic program scores consistently predicted incident renal outcomes, with evidence of time-varying effects (Figure 6D, Table S5-6). For incident AKI, the acute scores showed strongest association within the first 6 months (HR = 2.14) and remained predictive but with attenuated risk in long-term follow-up (>1 year), whereas the chronic score was not predictive for the first three months and instead had peak association at 3-6 months (HR = 2.15) with persistent risk beyond 1 year. Both acute and chronic scores were associated with increased risk of CKD over the whole follow-up timespan, with the acute score showing highest association at 1-3 years (HR = 1.65) and the acute score at 3-5 years (HR = 1.52) and thus suggests that the plasma-derived program scores can give insights into underlying tissue injury before it impacts eGFR measurements and that they are linked to both early and sustained risk of AKI and CKD.

Based on our observations in the KPMP transcriptomics and plasma analyses, we hypothesized that the programs capture distinct but co-occurring kidney injury processes that when present together have an additive effect on risk for AKI or CKD. We therefore stratified the cohort using a threshold at the median acute and chronic program scores respectively into four groups: Healthy/Reference (low acute, low chronic; n = 14,601), High Acute Only (n = 7,524), High Chronic Only (n = 7,524) and Double Hit (high acute, high chronic; n = 14,601) and assessed the association with incident AKI and CKD. Participants of the High Acute Only and Double Hit groups had a higher risk of incident AKI, but not the High Chronic group (Figure 6E, Table S7). The Double Hit group exhibited the highest AKI risk (HR 2.13), consistent with additive risk when both acute injury and chronic damage programs are elevated. Risk for CKD was increased for all three non-reference groups, with High Acute or Chronic Only having similar hazards ratio (HR_acute_ = 1.41; HR_chronic_1.31) and the strongest association again in the Double Hit group (HR = 2.23; Figure 6F, Table S7). These results extend the time-resolved analyses and show that while the acute and chronic programs separately increase the risk of future adverse renal events, their co-occurrence in an individual confers the highest risk.

## Discussion

In this study, we show that unsupervised, patient-centric analysis of single-cell transcriptomes from kidney biopsies from KPMP participants with AKI and CKD identifies two multi-cellular programs that capture molecular changes characteristic of acute injury and chronic damage. These programs describe compositional shifts toward adaptive epithelial cell states in the proximal and distal tubule. Although mainly describing epithelial changes, their activity correlates with immune infiltration and fibrosis, as assessed by cell state abundances in the single-cell data and pathology, respectively. The activation of these programs across individuals with AKI and CKD underscores and gives a molecular basis to the features of tubular damage that these conditions share, irrespective of the initiating event^6,7^. The scores of the acute and chronic program can therefore be used as unbiased continuous measures of acute and chronic damage respectively. Furthermore, we find that the acute program is activated broadly across cellular states in the tubule, with the strongest activation in adaptive states. In contrast, the chronic program is more specific to altered states, particularly failed repair states.

Previous studies have characterized these cellular state transitions in detail^19–21,23–25^. Our results extend these findings by showing that these regulatory changes directly explain patient variation as continuous processes of acute injury and unresolved tubular damage across AKI and CKD. This approach – identifying coordinated cell type changes across patients – has also been applied successfully in glomerular diseases, where we observed similar molecular changes in the proximal and distal tubule associated with kidney function, interstitial fibrosis and tubular atrophy^85^. Beyond the pattern of tubular injury previously described, the present study was able to distinguish two inter-related programs of acute and chronic injury through the inclusion of AKI participants. We were also able to link these programs to non-invasive surrogates in the urine and plasma within the KPMP and in an independent cohort (UK Biobank).

In conventional single cell transcriptomics, integration methods are designed to remove systematic differences between samples to correct for batch effects for the purpose of cell type annotation^86^. Our results indicate that a part of these systematic differences correspond to true biological variation between samples, in a manner consistent with disease severity. Although the acute program is highest in aPT cells, it is elevated across all PT states in highly acute samples. Consequently, integration methods may lead to underestimation of the expansion of aPT cells in individuals with a more acute signature. This has important implications for understanding the response to stress of cells in the proximal tubule, as adaptive states are a decision point between resolving injury and failed repair^23,26^. Our analysis supports the hypothesis that irreversible tubular damage and accumulation of failed-repair cell states might be a probabilistic transition, influenced by the frequency, intensity or persistence of tubular cell stress^7^.

To translate these tissue-derived programs to clinically meaningful and accessible measures, we used the paired snRNA-biofluid proteomics data available for the KPMP cohort. Taking advantage of the unique research biopsy setting in KPMP, we were able to build out a non-invasive signature of the tissue level acute and chronic program scores from urine and plasma proteins for participants where both data modalities were available at time of biopsy. The relationship between biofluid protein abundance and histologic features of acute and chronic injury held even in participants without transcriptomic data, and therefore unseen by the original model, supporting the robustness of the findings. Several of the most important proteins for predicting program scores, namely KIM-1 and MCP-1, EGF and testican-2, have previously been associated with long term renal outcomes^87,88,78,73^. Our analyses allowed us to identify the potential molecular and structural changes with which these biomarkers are associated. Extending this approach to the plasma proteomics data of the UK biobank showed that our signatures were not only associated with baseline kidney function, but also predictive of long-term AKI and CKD incidence. Individuals with elevation of both acute and chronic program scores were most at risk, further supporting the hypothesis of the additive effect of co-occurring injury processes in the CKD-AKI continuum^7^. Conversely, this also indicates that circulating proteins reflect the degree and type of kidney injury, which in turn shapes renal outcomes. Results such as these provide a mechanistic justification for the translation of our tissue-derived programs to non-invasive and cost-effective biomarker profiles. This would enable improved assessment of an individual’s position on the AKI-CKD continuum with implications for long term kidney function, maybe even without a biopsy. Future applications will focus on the use of tissue, plasma, and urinary markers of treatment response to protect against further damage.

This work has two main limitations. First, longitudinal outcome data are not yet available for KPMP participants with single cell transcriptomic profiling, for now preventing insights into how the multi-cellular programs relate to future events directly in the KPMP biopsy cohort. Second, the acute program was associated with clinical variables such as age and drug treatment (Figure 1B, Figure S5C), likely reflecting confounding with enrollment categories. We addressed these limitations by predicting acute and chronic program scores in the UK biobank using plasma proteins, where longitudinal outcome data is available and the predicted scores remained associated with incident AKI and CKD.

Collectively, our findings suggest that multicellular molecular programs can capture the critical processes of kidney injury progression across the AKI-CKD disease categories. Our results further demonstrate that this continuous and molecular measure of the type and degree of kidney injury provides a framework to identify novel plasma and urinary biomarkers enabling patient stratification based on intrarenal cellular programs rather than broad descriptive syndromic classifications.

## Methods

### Ethical compliance for human studies

We followed all relevant ethical regulations relating to human studies for this work. We include human single nuclei/cell RNA data from the Human Kidney v2 atlas^23^. This includes kidney biopsies, urine and plasma samples that were obtained with informed consent by the Kidney Precision Medicine Project under protocols approved by the University of Washington Institutional Review Board (IRB# 20190213) and samples from the Human Biomolecular Atlas Program (HuBMAP) that were collected under a protocol approved by the Washington University Institutional Review Board (IRB# 201102312). Samples from the RENAL-HEIR (NCT03584217, IRB# 16-1752), IMPROVE-T2D (NCT03620773, IRB# 18-0704) and CROCODILE (NCT04074668, IRB# 19-1282) trials are also used in this study. Data in the UK Biobank was collected with approval by the North-West Haydock Research Ethics Committee (11/NW/0382, 16/NW/0274, 21/NW/0157).

### Adjudication of samples and TIV scoring

The adjudication of cases was made by the KPMP Biopsy Adjudication Committee, consisting of nephrologists and nephropathologists from the consortium. It combines a rigorous review of the clinical case presentation, as well as a histology review of light microscopy, immunofluorescence and electron micrographs. The clinical and pathology reviews are followed by an in-committee discussion of the case where members select a consensus primary diagnosis of “diabetic nephropathy”, “hypertension-associated kidney disease”, “indeterminate” or “other”^31,89^.

Tubulointerstitial and vascular scoring descriptor scoring in KPMP was developed based on the NEPTUNE Digital Pathology Scoring System^90^. The descriptors are scored by two pathologists within KPMP: a primary and a quality control scorer. They review and score stained sections (H&E, PAS, trichrome and Jones silver histochemical stains, two of each), and resolve scoring discrepancies through adjudication, with the potential involvement of a third pathologist if necessary.

### MCFA on pseudobulk profiles

We use the combined snRNA-seq and snMultiome data and the scRNA from the HKA v2 atlas^23^. They are available on cellxgene (https://cellxgene.cziscience.com/collections/9c9d04c4-8899-417f-bb6f-6107dcadf14f). Data generation and cell type annotation is described in the atlas publication^23^. We harmonized labels of the major cell types (so-called “subclass level 1” in HKA v2) to have similar definitions across omics (e.g., lymphoid and myeloid have different granularity in snRNA vs. scRNA), and combine cell types expected to be represented only in the renal corpuscle (aEC-GC, EC-GC, MC, POD, PEC). To create cell type pseudobulk profiles, we aggregate the count data by summing the counts of cells of a given cell type for each sample. These were filtered to keep only cell type profiles based on at least 25 cells, with at least a 1000 total counts, using the adata_to_views function from the liana-py package^91^. The gene space was filtered independently for each cell type using edgeR’s approach to retain only genes with sufficient counts across samples, using decoupler-py’s implementation^92^. We filtered out cell types that were represented in fewer than 50% of the samples and excluded samples that had data for less than 50% of the remaining cell types measured. We also manually ensured that both omics had similar cell types included in the model. Finally, the data was normalized and log1p transformed prior to running MCFA.

MCFA was run using the MOFA+ implementation^93^ in the muon package^94^ with 7 latent dimensions and medium convergence mode. For snRNA data, it was run in a grouped fashion and on highly variable genes common to both 10X snRNA and 10X snMultiome data in the snRNA atlas, whereas in scRNA it used all highly variable genes. The variance explained by the model for each cell type is obtained directly from the MOFA model. For ease of interpretation, we orient the factors so that reference samples (i.e. DM-R) have the lowest factor values, by switching the sign of both the factor loadings and scores when necessary.

### Association of MCFA factors with metadata variables

Sample metadata (i.e. clinical, adjudication and histopathology descriptor scores) were associated to the factor scores using either ANOVA for categorical variables, or linear regression for continuous variables using the statsmodels package^95^. Samples with missing metadata information were filtered out for each variable separately, and categorical levels were excluded from ANOVA if they had fewer than 3 samples. Missingness was independent from enrollment or primary adjudicated category for all variables with the exception of nephropathy-specific information (e.g., diabetes/hypertension occurrence and duration, not shown). The p-values were adjusted for multiple testing using the Benjamini-Hochberg method. For significant categorical variables, we report the p-values of Tukey’s HSD test for post-hoc analysis in order to identify levels with different sample means.

### Gene set enrichment analyses

Gene set enrichment was generally computed using the univariate linear model (ULM) method from decoupler-py^92^.

Pathway activities in program loadings were computed using MSigDB hallmark gene sets^96^ with weights set to 1 for each gene in the genesets. The genesets were accessed through the decoupler-py package^92^.

TF activities, in program loadings or cell state (so-called “subclass level 2” in HKA v2^23^) pseudobulks, were computed based on the TF-target gene interactions and weights from CollecTRI^43^, which were retrieved through the decoupler-py package^92^.

Cell state marker enrichments in program loadings were computed using marker genes of cell states within their respective cell type (e.g., aPT within PT, one versus all others) obtained using the rank_genes_groups function from scanpy^97^. For each cell state, we used the top 200 genes ranked by smallest adjusted p-value and biggest t-statistic and used the t-statistic as weights.

Program activities in cell state pseudobulks were computed using gene loadings of the program as weights, using the respective cell type when appropriate. In that case, only genes with loadings with absolute value greater than 0.1 were included for the analysis. The p-values were adjusted for multiple testing using the Benjamini-Hochberg method when appropriate.

### Cell type and state abundances association with factors

Relative cell type abundances or relative cell state abundances within their respective cell type were computed and center-log ratio (CLR) transformed with scikit-bio’s implementation^98^. We also use its multiplicative_replacement function to deal with zero proportions. For downstream analysis, we exclusively use the patient-cell type combinations that were used in the MCFA analysis.

To understand how factor scores describe composition shifts, we predict clr-transformed abundances with mixed-effect models where factor scores are modelled as fixed effects and the assay as random intercept (i.e. 10X snRNA or 10X Multiome), using the statsmodels package^95^.

For the snRNA data, we use factor 1 (to account for ribosomal effect), 2 and 5 scores as fixed effects, and only factor 1 and 5 scores in scRNA. We use the Benjamini-Hochberg method for multiple-testing correction.

### Modelling of cell state- vs. disease-specificity

To understand whether disease processes – gene expression, TF activity or program activity – behaves in a cell state- or disease-specific manner, we use lmer from the lme4 R package^99^ to fit linear mixed effects models where the cell state identity and program/factor score are modelled as fixed effects, and a random intercept is computed per patient per assay (if applicable, i.e. 10X snRNA or 10X Multiome). Partial R2 values for the fixed effects are computed using the r2glmm R package^100^.

### Prediction of program scores from plasma and urine SomaScan data

We use the plasma and urine Somascan data from KPMP available at atlas.kpmp.org/repository. The urine protein data is first normalized against urine creatinine. The urine and plasma data is then log2-transformed. We filter the data to include aptamers and samples that pass quality control as described in the attached data sheets. We only include aptamers that target proteins whose genes were included in the MCFA model in at least one of the cell types. For samples with both snRNA and urine or plasma Somascan data, the program scores from MCFA are predicted by LASSO regression (with sklearn) using a 3-fold cross-validation scheme to estimate performance. Train and test fold plasma/urine proteins are scaled within the cross-validation based on the train fold. A final model is then similarly fitted on the entire scaled dataset. This is repeated for each combination of urine/plasma and acute/chronic program scores.

To assess the stability of the final model coefficients, we use bootstrap with replacement of samples (with 500 iterations) to compute how the likelihood of each protein/gene in the final model to be selected, as well as the mean and standard deviation of the regression coefficients over the bootstraps. For the figures, we show only the 20 most stable coefficients based on a stability measure computed as the product of the inclusion likelihood and the mean weight, divided by the standard deviation of the model weight.

Using these models, we predict the acute/chronic program scores for samples without transcriptomic information and that were included neither in the fitting of the MCFA nor the LASSO models. These predicted scores were then associated to participant metadata using ANOVA or linear regression as explained above.

### External validation of projected acute and chronic programs in UK Biobank Study population and plasma proteomics data

To validate the acute and chronic kidney injury programs in an independent large-scale cohort, we utilized data from the UK Biobank, a prospective population-based study of approximately 500,000 participants aged 40-69 years at recruitment between 2006-2010. We included participants with available plasma proteomic measurements from the Olink platform and complete clinical data for baseline kidney function assessment. Baseline estimated glomerular filtration rate (eGFR) was calculated from serum creatinine using the CKD-EPI 2021 equation. Participants were excluded if they had missing covariate data or a history of kidney transplantation at baseline. This research was conducted under UK Biobank application number 49978.

### Projection of KPMP-derived factor scores to UK Biobank proteomics

We applied the LASSO regression models trained on KPMP plasma SomaScan data to the UK Biobank Olink proteomics platform. Because the two platforms measure partially overlapping but distinct protein panels, we harmonized protein identifications based on protein names. Program scores were computed as a weighted linear combination of standardized plasma protein abundances. We computed signature scores with column-wise z-scaling of proteomic features, winsorization of weights at the 99th percentile of absolute weight magnitude, and L2-normalization of weights to stabilize scale across signatures. The resulting raw signature scores were then z-standardized across participants to yield comparable effect estimates per 1 SD increase.

### Cross-sectional association with baseline kidney function

We assessed the relationship between the projected factor scores and baseline eGFR using multivariable linear regression. Models were adjusted for age, sex, body mass index (BMI), smoking status (current/former vs. never), and baseline comorbidities including diabetes mellitus, hypertension, heart failure, and myocardial infarction, as well as use of renin-angiotensin system (RAS) inhibitors. We examined potential effect modification by baseline chronic kidney disease (CKD) status (eGFR < 60 mL/min/1.73m^2^) by fitting stratified models and testing for statistical interaction. Non-linearity of associations was assessed using restricted cubic splines with 3-4 degrees of freedom, with model comparisons performed using likelihood ratio tests and Akaike Information Criterion (AIC). For visualization, we generated predicted eGFR curves across the continuous range of factor scores using the spline models, holding covariates at their mean or reference values.

### Disease specificity assessment across cardiometabolic and inflammatory phenotypes

To evaluate the disease specificity of the acute and chronic kidney injury programs, we conducted a phenome-wide association study (PheWAS) examining the relationship between factor scores and baseline prevalence of 28 relevant cardiometabolic, inflammatory, and chronic conditions in the UK Biobank. This analysis aimed to determine whether the factor scores represent kidney-specific pathobiology or reflect broader systemic inflammatory or metabolic states.

We ascertained baseline disease status using a combination of hospital admission records, primary care data, and self-reported conditions at the time of baseline assessment. Conditions were classified using ICD-10 diagnosis codes and UK Biobank-specific field codes. We included the following disease categories: chronic kidney disease (CKD, defined both based on reporting and as eGFR < 60 mL/min/1.73m^2^), hypertension, diabetes mellitus (any type), heart failure with cardiomyopathy, myocardial infarction, ischemic heart disease, atherosclerosis, peripheral vascular disease, atrial fibrillation, stroke, hypothyroidism, hyperthyroidism, obesity, dyslipidemia, chronic obstructive pulmonary disease (COPD), asthma, interstitial lung disease, inflammatory bowel disease, liver disease, Parkinson’s disease, other neurodegenerative diseases, rheumatoid arthritis, gout, osteoarthritis (knee), systemic lupus erythematosus, osteoporosis, and benign prostatic hyperplasia.

Phenotypes with fewer than 20 prevalent cases were excluded from analysis to ensure adequate statistical power and model convergence. For each phenotype, we fitted multivariable logistic regression models to estimate the association with standardized factor scores (Factor 2 for acute injury, Factor 5 for chronic damage), adjusting for age, sex, body mass index, smoking status (current/former vs. never), and use of renin-angiotensin system inhibitors. Models were fitted independently for each factor. We report odds ratios (OR) per standard deviation increase in factor score with 95% confidence intervals and two-sided P-values. To facilitate comparison of effect sizes across conditions, we ranked phenotypes by the magnitude of association (absolute value of log OR) for each factor.

### Longitudinal association with incident kidney disease outcomes

To examine whether the projected factor scores predict future kidney disease, we conducted time-to-event analyses in participants with preserved kidney function at baseline (eGFR ≥ 60 mL/min/1.73m^2^). We defined two incident outcomes ascertained using ICD-10 codes: (1) acute kidney injury (AKI), defined as N17.x or N19; and (2) incident CKD, defined as N18.x. Follow-up time was calculated from the date of baseline assessment to the first occurrence of the outcome, death, loss to follow-up, or administrative censoring at 5 years, whichever occurred first.

Associations between projected program activity and incident outcomes were evaluated using multivariable Cox proportional hazards models adjusted for age, sex, BMI, smoking status, baseline diabetes, hypertension, heart failure, prior myocardial infarction, baseline eGFR, and RAS inhibitor use. We fit pre-specified piecewise Cox models by splitting follow-up time into clinically motivated early intervals for AKI (0–3, 3–6, 6–12, and > 12 months) and CKD (0-1 year, 1-3 years, 3-5 years, >5 years), allowing the estimated effect of program activity to vary across periods to explore the relevance of incident event timing in the association with each factor projection.

### Risk stratification by dual factor classification

We implemented a dual-factor classification based on median splits of the blood-derived acute and chronic scores, yielding four groups: reference (low/low), High Acute Only, High Chronic Only, and Double Hit (high/high). Median thresholds were chosen a priori to provide a simple, distribution-agnostic stratification that (i) avoids assumptions about linearity or calibration of projected scores, (ii) yields balanced group sizes that stabilize estimation and enable direct comparison of discordant phenotypes, and (iii) produces an easily communicable risk taxonomy aligned with the conceptual biology of acute injury versus chronic damage programs. Group differences in incident AKI and CKD were quantified using adjusted Cox models with the reference group as the reference category.

All analyses were performed in R version 4.3.0. Linear models were fitted using the lm() function, Cox models using the survival package, restricted cubic splines using the splines package, and marginal means estimated using the emmeans package. Survival curves were visualized using the ggsurvfit package. Statistical significance was defined as a two-sided P < 0.05.

## Code and data availability

snRNA and scRNA data from the Human Kidney Atlas v2 were downloaded from cellxgene (https://cellxgene.cziscience.com/collections/9c9d04c4-8899-417f-bb6f-6107dcadf14f).

SomaScan urine and plasma data as well as pen access clinical information for KPMP participants are available at atlas.kpmp.org, whereas controlled access data are not publicly available to safeguard participant privacy and can be requested. UK biobank data were accessed under project number 49978 and are available upon application (see https://www.ukbiobank.ac.uk). The code used to generate the results will be made public upon journal acceptance.

## Supporting information

KPMP Consortium Members

## Data Availability

snRNA and scRNA data from the Human Kidney Atlas v2 were downloaded from cellxgene (https://cellxgene.cziscience.com/collections/9c9d04c4-8899-417f-bb6f-6107dcadf14f). SomaScan urine and plasma data as well as pen access clinical information for KPMP participants are available at atlas.kpmp.org, whereas controlled access data are not publicly available to safeguard participant privacy and can be requested. UK biobank data were accessed under project number 49978 and are available upon application (see https://www.ukbiobank.ac.uk). The code used to generate the results will be made public upon journal acceptance.

https://atlas.kpmp.org

## Acknowledgements

The Kidney Precision Medicine Project (KPMP) is supported by the National Institute of Diabetes and Digestive and Kidney Diseases (NIDDK) through the following grants: U01DK133081, U01DK133091, U01DK133092, U01DK133093, U01DK133095, U01DK133097, U01DK114866, U01DK114908, U01DK133090, U01DK133113, U01DK133766, U01DK133768, U01DK114907, U01DK114920, U01DK114923, U01DK114933, U24DK114886, UH3DK114926, UH3DK114861, UH3DK114915, and UH3DK114937. Research reported in this publication was partly supported by the Office Of The Director, National Institutes Of Health under Award Number U54DK134301 (HuBMAP consortium, S.J.). We gratefully acknowledge the essential contributions of our patient participants and the support of the American public through their tax dollars. The content is solely the responsibility of the authors and does not necessarily represent the official views of the National Institutes of Health.

The authors acknowledge support by the state of Baden-Württemberg through bwHPC and the German Research Foundation (DFG) through grant INST 35/1597-1 FUGG, as well as the storage service SDS@hd supported by the Ministry of Science, Research and the Arts Baden-Württemberg (MWK) and the German Research Foundation (DFG) through grant INST 35/1803-1 FUGG and INST 35/1804-1 LAGG.

The authors acknowledge the University of Michigan Medical School Central Biorepository (RRID:SCR_026845) for providing biospecimen storage, management, and distribution services in support of the research reported in this publication/grant application/presentation.

Part of this research has been conducted using the UK Biobank Resource under Application Number 49978, and uses data provided by patients and collected by the NHS as part of their care and support. This research used data assets made available by National Safe Haven as part of the Data and Connectivity National Core Study, led by Health Data Research UK in partnership with the Office for National Statistics and funded by UK Research and Innovation (grant ref MC_PC_20058).

## Authors contributions

RF, CB, EP, JH, JSR, JT, MK, MTE, RORF, SAGO conceived and planned the study. FPW, JRS, KK, LCH, MAV, MLC, PMP, SER contributed participant samples as part of KPMP recruitment sites. ASB, AZR, CEA, DD, FPW, KK, LB, LCH, MAV, MK, PMP, R. Murugan, RF, SER contributed to the analysis of clinical variables and/or the adjudication of cases. AZR, CEA, DD, JBH, LB, LCH contributed to the scoring and/or analysis of the pathology TIV descriptors. DD, FMA, JBH, JFOT, LP, MK, MTE, PB, RM, SE, SJ, VN, YJC contributed to the generation, processing, or annotation of the single cell/nuclei data. RF performed the analyses with MCFA and the downstream results including the association with participant metadata, cell type and cell state abundances, transcription factor regulation and how it relates to urine and plasma SomaScan data. SAGO performed the analysis on the UK biobank Olink data and its link to eGFR and longitudinal outcome. CB, DF, EP, JT, MP, RF, RORF contributed to MCFA analysis. CB, JT, RF, RORF, SAGO, SE, VN, WJ contributed to the analysis of the KPMP urine and plasma SomScan biomarker data. CB, DF, EP, JH, JSR, JT, MK, MP, MTE, OGT, RF, RORF, RSGS, SAGO, WM, XC, ZZ interpreted the findings. JSR supervised the study, and MTE, MK supervised the clinical and translational aspects of the study. RF, SAGO designed and generated the figures. RF wrote the first draft of the manuscript. All authors reviewed and approved the manuscript.

## List of Supplementary Materials

- List of contributors for the KPMP study
- Supplementary Tables

- Supplementary Table S1: scRNA cohort characteristics
- Supplementary Table S2: SomaScan data characteristics and models
- Supplementary Table S3: Baseline Characteristics of UK Biobank Participants by CKD status
- Supplementary Table S4 Adjusted association between predicted program scores and eGFR
- Supplementary Table S5: Hazard ratio of incident AKI based on acute or chronic plasma signature in UK biobank
- Supplementary Table S6: Hazard ratio of incident CKD based on acute or chronic plasma signature in UK biobank
- Supplementary Table S7: Hazard ratio of incident AKI or CKD based on combination of acute and chronic plasma signature in UK biobank
- Supplementary Methods and Figures

### Supplementary Methods

#### MCFA model including healthy reference samples

We apply MCFA on all samples included in the snRNA and scRNA data of the Human Kidney Atlas v2^23^, including healthy reference samples. Preprocessing steps, including filtering of samples and cell types, and MCFA model fit are done as described in the main methods.

#### Projection of external dataset

We use the single cell data from a cohort with healthy reference samples obtained through percutaneous biopsy^101^, and project these samples by matrix multiplication into the factor space built by HKA v2 scRNA data. This dataset comprises samples from Impact of Metabolic Surgery on Pancreatic, Renal and Cardiovascular Health in Youth with Type 2 Diabetes (IMPROVE-T2D, NCT03620773), Renal Hemodynamics, Energetics and Insulin Resistance in Youth Onset Type 2 Diabetes (Renal HEIR, NCT03584217) Control of Renal Oxygen Consumption, Mitochondrial Dysfunction, and Insulin Resistance (CROCODILE, NCT04074668) studies. For the projection, we take the pseudo-inverse of the loadings matrix (from KPMP scRNA) and multiply it by the normalized, log1p-transformed and then centered multicellular gene expression matrix of the external dataset to obtain the factor scores for the projected samples. In that process, we manually matched cell types between the

#### Deriving of ischemia signature

To identify a consensus ischemia signature across omics and cell types, we used a rank-based approach, where gene loadings for each cell type were ranked by value, and then averaged across snRNA/scRNA. Assuming that ischemia affects all cell types similarly, we then averaged the ranks across cell types to obtain a consensus ranking, further selecting genes that were measured consistently across at least 3 cell types. The top 50 genes with the highest average rank were selected as ischemia signature genes.

To show that factor scores yield a similar signature to the one from Menon et al.^28^, we show that their gene signature is enriched in the factor loadings from snRNA using ORA, and that the genes from their signature are highly ranked in our consensus ranking using GSEA’s running score, both using the decoupler-py package^92^.

#### Ischemia signature in bulk datasets

We selected previously published bulk RNA and proteome studies that use control samples obtained through nephrectomy^102–107^ and that report differentially expressed gene probes, genes and/or differentially regulated proteins. Given that most studies only report differentially expressed probes/genes/proteins, we used Fisher’s exact test – implemented in scipy^108^ – to assess the enrichment of our ischemia signature in their reported downregulated probes/genes/proteins. The studies used different analytical approaches and thresholds to define differential expression, and we used the reported p-values and thresholds from each study without re-analyzing the data. When available, we use the reported feature space used by the study as background for Fisher’s exact test, otherwise we use the feature space size reported by the study or measured by the platform (e.g., microarray).

## Supplementary Figures and Tables

**Supplementary Figure 1:**
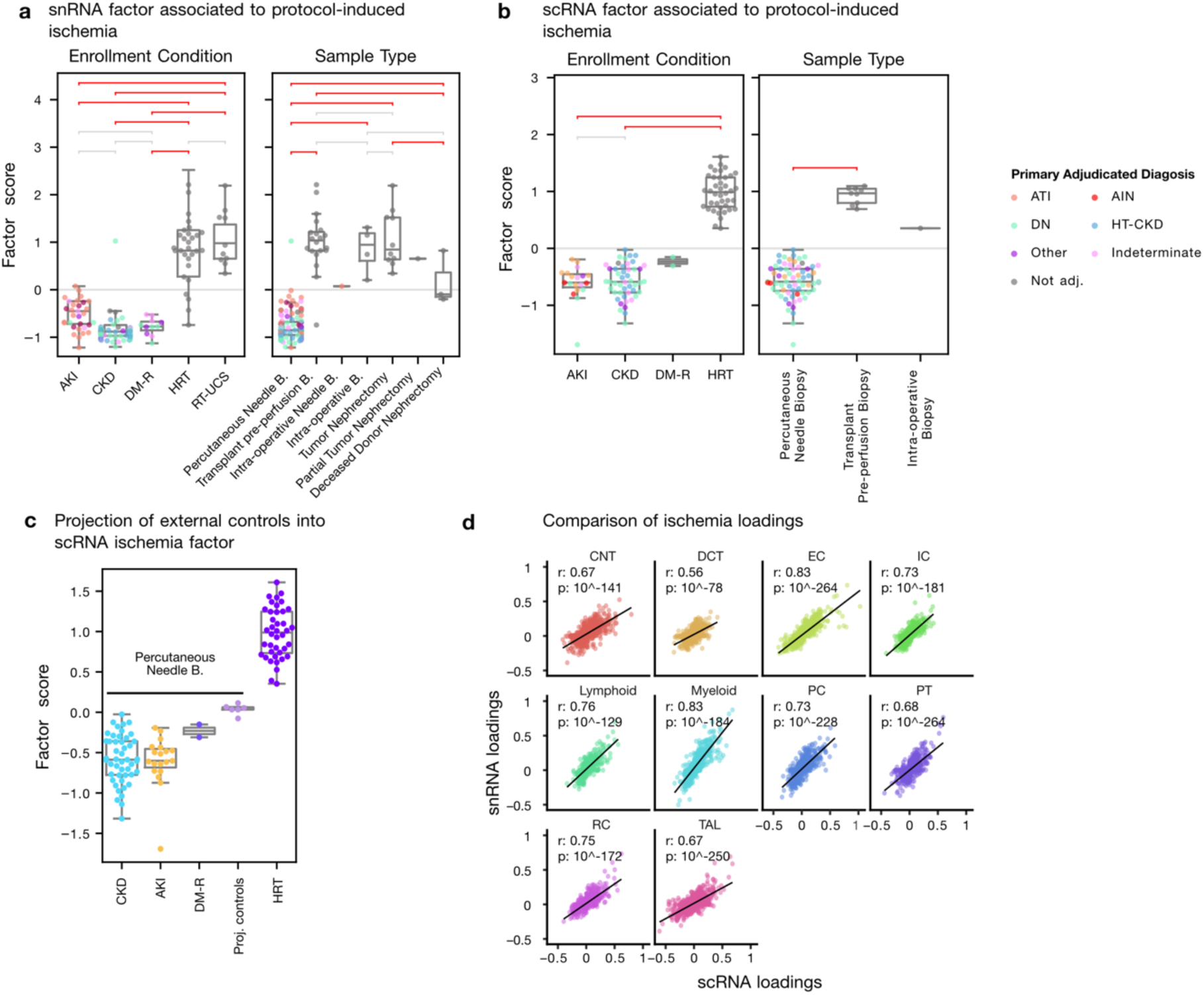
Factors capture ischemic signature due to procurement. a./b. Technical factors in snRNA and scRNA separate samples based on enrollment conditions, and is confounded with sample procurement method c. Projection of healthy reference samples from the CROCODILE study (percutaneous biopsies) onto scRNA factor capturing ischemia d. Correlation between factor loadings from snRNA and scRNA for factors relating to ischemia signature

**Supplementary Figure 2:**
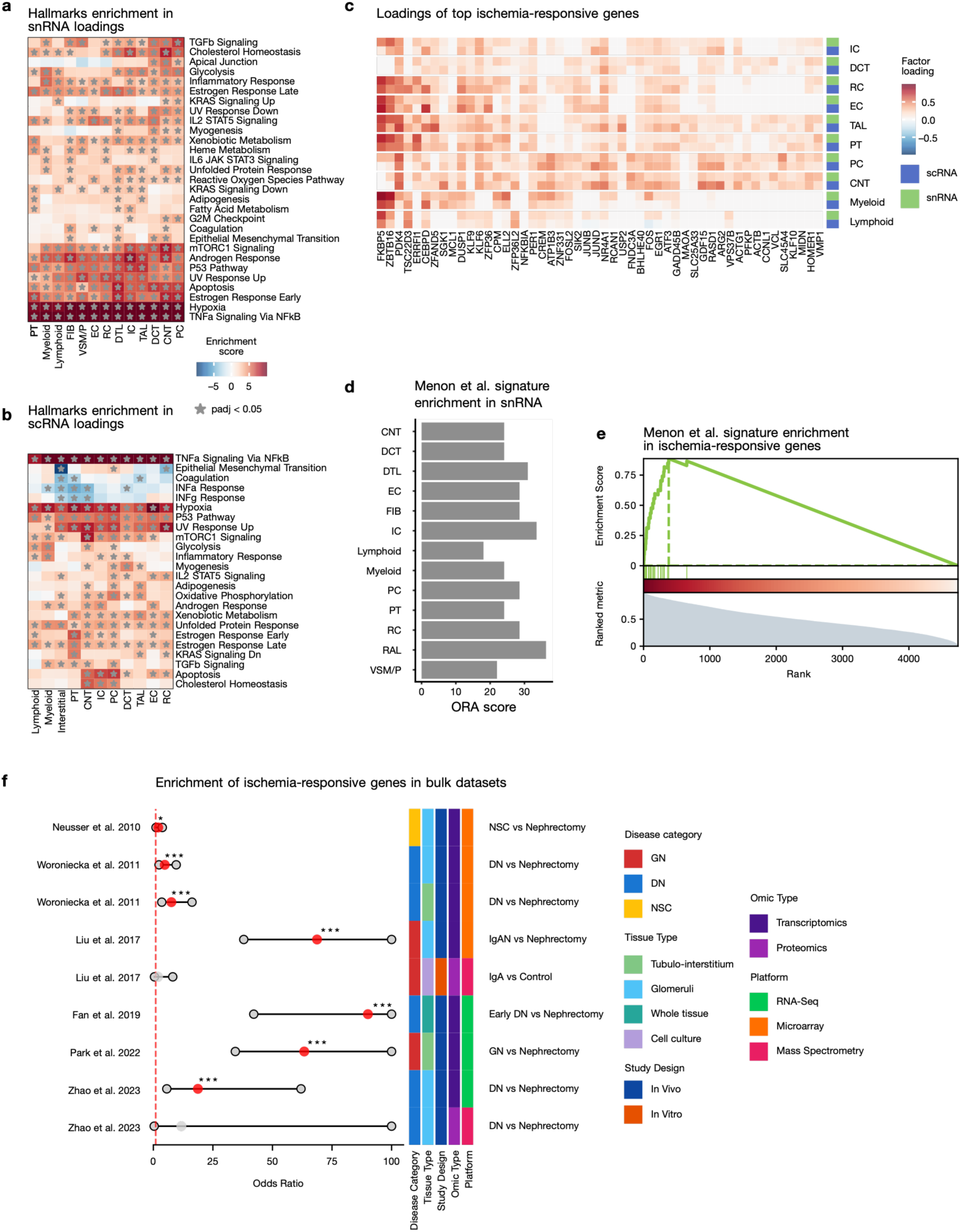
Ischemic signature is captured by factors in snRNA and scRNA. a./b. MSigDB hallmark enrichment in factor shows increased inflammatory signaling, hypoxia and apoptosis in snRNA and scRNA respectively c. Ischemia-induced gene signature based on consensus between snRNA and scRNA factor loadings across cell types d. Enrichment of gene signature from Menon et al.^28^ (derived from scRNA) in the factor loadings of snRNA e. GSEA running score in the ischemia ranking shown in c. of the signature from Menon et al. f. Odds ratio of finding ischemia signature genes (from c) in genes more highly expressed in controls from bulk RNA and protein studies which use nephrectomy samples as control.

**Supplementary Table S1:**
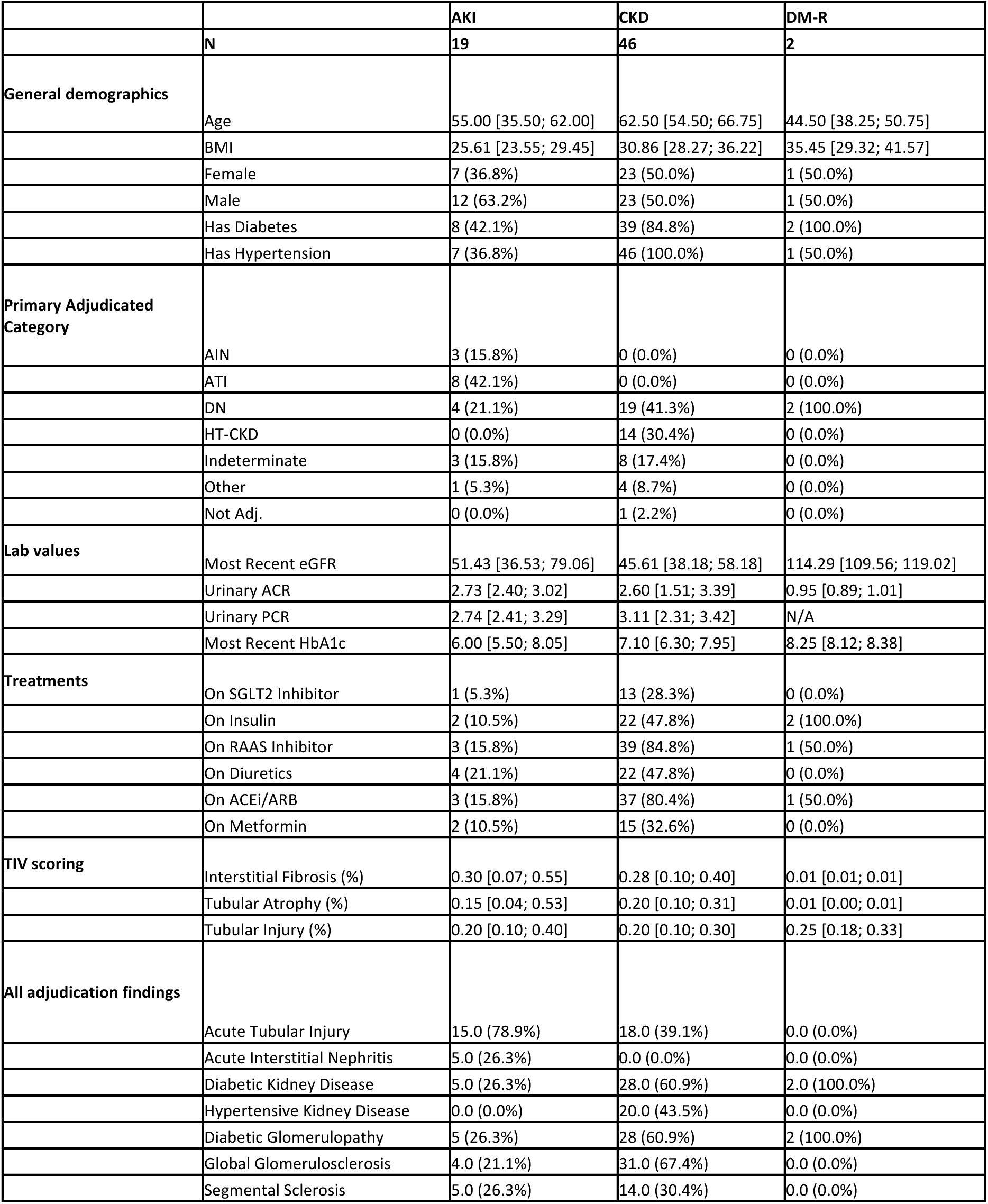
scRNA cohort characteristics.

**Supplementary Figure 3:**
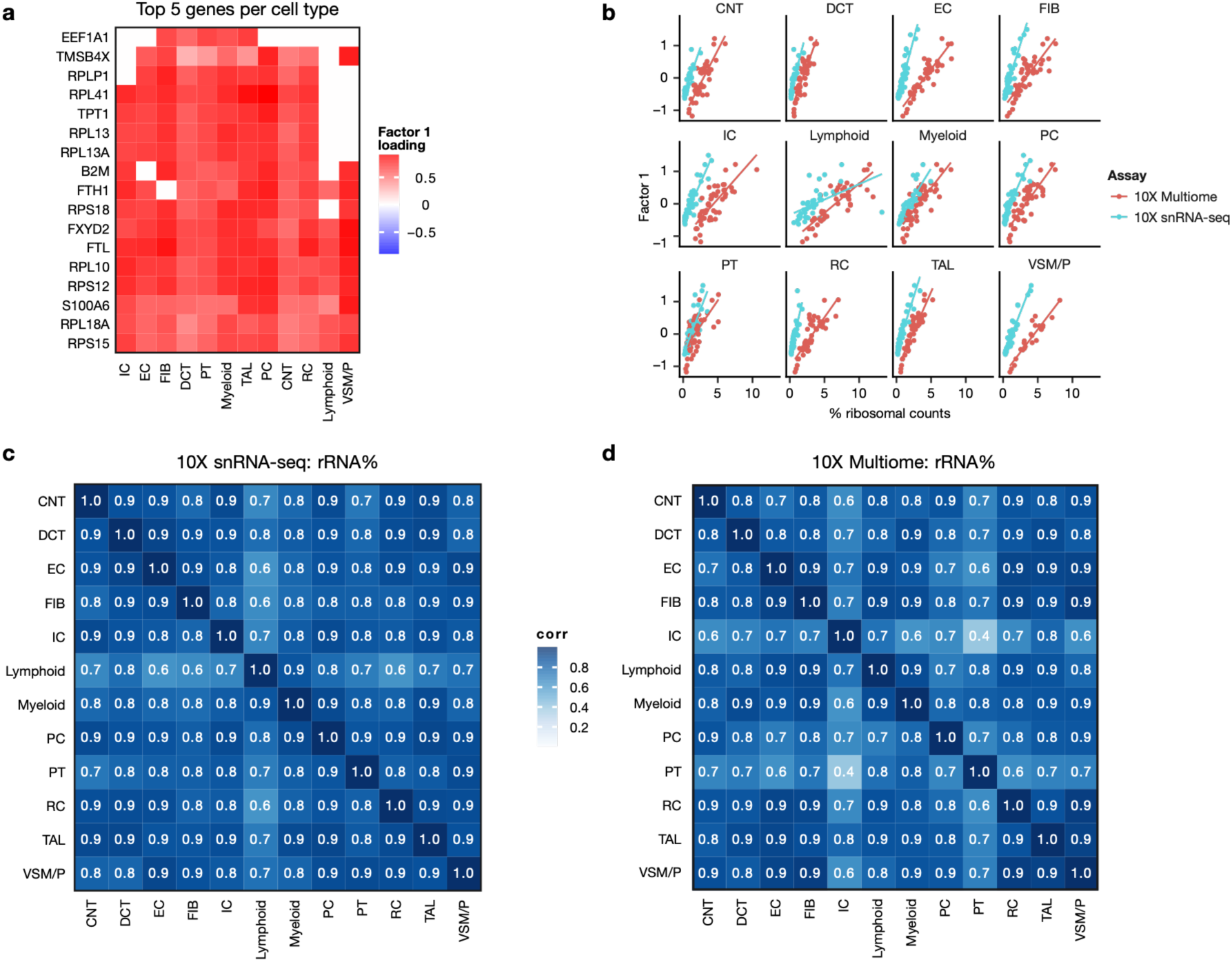
Factor 1 explains ribosomal content differences between samples in snRNA. a. Top 5 Factor 1 gene loadings per cell type from snRNA. b. snRNA factor 1 scores relative to percentage of ribosomal counts c./d. Pearson correlation of ribosomal content across cell types pseudobulks in 10X snRNA-seq and 10X Multiome samples

**Supplementary Figure 4:**
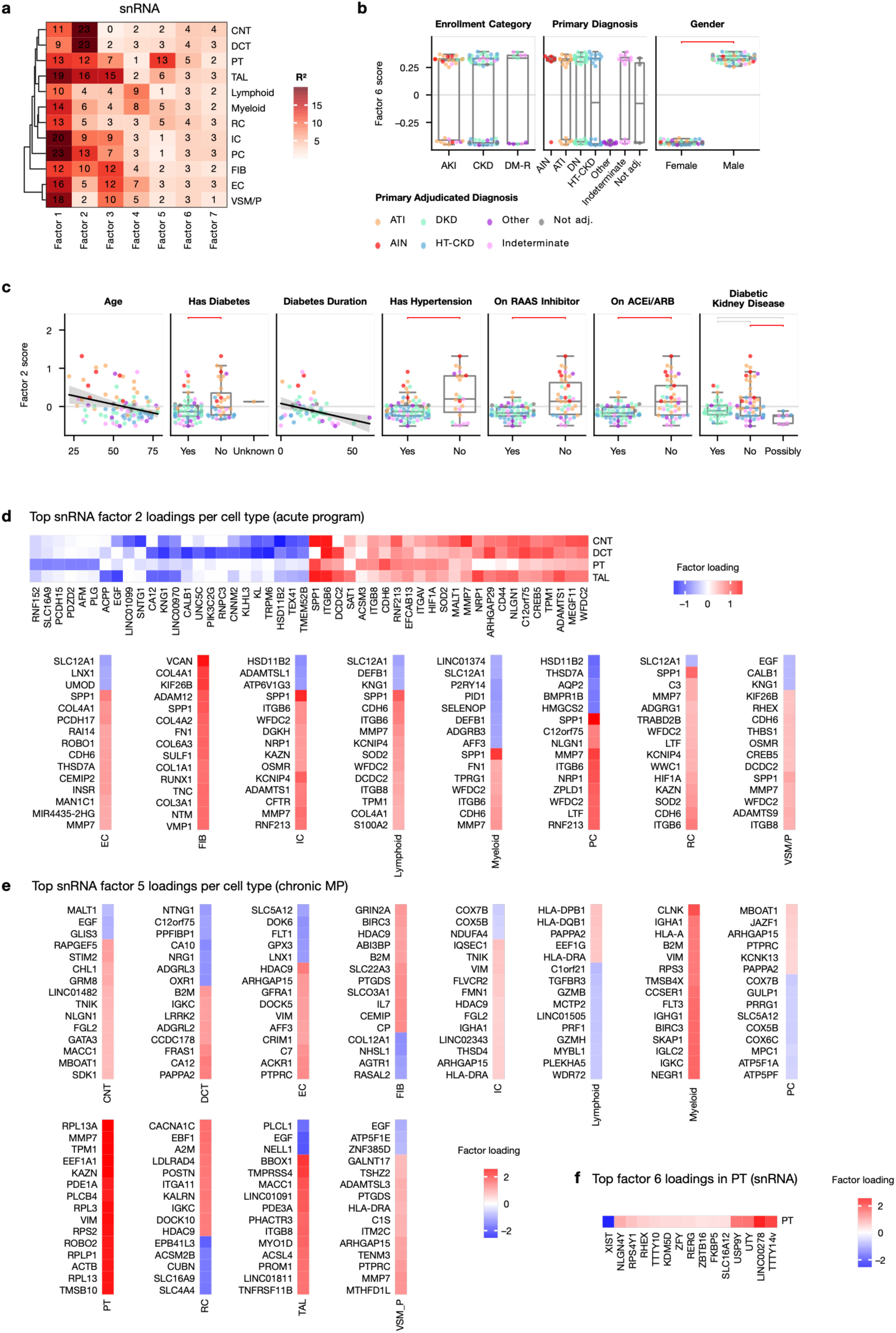
a. Gene expression variance explained by MCFA model in snRNA per cell type b. Factor 6 captures sex differences c. additional metadata associations with Factor 2 of snRNA. Horizontal brackets show Tukey’s HSD tests in red if p < 0.05 and grey if not significant. CBD: Cannot be Determined d. Top gene loadings in factor 2 (acute program) per cell type e. Top loadings in factor 5 (chronic program) per cell type f. Top loadings in factor 6 in PT

**Supplementary Figure 5:**
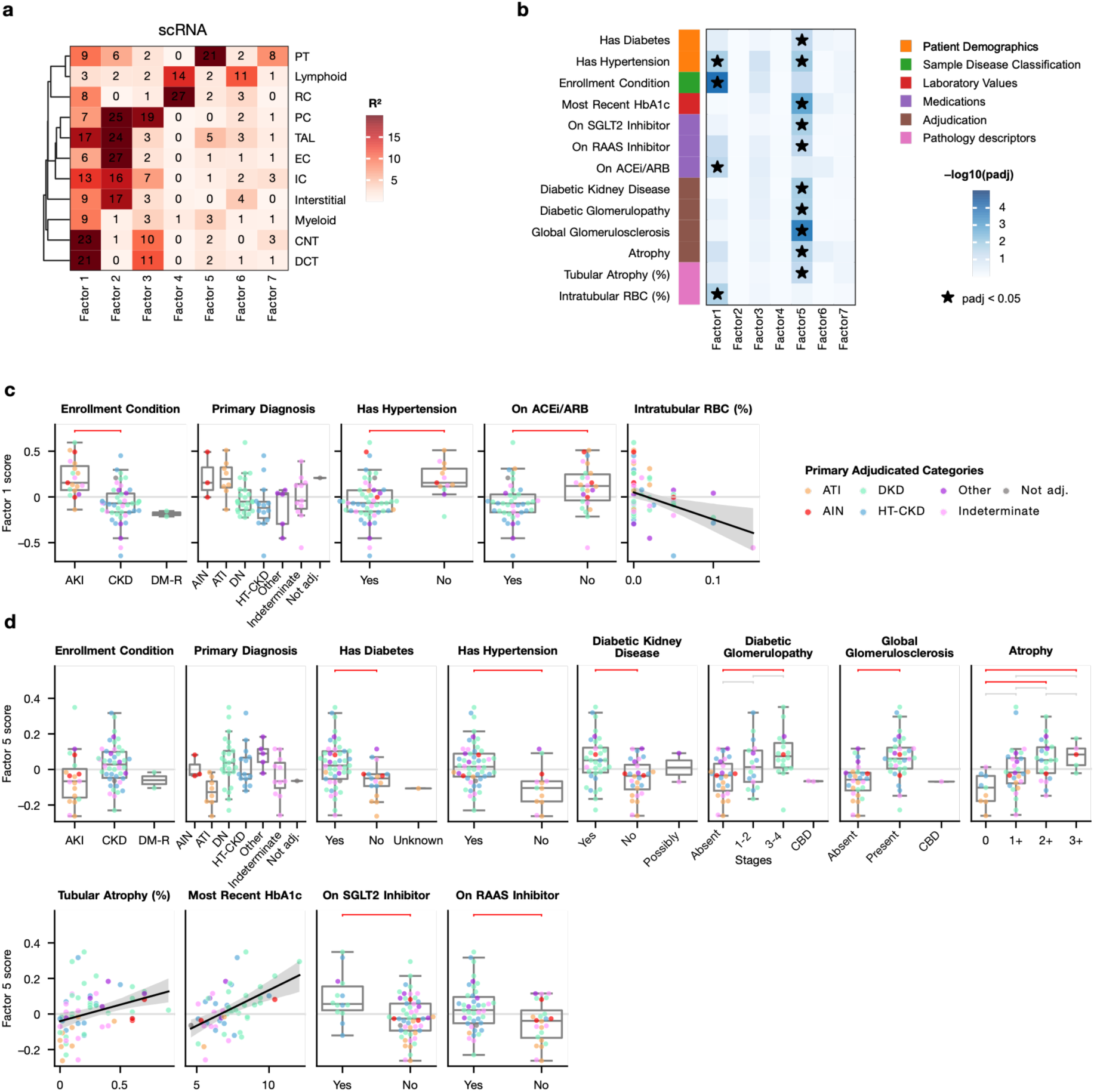
a. Gene expression variance explained by MCFA model in scRNA per cell type b. Statistical significance of the association between sample metadata and the MCFA factor scores in scRNA; * marks significant associations (adj. p. < 0.05). c./d. Metadata associations with Factor 1 and 5 respectively (acute, chronic) of scRNA. Horizontal brackets show Tukey’s HSD tests in red if p < 0.05 and grey if not significant. CBD: Cannot be Determined

**Supplementary Figure 6:**
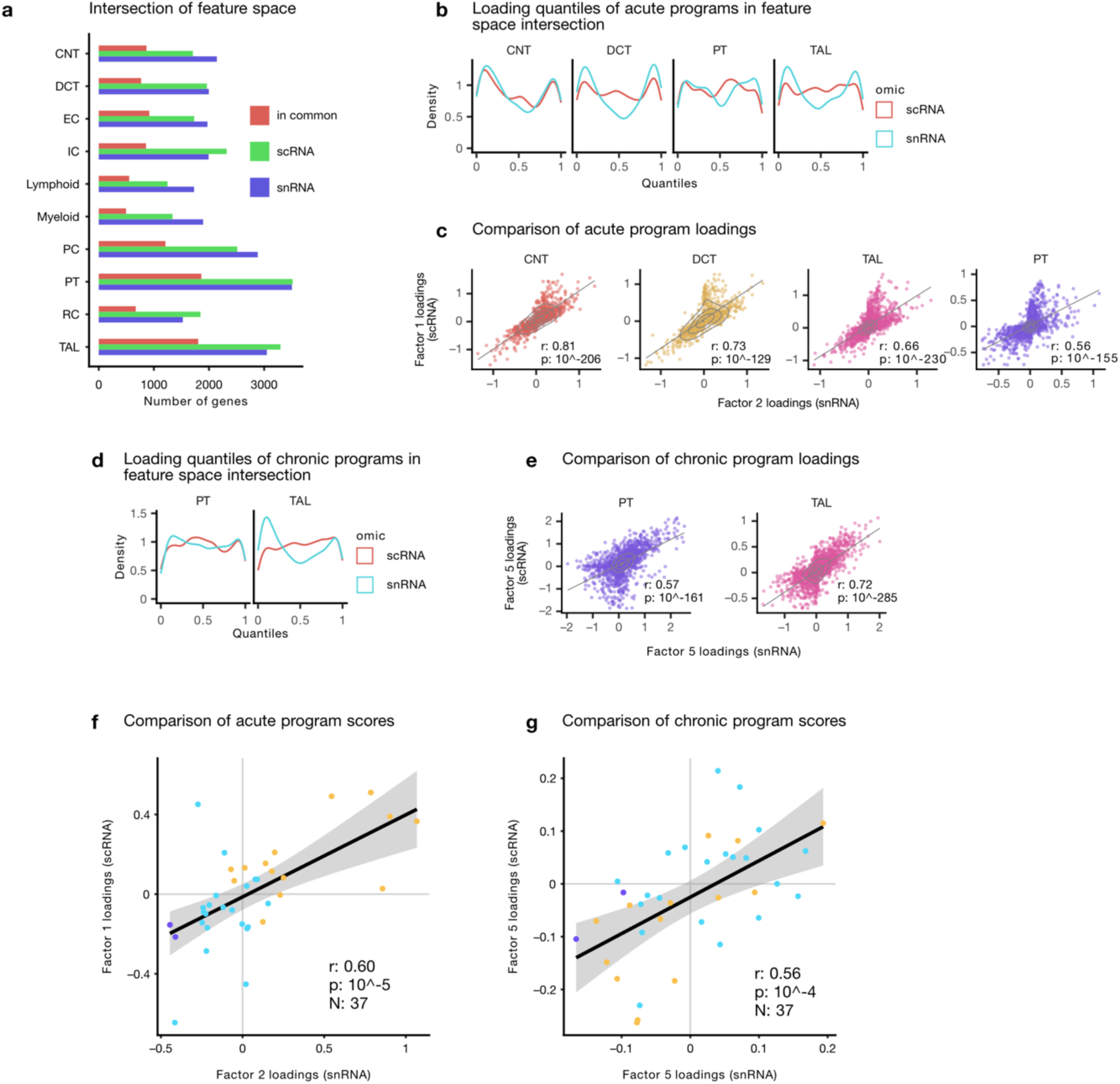
Comparison of factors from snRNA and scRNA. a. Gene overlap between snRNA and scRNA MCFA models, b. quantiles of gene loadings for genes included in the intersection of genes between snRNA factor 2 and scRNA factor 1 (acute programs) for best explained cell types, c. Pearson correlation of the gene loadings between snRNA factor 2 and scRNA factor 1 (acute programs), d. as in b. for factors 5 (chronic programs) from snRNA/scRNA, e. as in c. for factors 5 (chronic programs) from snRNA/scRNA, f. Pearson correlation of snRNA factor 2 and scRNA factor 1 scores (acute programs) for patients with both snRNA and scRNA data, g. as in f. for factors 5 (chronic programs) from snRNA/scRNA

**Supplementary Figure 7:**
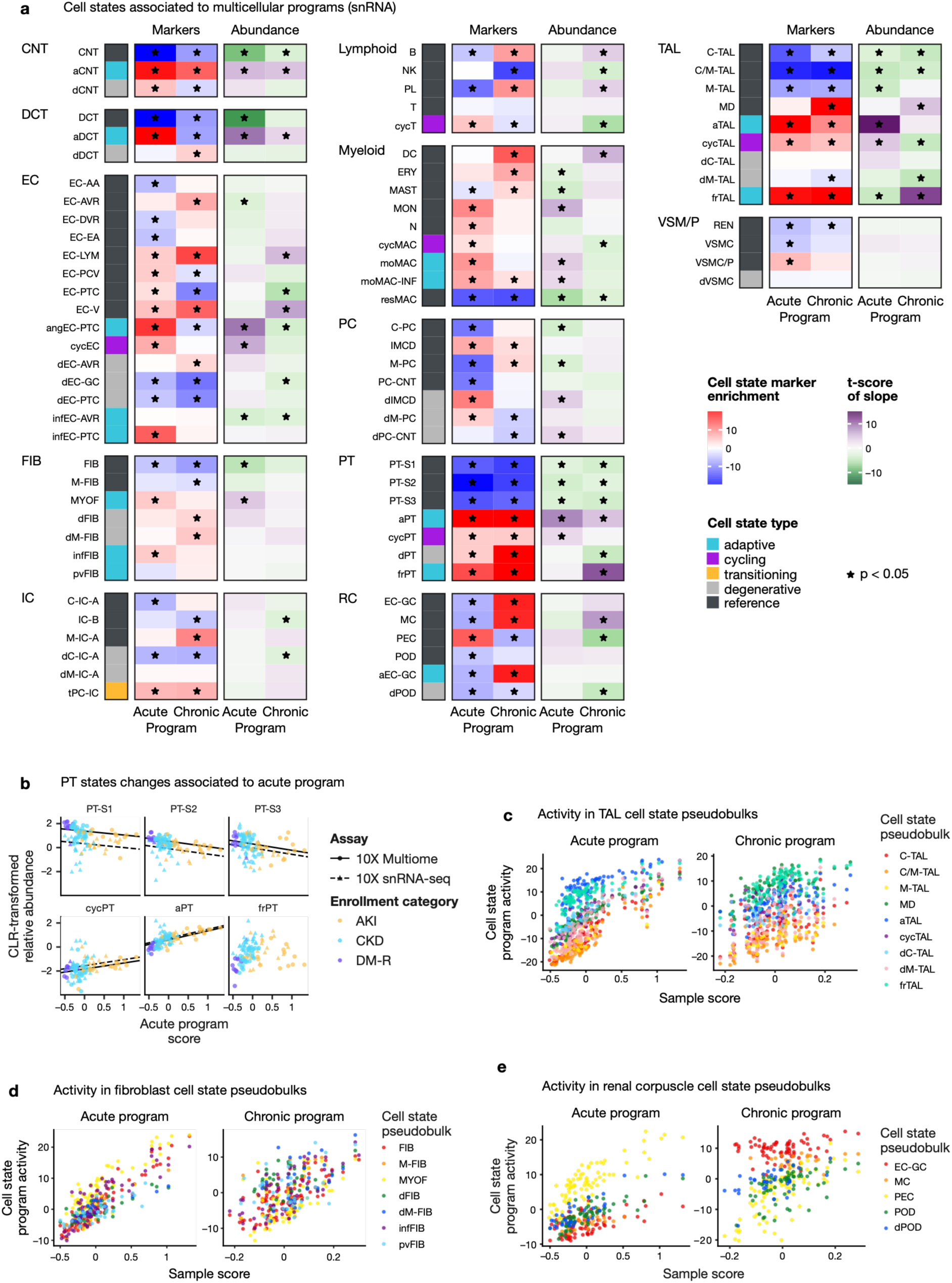
Compositional and molecular shifts captured by acute and chronic programs in snRNA. a. Cell state marker enrichment scores (red-blue heatmaps) in program loadings and t-scores of the slopes associating (e.g., in b) within-cell type relative cell state abundance to sample program scores (purple-green heatmaps) for major cell types in snRNA. * marks enrichments or slopes with padj < 0.05 b. CLR-transformed cell state proportion within PT relative to the acute sample program score. Regression lines show slope of the associated mixed-effect model, c-e acute and chronic program activity in PT, TAL and renal corpuscle cell state-level pseudobulks by sample program score in snRNA

**Supplementary Figure 8:**
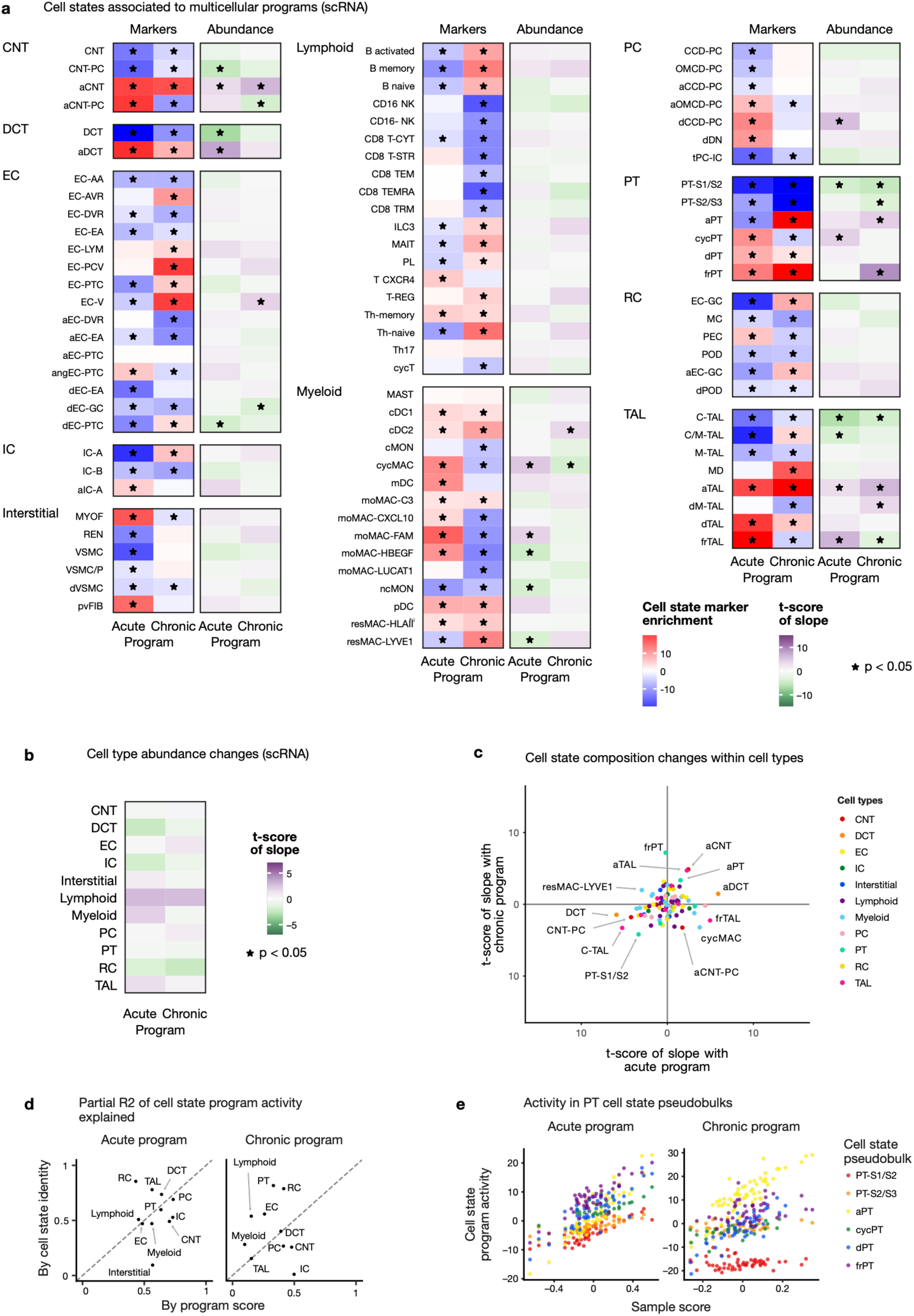
Compositional and molecular shifts captured by acute and chronic programs in scRNA. a. Cell state marker enrichment scores (red-blue heatmaps) in program loadings and t-scores of the slopes associating within-cell type relative cell state abundance to sample program scores (purple-green heatmaps) for major cell types in scRNA. * marks enrichments or slopes with padj < 0.05 b. Associations of acute and chronic program scores to overall shifts in relative cell type abundance in scRNA * mark significant associations (adj. p. < 0.05) c. t-scores of the slopes from mixed-effect model associating sample level acute (x-axis) and chronic (y-axis) program scores with relative cell state abundance within the respective cell types d. Partial R2 contribution of cell state identity and sample program score to the prediction of program activity in cell state pseudobulks in scRNA (e.g., show in e) e. Acute and chronic program activity in PT cell state-level pseudobulks by sample program score in scRNA

**Supplementary Figure 9:**
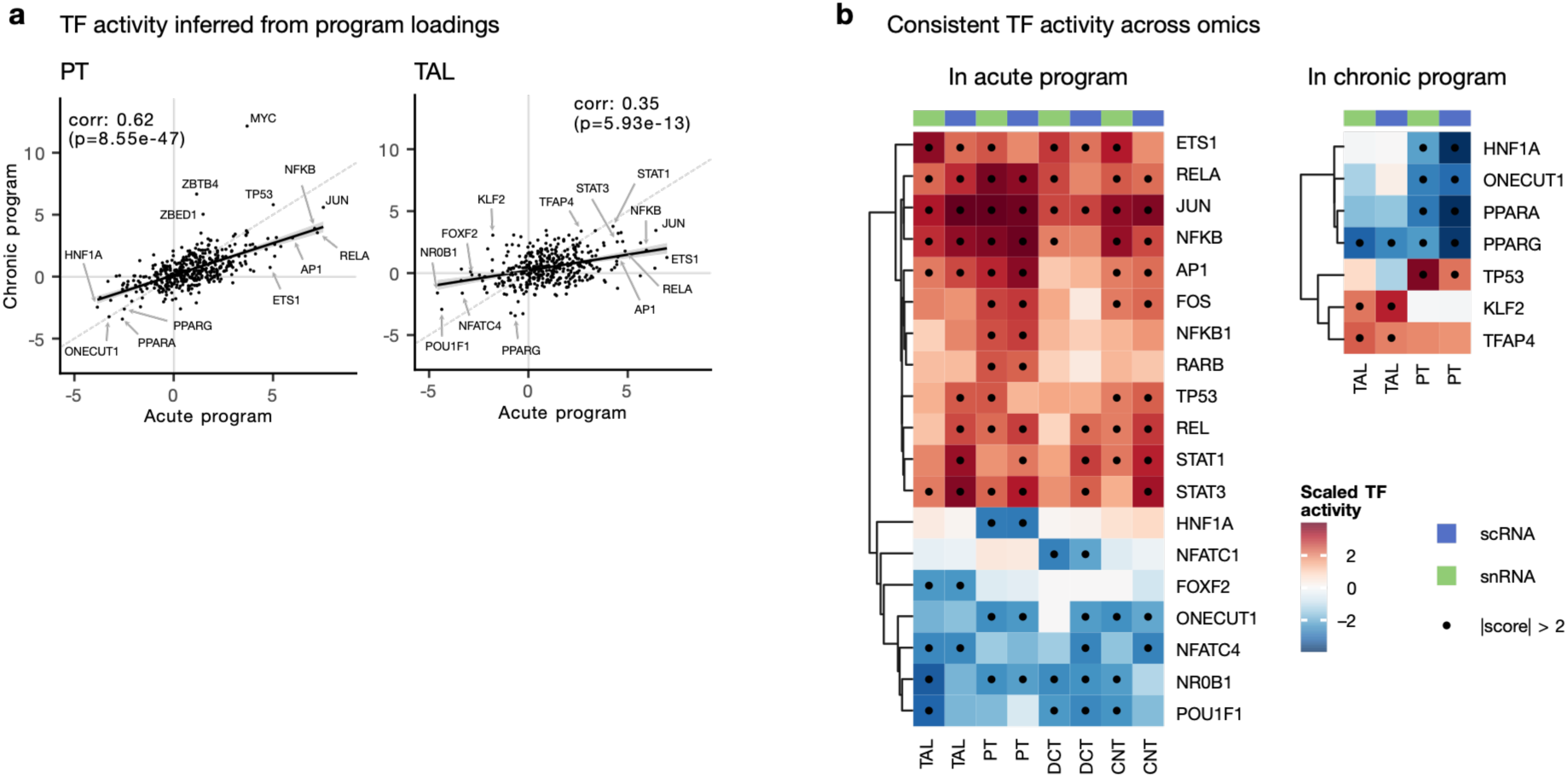
a. Correlation between transcription factor activities in the loadings of the acute (left) and chronic (right) programs for PT and TAL b. Top most deregulated transcription factors (absolute z-standardized activity > 2) in common between snRNA and scRNA for the acute (left) and chronic (right) program

**Supplementary Figure 10:**
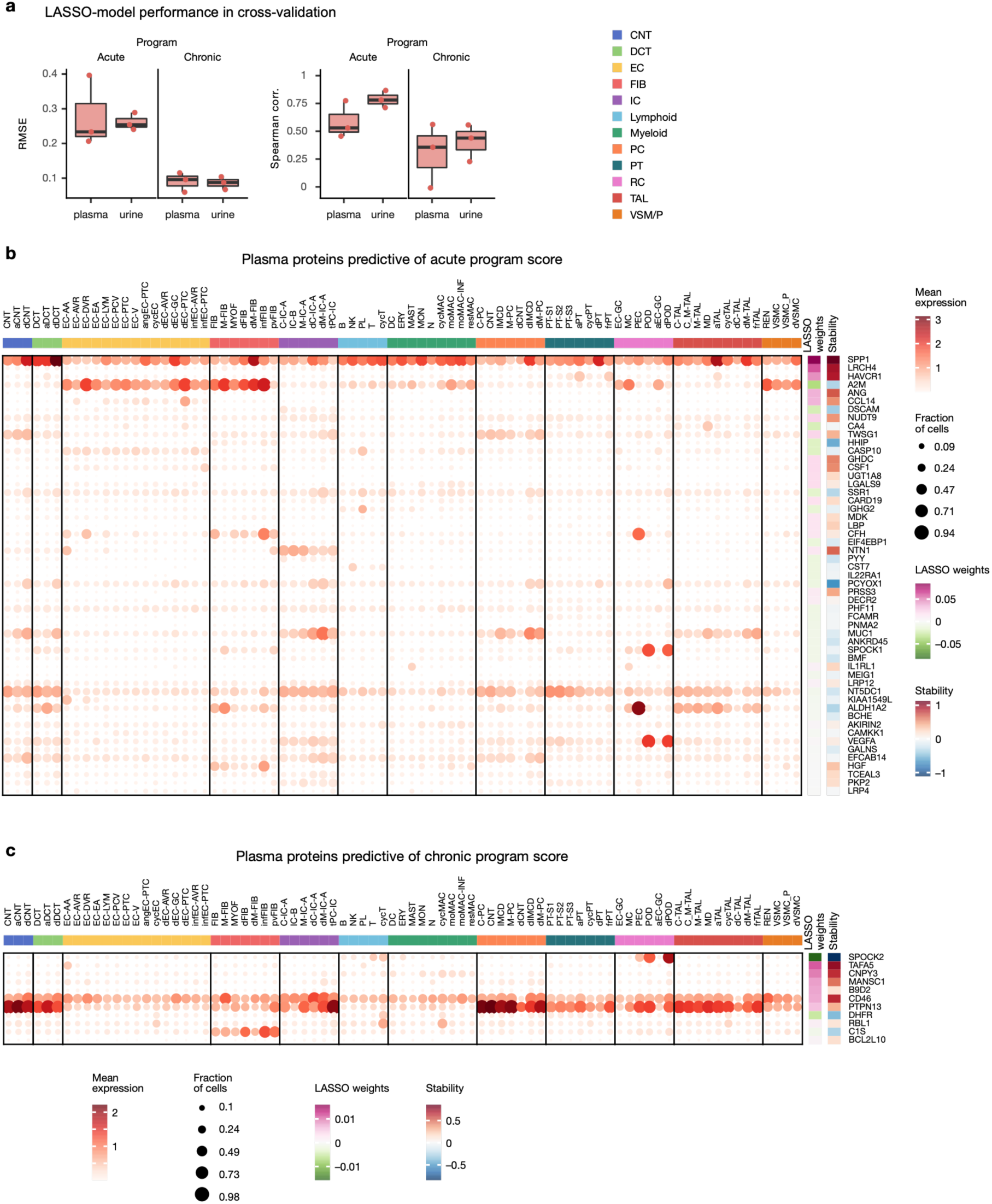
Cell type and cell state specificity of expression of plasma proteins predictive of acute and chronic program scores. a. LASSO-model performance in cross-validation in terms of root mean-square error (RMSE) and spearman correlation to predict acute/chronic program scores of samples from plasma or urine somascan proteins b./c. gene expression of plasma proteins that are cell type or cell state markers (dotplot), which were selected by the LASSO model to predict the acute and chronic program scores, respectively. The LASSO weights are in SD units for each protein. The stability is based on the mean/sd LASSO weight in bootstrap resampling times the inclusion rate (higher absolute value shows better numerical stability)

**Supplementary Table S2:**
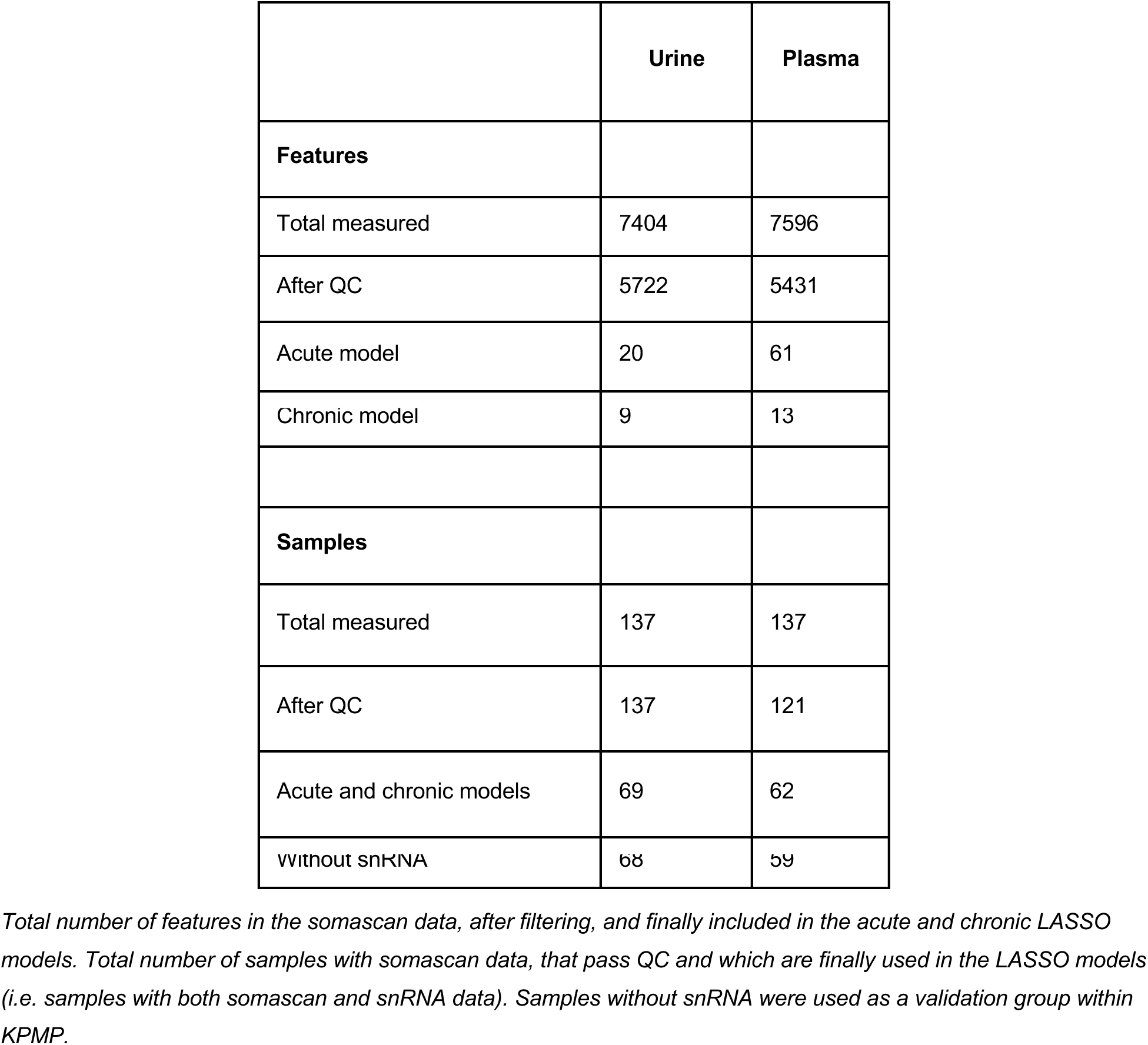
Somascan data characteristics and models.

**Supplementary Figure 11:**
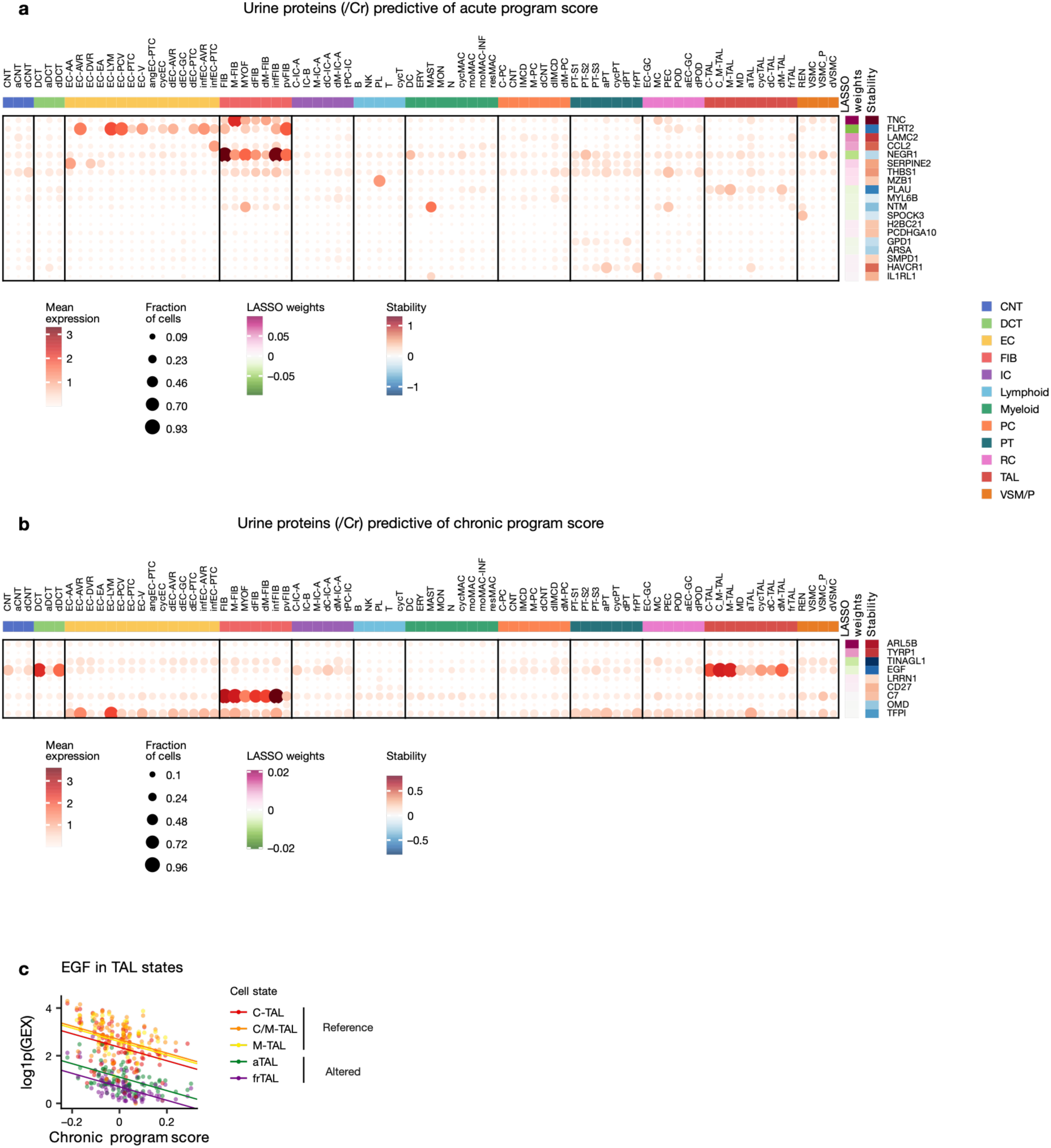
Cell type and cell state specificity of expression of urine proteins predictive of acute and chronic program scores. a./b. Gene expression of urinary proteins (normalized against urinary creatinine) that are cell type or cell state markers (dotplot), which were selected by the LASSO model to predict the acute and chronic program scores, respectively. The LASSO weights are in SD units for each protein. The stability metric is the mean/sd LASSO weight in bootstrap resampling times the inclusion rate (higher absolute value shows better numerical stability) c. EGF expression in TAL cell state pseudobulks relative to chronic program score. Slopes show the relation between gene expression and chronic program score with an intercept for each cell state, as determined by a mixed effect linear model.

**Supplementary Figure 12:**
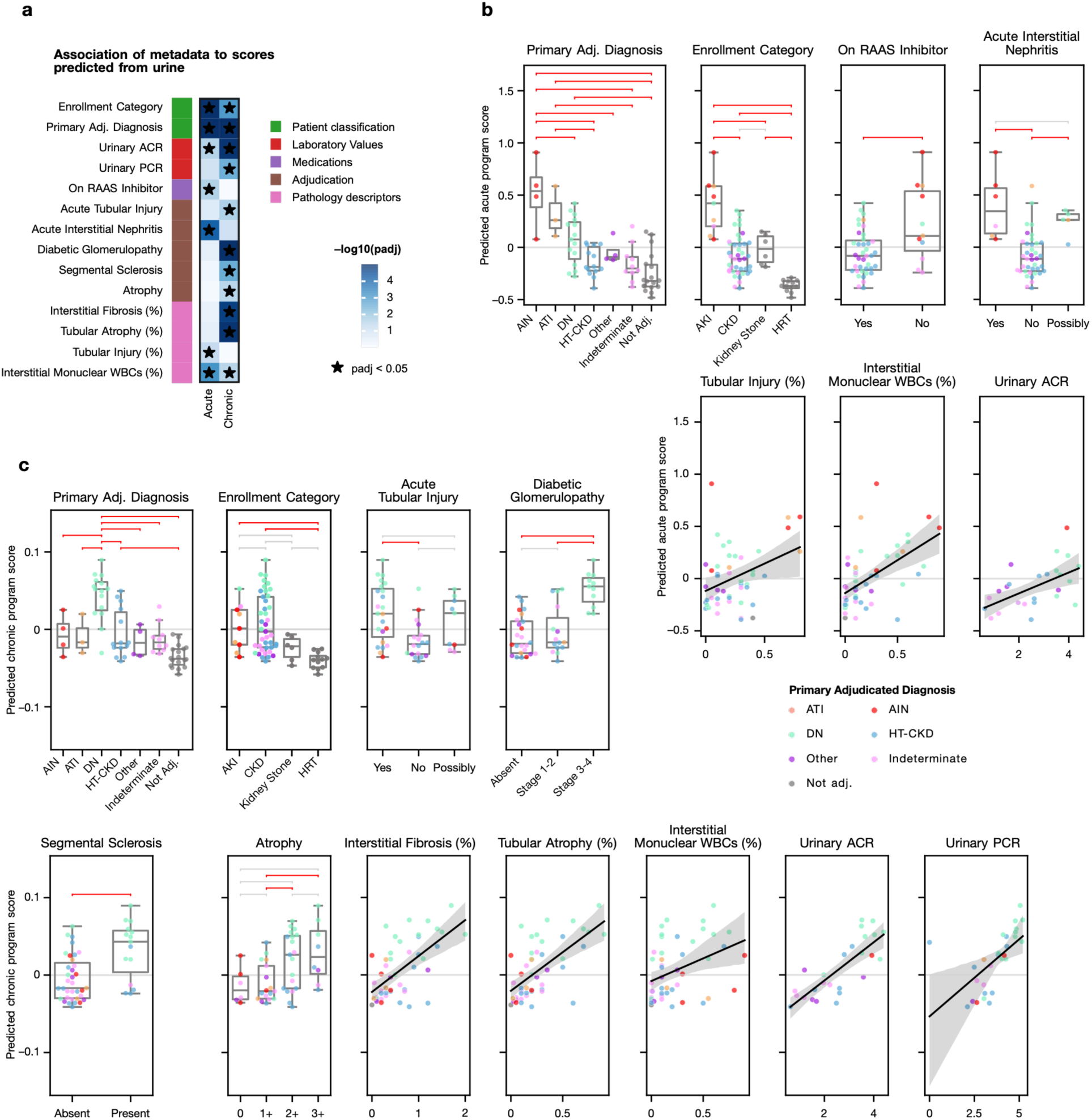
Association of acute and chronic scores predicted from urinary protein abundances. a. Significant associations between predicted scores and patient metadata. Adjusted p-values are shown based on analysis of variance or linear regression for categorical and continuous variables respectively. * marks significant associations (adj. p. < 0.05) b./c. Sample metadata associated with predicted acute and chronic scores. Horizontal brackets show Tukey’s HSD tests in red if p < 0.05 and grey if not significant. CBD: cannot be determined.

**Supplementary Figure 13:**
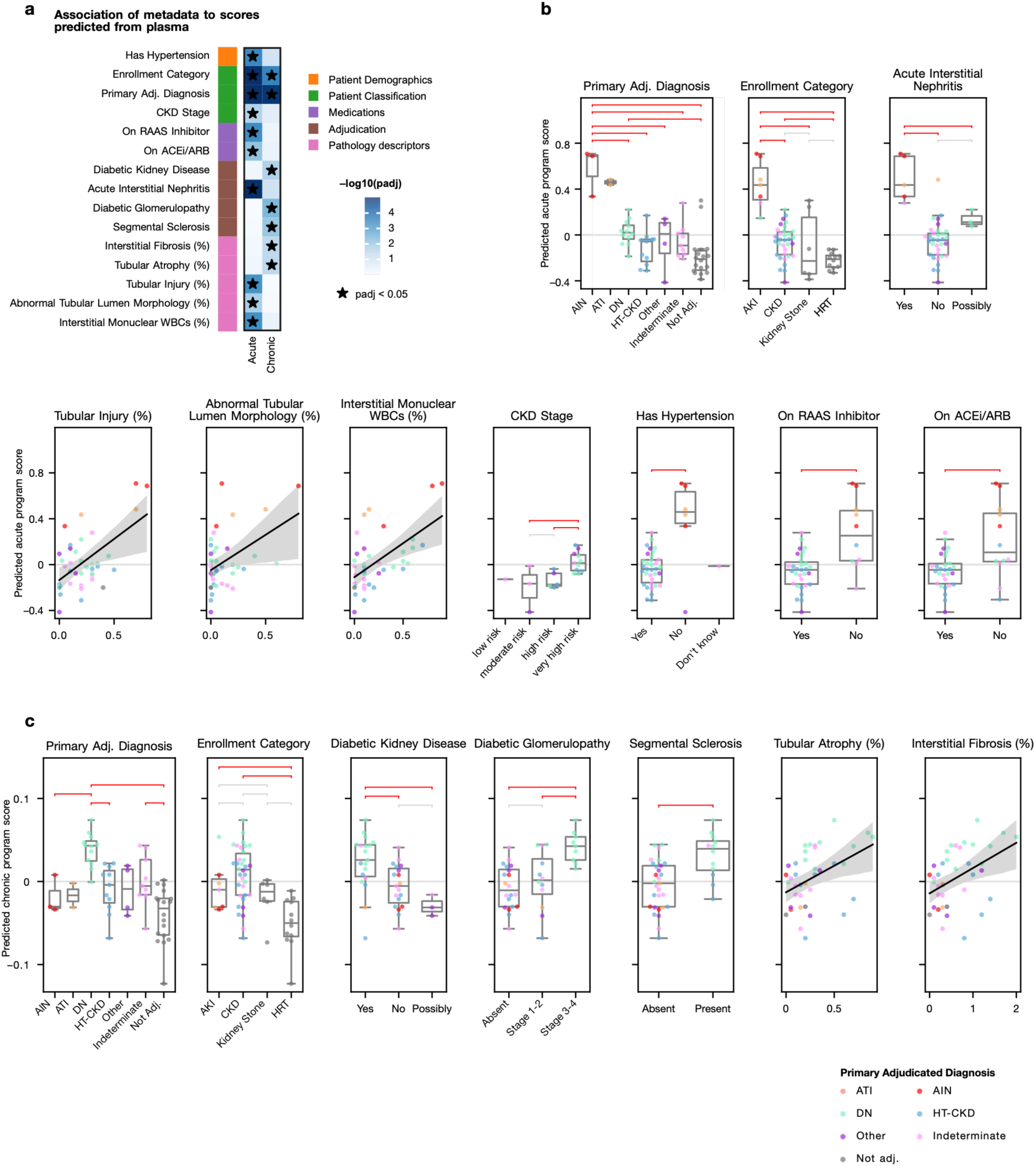
Association of acute and chronic scores predicted from plasma protein abundances. a. Significant associations between predicted scores and patient metadata. Adjusted p-values are shown based on analysis of variance or linear regression for categorical and continuous variables respectively. * marks significant associations (adj. p. < 0.05) b./c. Sample metadata associated with predicted acute and chronic scores. Horizontal brackets show Tukey’s HSD tests in red if p < 0.05 and grey if not significant. CBD: cannot be determined.

**Supplementary Figure 14:**
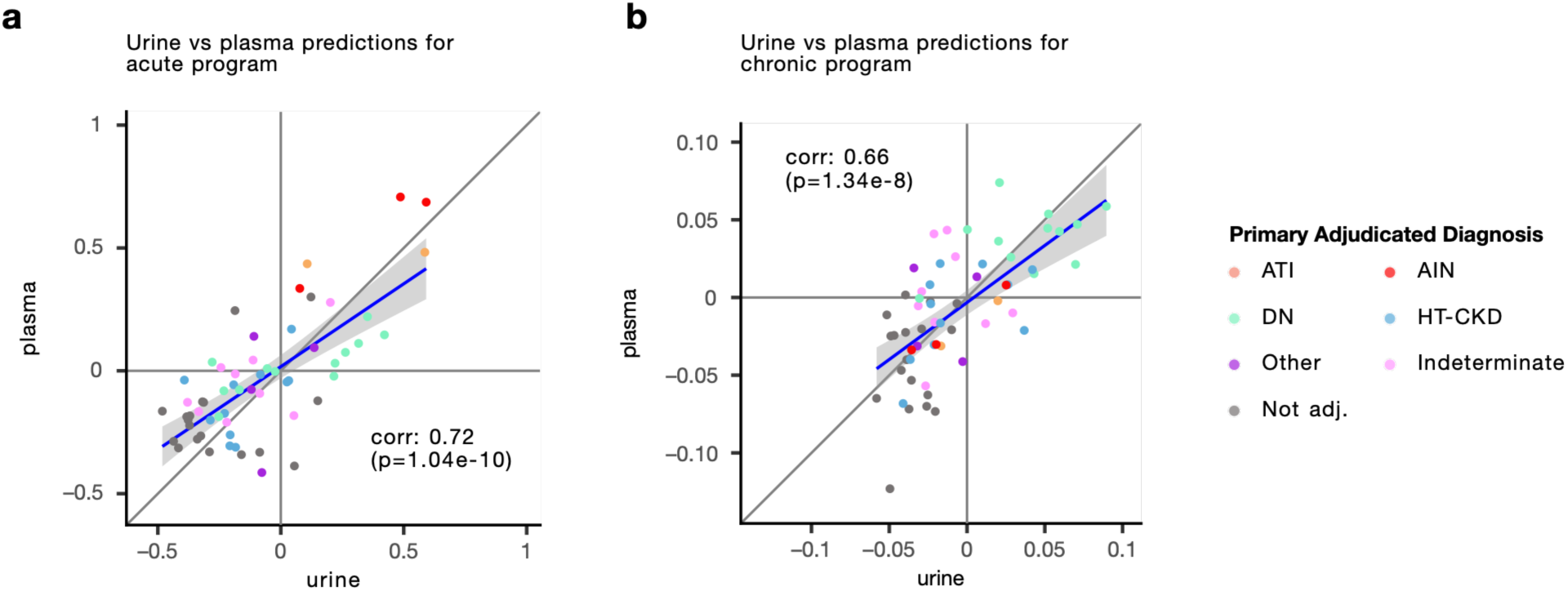
Concordance between predicted acute and chronic scores. a./b. Acute and chronic scores, respectively, predicted from urine and plasma proteins

**Supplementary Table S3:**
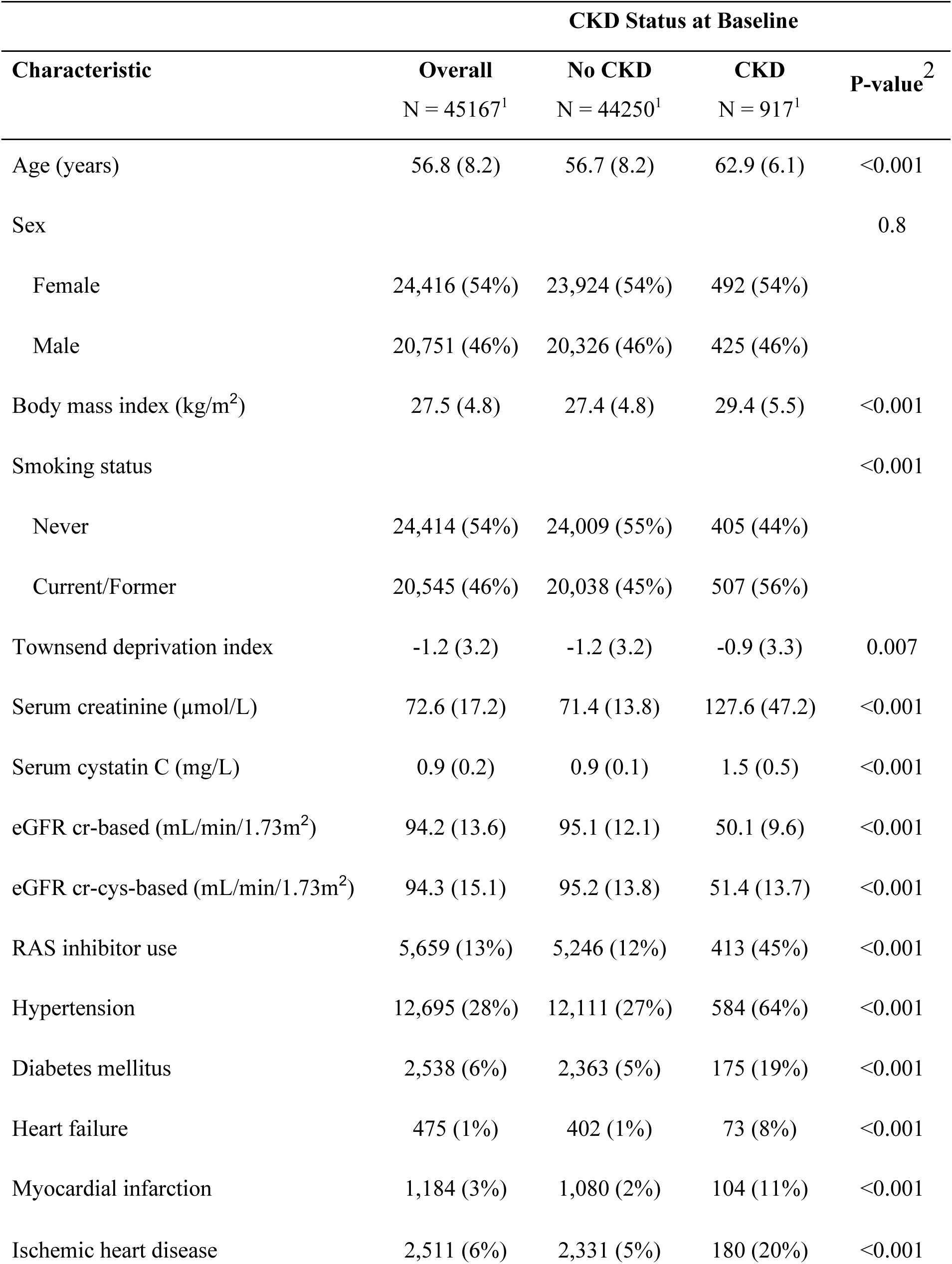
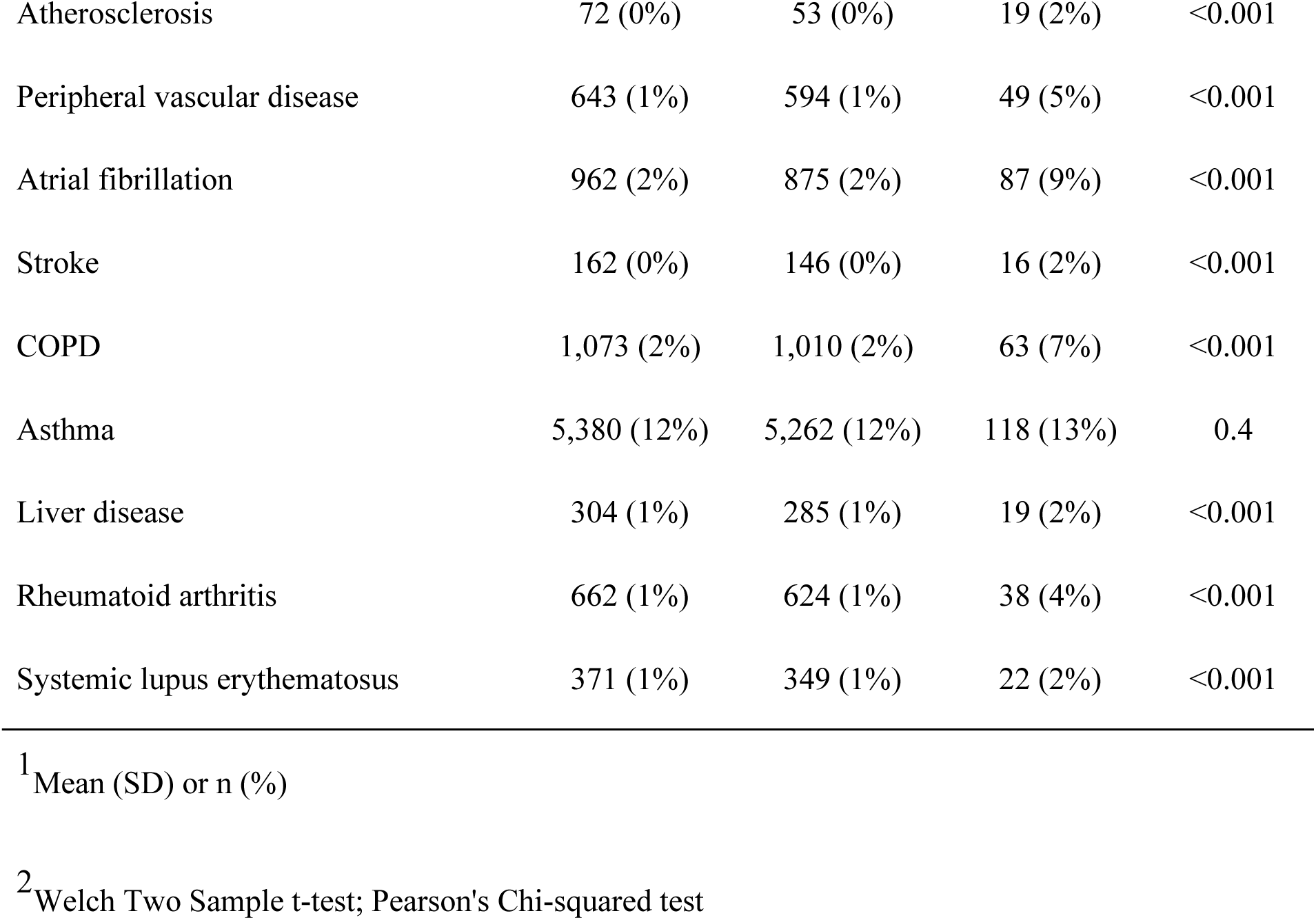
Baseline Characteristics of UK Biobank Participants by CKD Status.

**Supplementary Figure 15:**
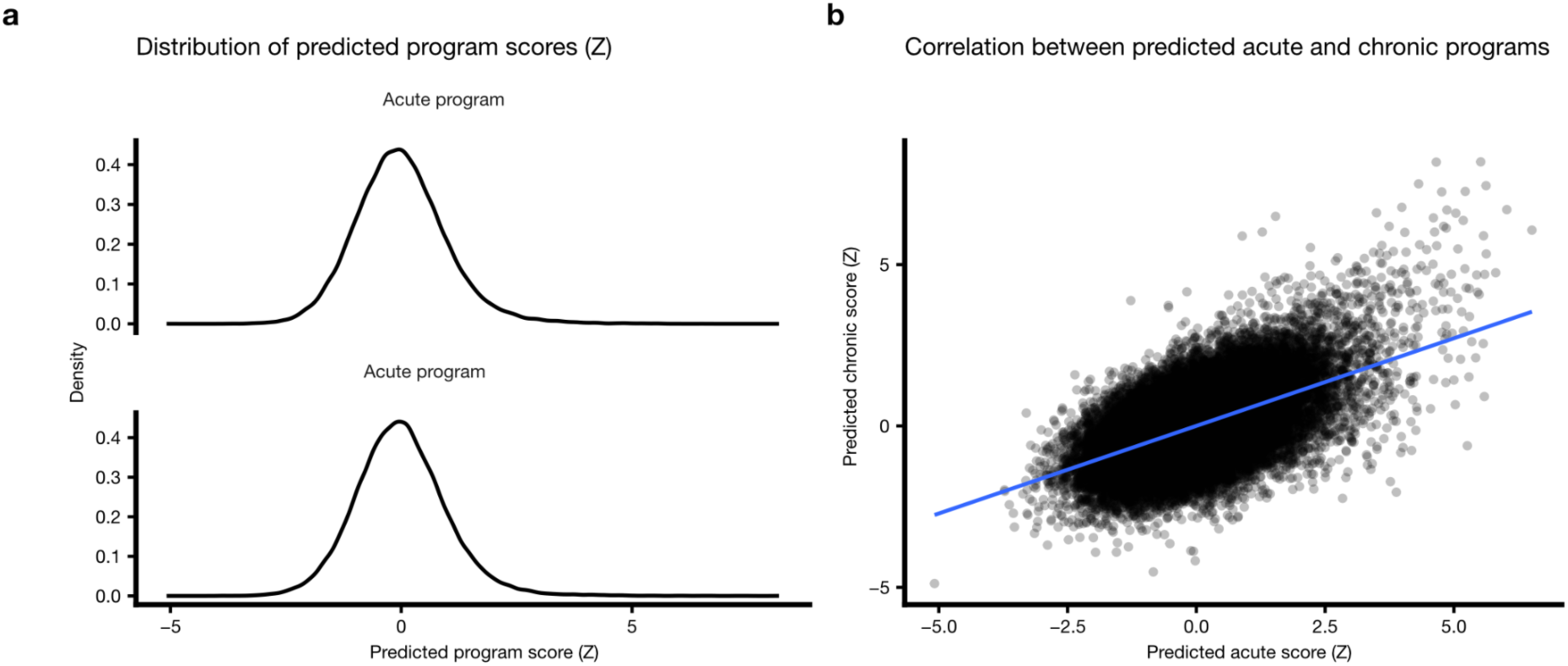
Acute and chronic scores in the UK biobank. a. Distribution of acute and chronic scores (z-transformed) predicted from plasma Olink data b. Relationship between predicted acute and chronic scores in the UK biobank

**Supplementary Table S4:**
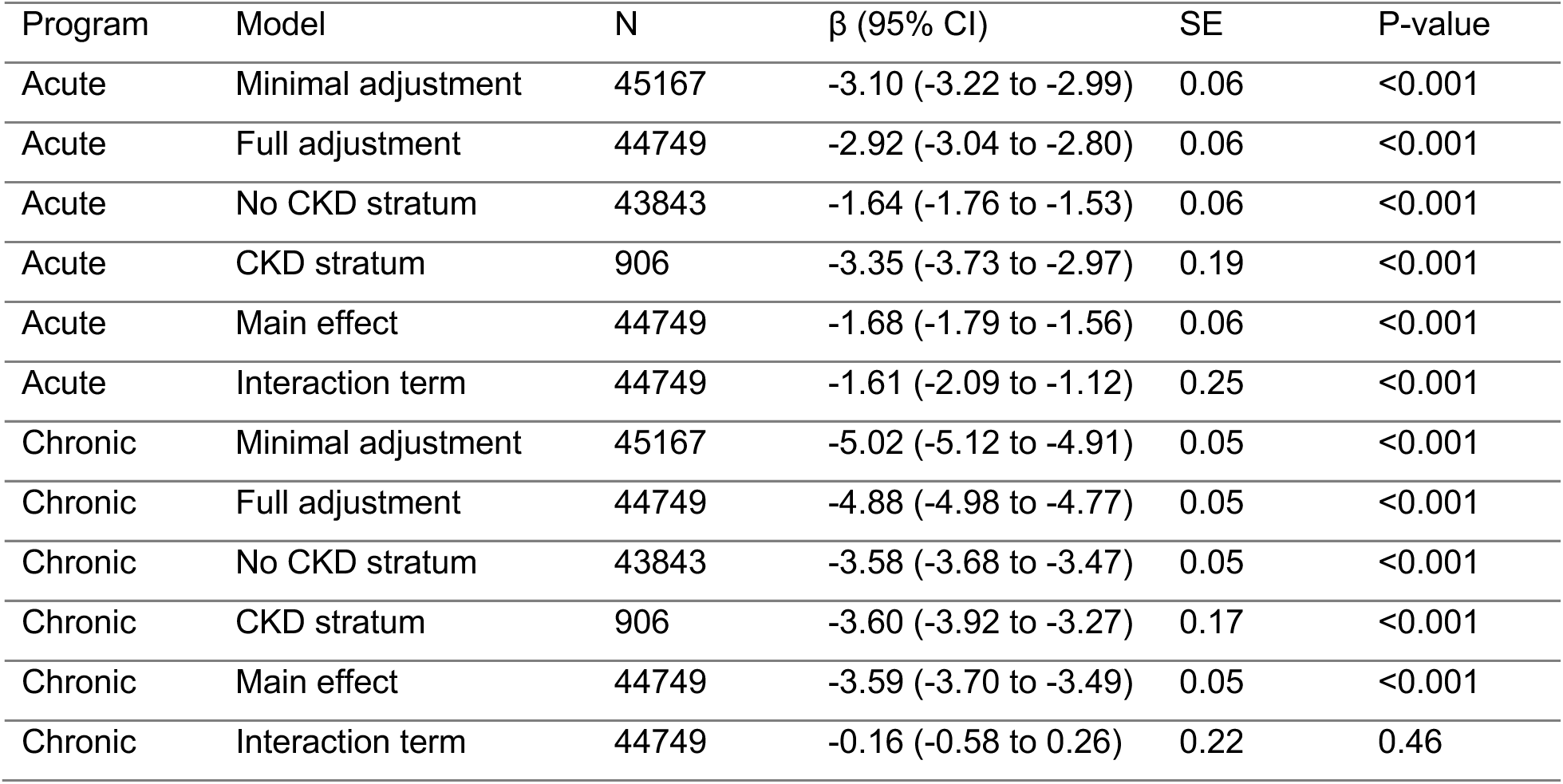
Adjusted association between predicted program scores and eGFR.

**Supplementary Table S5:**
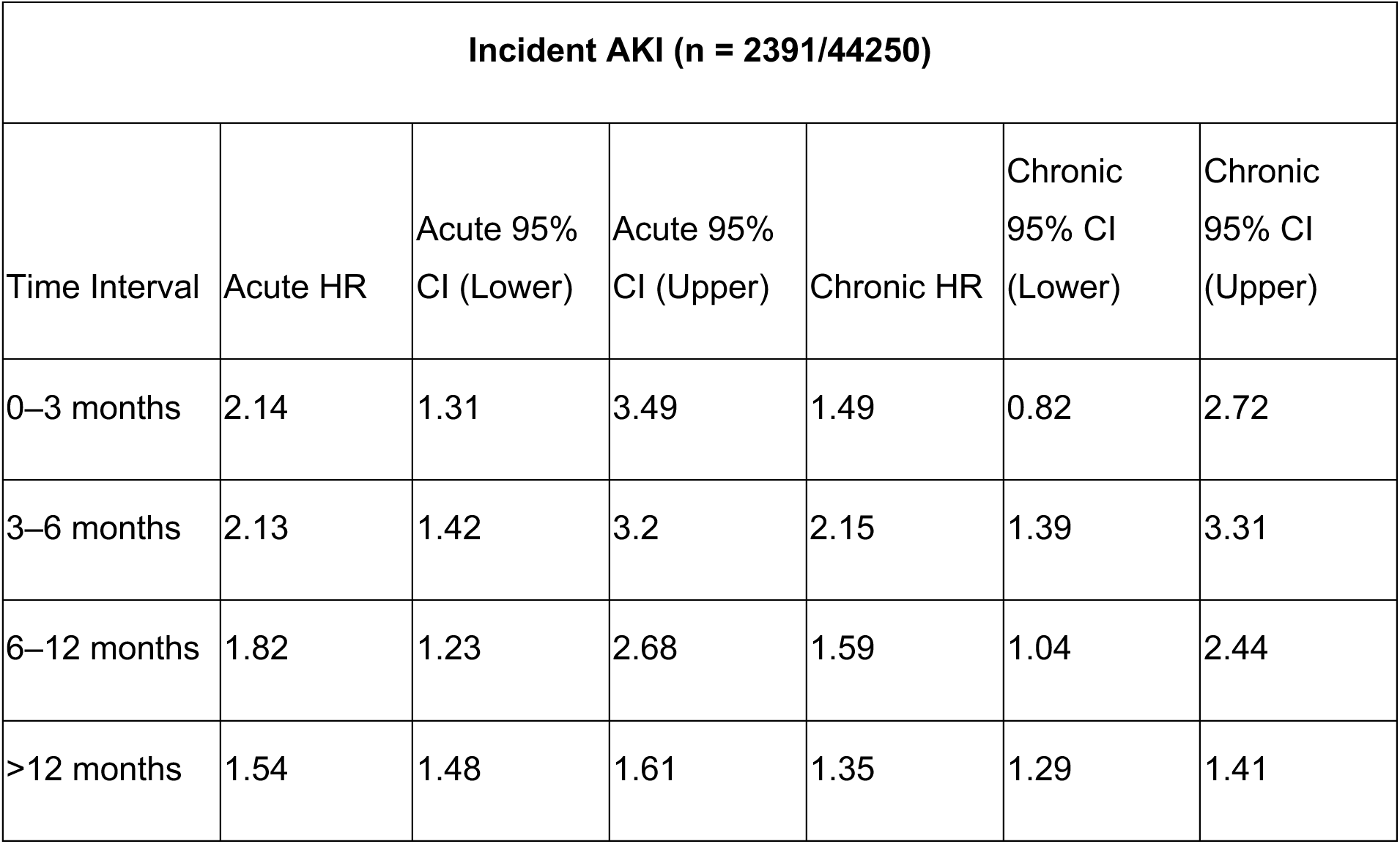
Hazard ratio of incident AKI based on acute or chronic plasma signature in UK biobank.

**Supplementary Table S6:**
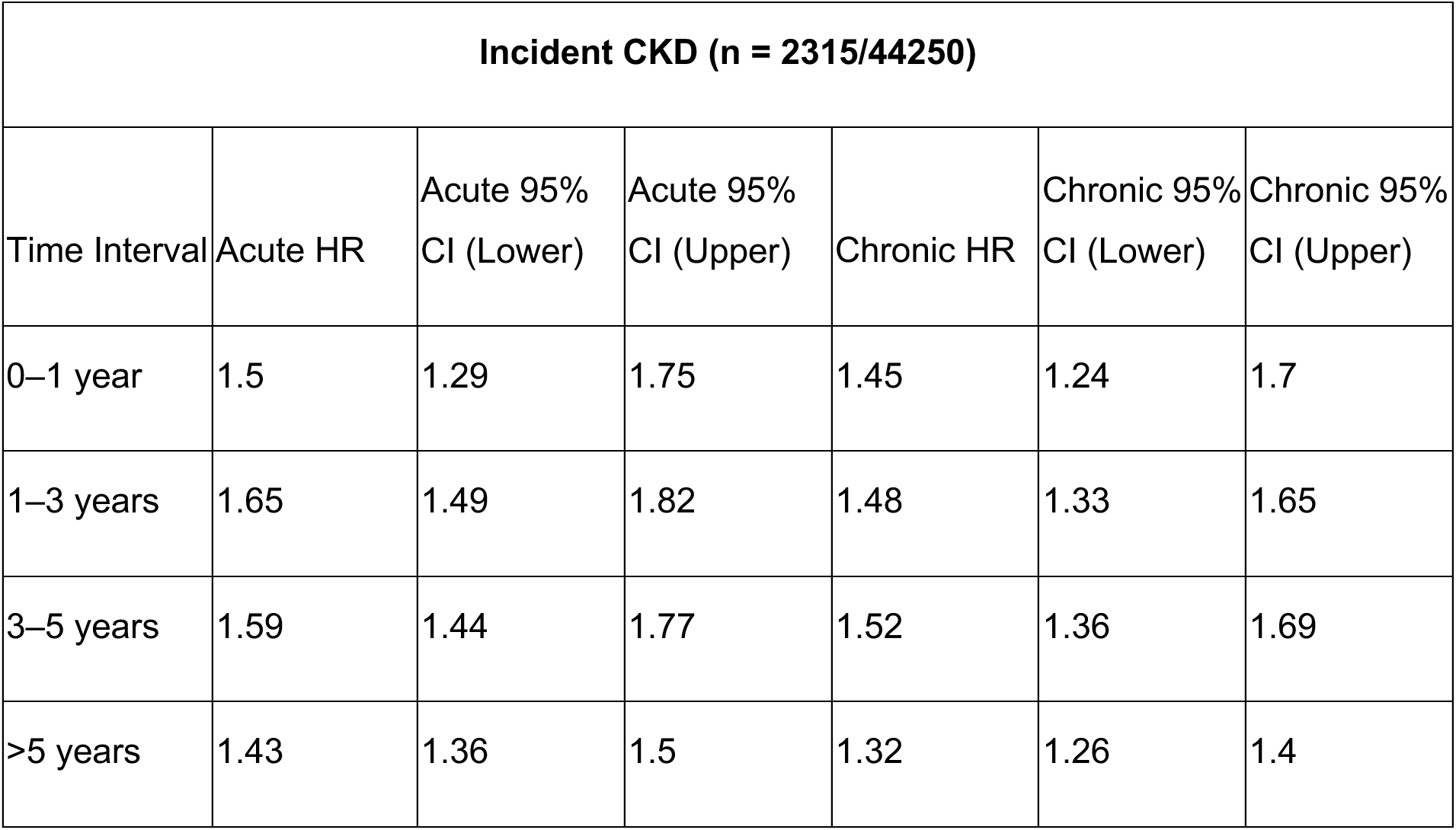
Hazard ratio of incident CKD based on acute or chronic plasma signature in UK biobank.

**Supplementary Table S7:**
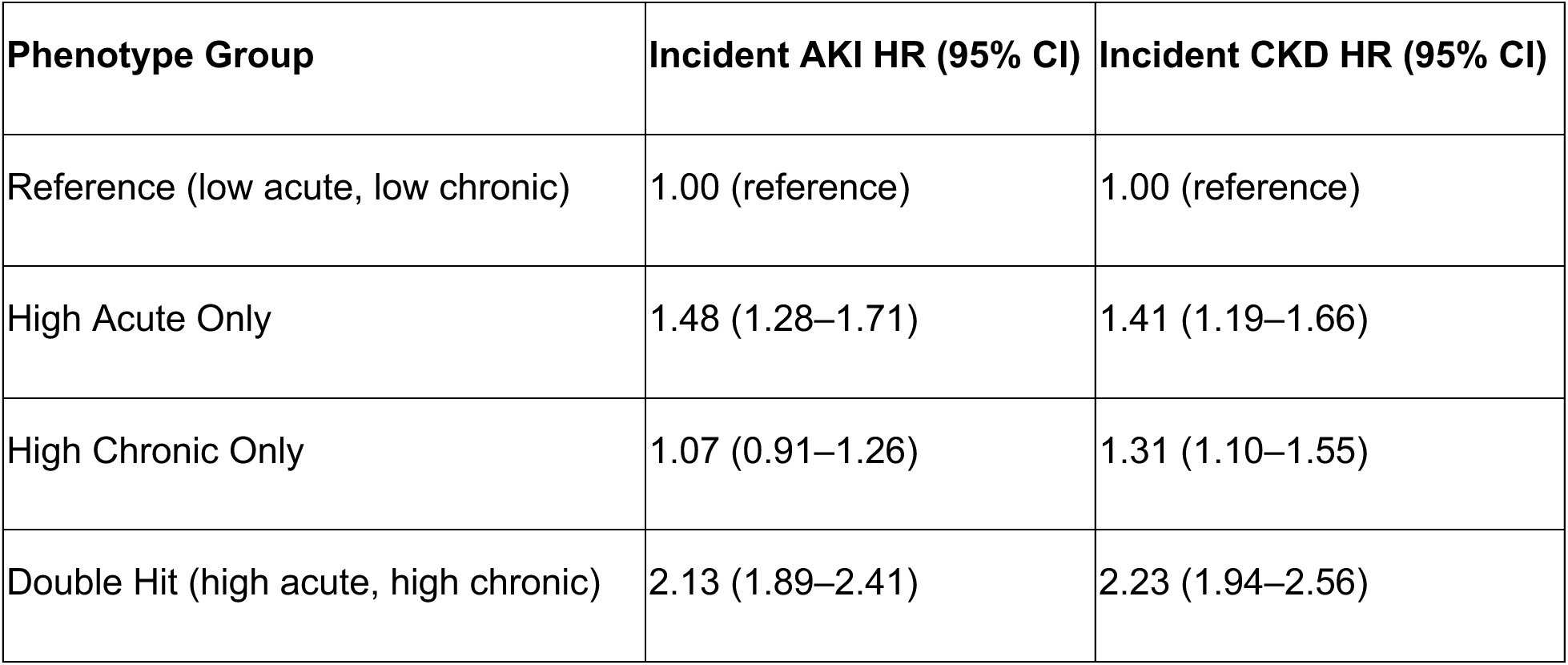
Hazard ratio of incident AKI or CKD based on combination of acute and chronic plasma signature in UK biobank.

## References

1 Kellum, J. A. et al. Acute kidney injury. Nat. Rev. Dis. Primer 7, 52 (2021).

2. Mayer, K. P. et al. Acute kidney injury contributes to worse physical and quality of life outcomes in survivors of critical illness. BMC Nephrol. 23, 137 (2022).

3. Kovesdy, C. P. Epidemiology of chronic kidney disease: an update 2022. Kidney Int. Suppl. 12, 7–11 (2022).

4. Fletcher, B. R. et al. Symptom burden and health-related quality of life in chronic kidney disease: A global systematic review and meta-analysis. PLOS Med. 19, e1003954 (2022).

5. Hoste, E. A. J. et al. Epidemiology of acute kidney injury in critically ill patients: the multinational AKI-EPI study. Intensive Care Med. 41, 1411–1423 (2015).

6. Venkatachalam, M. A. et al. Acute kidney injury: a springboard for progression in chronic kidney disease. Am. J. Physiol.-Ren. Physiol. 298, F1078–F1094 (2010).

7. Chawla, L. S., Eggers, P. W., Star, R. A. & Kimmel, P. L. Acute kidney injury and chronic kidney disease as interconnected syndromes. N. Engl. J. Med. 371, 58–66 (2014).

8. Hsu, C. et al. Post–Acute Kidney Injury Proteinuria and Subsequent Kidney Disease Progression: The Assessment, Serial Evaluation, and Subsequent Sequelae in Acute Kidney Injury (ASSESS-AKI) Study. JAMA Intern. Med. 180, 402–410 (2020).

9. Zarbock, A. et al. Recommendations for clinical trial design in acute kidney injury from the 31st acute disease quality initiative consensus conference. A consensus statement. Intensive Care Med. 50, 1426–1437 (2024).

10. Heerspink, H. J. L. & Kretzler, M. Clinical Trials for Kidney Disease in the Era of Personalized Medicine. J. Am. Soc. Nephrol. JASN 35, 1123–1126 (2024).

11. Birkelo, B. C., Koyner, J. L., Ostermann, M. & Bhatraju, P. K. The Road to Precision Medicine for Acute Kidney Injury. Crit. Care Med. 52, 1127–1137 (2024).

12. Stevens, P. E. et al. KDIGO 2024 Clinical Practice Guideline for the Evaluation and Management of Chronic Kidney Disease. Kidney Int. 105, S117–S314 (2024).

13. Ostermann, M. et al. Acute kidney injury. The Lancet 405, 241–256 (2025).

14. Lindenmeyer, M. T., Alakwaa, F., Rose, M. & Kretzler, M. Perspectives in systems nephrology. Cell Tissue Res. 385, 475–488 (2021).

15. Reznichenko, A. et al. Unbiased kidney-centric molecular categorization of chronic kidney disease as a step towards precision medicine. Kidney Int. 105, 1263–1278 (2024).

16. Bhatraju, P. K. et al. Identification of Acute Kidney Injury Subphenotypes with Differing Molecular Signatures and Responses to Vasopressin Therapy. Am. J. Respir. Crit. Care Med. 199, 863–872 (2019).

17. Wen, Y. & Parikh, C. R. Current concepts and advances in biomarkers of acute kidney injury. Crit. Rev. Clin. Lab. Sci. 58, 354–368 (2021).

18. Schaub, J. A., Alaba, M. & Kretzler, M. A Holistic Approach to AKI: Integrating Clinical and Molecular Data in the Human Kidney. Semin. Nephrol. 45, 151663 (2025).

19. Abedini, A. et al. Single-cell multi-omic and spatial profiling of human kidneys implicates the fibrotic microenvironment in kidney disease progression. Nat. Genet. 56, 1712–1724 (2024).

20. Ledru, N. et al. Predicting proximal tubule failed repair drivers through regularized regression analysis of single cell multiomic sequencing. Nat. Commun. 15, 1291 (2024).

21. Lake, B. B. et al. An atlas of healthy and injured cell states and niches in the human kidney. Nature 619, 585–594 (2023).

22. Kuppe, C. et al. Decoding myofibroblast origins in human kidney fibrosis. Nature 589, 281–286 (2021).

23. Lake, B. B. et al. Cellular and Spatial Drivers of Unresolved Injury and Functional Decline in the Human Kidney. bioRxiv 2025.09.26.678707 (2025) doi:10.1101/2025.09.26.678707.

24. Gerhardt, L. M. S. et al. Lineage Tracing and Single-Nucleus Multiomics Reveal Novel Features of Adaptive and Maladaptive Repair after Acute Kidney Injury. J. Am. Soc. Nephrol. 34, 554–571 (2023).

25. Reck, M. et al. Multiomic analysis of human kidney disease identifies a tractable inflammatory and pro-fibrotic tubular cell phenotype. Nat. Commun. 16, 4745 (2025).

26. Janosevic, D., De Luca, T. & Eadon, M. T. The Kidney Precision Medicine Project and Single-Cell Biology of the Injured Proximal Tubule. Am. J. Pathol. 195, 7–22 (2025).

27. de Boer, I. H. et al. Rationale and design of the Kidney Precision Medicine Project. Kidney Int. 99, 498–510 (2021).

28. Menon, R. et al. Not all controls are made equal: Definition of human kidney reference samples by single cell gene expression profiles. 2025.03.17.25324134 Preprint at 10.1101/2025.03.17.25324134 (2025).

29. Hansen, J. et al. A reference tissue atlas for the human kidney. Sci. Adv. 8, eabn4965 (2022).

30. Schaub, J. A. et al. AKI in the Kidney Precision Medicine Project: Acute Tubular Injury vs. Acute Interstitial Nephritis: TH-OR12. J. Am. Soc. Nephrol. 35, 10.1681/ASN.2024cxh8wvky (2024).

31. Limonte, C. P. et al. Case Series of Histopathological Findings in Chronic Kidney Disease: Insights From the Kidney Precision Medicine Project. Kidney Med. 8, (2026).

32. Limonte, C. P. et al. Redefining kidney disease: Clinico-pathological and molecular findings from the Kidney Precision Medicine Project. 2026.02.24.26347022 Preprint at 10.64898/2026.02.24.26347022 (2026).

33. Ramirez Flores, R., Lanzer, J., Dimitrov, D., Velten, B. & Saez-Rodriguez, J. Multicellular factor analysis of single-cell data for a tissue-centric understanding of disease. eLife 12, e93161 (2023).

34. Kellum, J. A., Lameire, N., & for the KDIGO AKI Guideline Work Group. Diagnosis, evaluation, and management of acute kidney injury: a KDIGO summary (Part 1). Crit. Care 17, 204 (2013).

35. Misra, P. S., Szeto, S. G., Krizova, A., Gilbert, R. E. & Yuen, D. A. Renal histology in diabetic nephropathy predicts progression to end-stage kidney disease but not the rate of renal function decline. BMC Nephrol. 21, 285 (2020).

36. Menn-Josephy, H. et al. Renal interstitial fibrosis: an imperfect predictor of kidney disease progression in some patient cohorts. Am. J. Nephrol. 44, 289–299 (2016).

37. Hommos, M. S. et al. Global glomerulosclerosis with nephrotic syndrome; the clinical importance of age adjustment. Kidney Int. 93, 1175–1182 (2018).

38. Bhatraju, P. K. et al. Association Between Early Recovery of Kidney Function After Acute Kidney Injury and Long-term Clinical Outcomes. *JAMA Netw*. Open 3, e202682 (2020).

39. Li, Z. et al. Chromatin-accessibility estimation from single-cell ATAC-seq data with scOpen. Nat. Commun. 12, 6386 (2021).

40. Fu, H. et al. Tenascin-C Is a Major Component of the Fibrogenic Niche in Kidney Fibrosis. J. Am. Soc. Nephrol. 28, 785 (2017).

41. Alicic, R. Z., Rooney, M. T. & Tuttle, K. R. Diabetic Kidney Disease: Challenges, Progress, and Possibilities. Clin. J. Am. Soc. Nephrol. 12, 2032 (2017).

42. Thomas, H. Y. & Ford Versypt, A. N. Pathophysiology of mesangial expansion in diabetic nephropathy: mesangial structure, glomerular biomechanics, and biochemical signaling and regulation. J. Biol. Eng. 16, 19 (2022).

43. Müller-Dott, S. et al. Expanding the coverage of regulons from high-confidence prior knowledge for accurate estimation of transcription factor activities. Nucleic Acids Res. 51, 10934–10949 (2023).

44. Masenga, S. K., Desta, S., Hatcher, M., Kirabo, A. & Lee, D. L. How PPAR-alpha mediated inflammation may affect the pathophysiology of chronic kidney disease. Curr. Res. Physiol. 8, 100133 (2025).

45. Gold, L. et al. Aptamer-Based Multiplexed Proteomic Technology for Biomarker Discovery. PLOS ONE 5, e15004 (2010).

46. SOMAmer reagents and the SomaScan platform: Chemically modified aptamers and their applications in therapeutics, diagnostics, and proteomics. in RNA Therapeutics 171–260 (Academic Press, 2022). doi:10.1016/B978-0-12-821595-1.00007-5.

47. Candia, J., Daya, G. N., Tanaka, T., Ferrucci, L. & Walker, K. A. Assessment of variability in the plasma 7k SomaScan proteomics assay. Sci. Rep. 12, 17147 (2022).

48. Kaleta, B. The role of osteopontin in kidney diseases. Inflamm. Res. 68, 93–102 (2019).

49. Ichimura, T., Hung, C. C., Yang, S. A., Stevens, J. L. & Bonventre, J. V. Kidney injury molecule-1: a tissue and urinary biomarker for nephrotoxicant-induced renal injury. Am. J. Physiol.-Ren. Physiol. 286, F552–F563 (2004).

50. Vaidya, V. S. et al. Kidney injury molecule-1 outperforms traditional biomarkers of kidney injury in preclinical biomarker qualification studies. Nat. Biotechnol. 28, 478–485 (2010).

51. Aloor, J. J. et al. Leucine-rich repeats and calponin homology containing 4 (Lrch4) regulates the innate immune response. J. Biol. Chem. 294, 1997–2008 (2019).

52. Fett, J. W. et al. Isolation and characterization of angiogenin, an angiogenic protein from human carcinoma cells. Biochemistry 24, 5480–5486 (1985).

53. Tavernier, Q. et al. Urinary Angiogenin Reflects the Magnitude of Kidney Injury at the Infrahistologic Level. J. Am. Soc. Nephrol. JASN 28, 678–690 (2017).

54. Gurung, R. L. et al. Association of Plasma Angiogenin With Risk of Incident End-Stage Kidney Disease in Individuals With Type 2 Diabetes. Diabetes 74, 998–1006 (2025).

55. Alikhan, M. A. et al. Colony-Stimulating Factor-1 Promotes Kidney Growth and Repair via Alteration of Macrophage Responses. Am. J. Pathol. 179, 1243–1256 (2011).

56. Wang, Y. et al. Proximal tubule-derived Colony Stimulating Factor-1 mediates polarization of renal macrophages and dendritic cells, and recovery in acute kidney injury. Kidney Int. 88, 1274–1282 (2015).

57. Hoste, E. et al. Identification and validation of biomarkers of persistent acute kidney injury: the RUBY study. Intensive Care Med. 46, 943–953 (2020).

58. Kellum, J. A. et al. CCL14 testing to guide clinical practice in patients with AKI: Results from an international expert panel. J. Crit. Care 82, 154816 (2024).

59. Zhu, H. et al. Tenascin-C promotes acute kidney injury to chronic kidney disease progression by impairing tubular integrity via αvβ6 integrin signaling. Kidney Int. 97, 1017–1031 (2020).

60. Caspers, T., Boor, P. & Klinkhammer, B. M. The roles of Tenascin C in kidney diseases. Am. J. Physiol.-Cell Physiol. 330, C552–C569 (2026).

61. Rovin, B. H., Doe, N. & Tan, L. C. Monocyte chemoattractant protein-1 levels in patients with glomerular disease. Am. J. Kidney Dis. 27, 640–646 (1996).

62. Munshi, R. et al. MCP-1 Gene Activation Marks Acute Kidney Injury. J. Am. Soc. Nephrol. 22, 165 (2011).

63. Titan, S. M. et al. Urinary MCP-1 and RBP: Independent predictors of renal outcome in macroalbuminuric diabetic nephropathy. J. Diabetes Complications 26, 546–553 (2012).

64. Puthumana, J. et al. Biomarkers of inflammation and repair in kidney disease progression. J. Clin. Invest. 131, (2021).

65. Tuechler, N. et al. Dynamic multi-omics and mechanistic modeling approach uncovers novel mechanisms of kidney fibrosis progression. Mol. Syst. Biol. 21, 1030–1065 (2025).

66. Hwang, H. J., et al. FLRT2 prevents endothelial cell senescence and vascular aging by regulating the ITGB4/mTORC2/p53 signaling pathway. JCI Insight 9, e172678 (2024).

67. Benetó, N., Vilageliu, L., Grinberg, D. & Canals, I. Sanfilippo Syndrome: Molecular Basis, Disease Models and Therapeutic Approaches. Int. J. Mol. Sci. 21, 7819 (2020).

68. Konno, K. et al. A molecule that is associated with Toll-like receptor 4 and regulates its cell surface expression. Biochem. Biophys. Res. Commun. 339, 1076–1082 (2006).

69. Ghait, M. et al. The TLR-chaperone CNPY3 is a critical regulator of NLRP3-inflammasome activation. Eur. J. Immunol. 52, 907–923 (2022).

70. Zipfel, P. F. & Skerka, C. Complement regulators and inhibitory proteins. Nat. Rev. Immunol. 9, 729–740 (2009).

71. Michielsen, L. A., van Zuilen, A. D., Kardol-Hoefnagel, T., Verhaar, M. C. & Otten, H. G. Association Between Promoter Polymorphisms in CD46 and CD59 in Kidney Donors and Transplant Outcome. Front. Immunol. 9, (2018).

72. Buettner, R. et al. Activated Signal Transducers and Activators of Transcription 3 Signaling Induces CD46 Expression and Protects Human Cancer Cells from Complement-Dependent Cytotoxicity. Mol. Cancer Res. 5, 823–832 (2007).

73. Ngo, D. et al. Circulating testican-2 is a podocyte-derived marker of kidney health. Proc. Natl. Acad. Sci. 117, 25026–25035 (2020).

74. Char, R. & Pierre, P. The RUFYs, a Family of Effector Proteins Involved in Intracellular Trafficking and Cytoskeleton Dynamics. Front. Cell Dev. Biol. 8, 779 (2020).

75. Cormont, M., Mari, M., Galmiche, A., Hofman, P. & Le Marchand-Brustel, Y. A FYVE-finger-containing protein, Rabip4, is a Rab4 effector involved in early endosomal traffic. Proc. Natl. Acad. Sci. U. S. A. 98, 1637–1642 (2001).

76. Gosney, J. A., Wilkey, D. W., Merchant, M. L. & Ceresa, B. P. Proteomics reveals novel protein associations with early endosomes in an epidermal growth factor–dependent manner. J. Biol. Chem. 293, 5895–5908 (2018).

77. Houghton, F. J. et al. Arl5b is a Golgi-localised small G protein involved in the regulation of retrograde transport. Exp. Cell Res. 318, 464–477 (2012).

78. Ju, W. et al. Tissue transcriptome-driven identification of epidermal growth factor as a chronic kidney disease biomarker. Sci. Transl. Med. 7, 316ra193-316ra193 (2015).

79. Zhou, J. et al. Urinary epidermal growth factor predicts complete remission of proteinuria in Chinese children with IgA nephropathy. Pediatr. Res. 94, 747–755 (2023).

80. Canela, V. H. et al. A spatially anchored transcriptomic atlas of the human kidney papilla identifies significant immune injury in patients with stone disease. Nat. Commun. 14, 4140 (2023).

81. Lundberg, M., Eriksson, A., Tran, B., Assarsson, E. & Fredriksson, S. Homogeneous antibody-based proximity extension assays provide sensitive and specific detection of low-abundant proteins in human blood. Nucleic Acids Res. 39, e102 (2011).

82. Souza, V. C. D. et al. Schwartz Formula: Is One k-Coefficient Adequate for All Children? PLOS ONE 7, e53439 (2012).

83. Grubb, A. et al. Generation of a New Cystatin C–Based Estimating Equation for Glomerular Filtration Rate by Use of 7 Assays Standardized to the International Calibrator. Clin. Chem. 60, 974–986 (2014).

84. Berg, U. B. et al. New standardized cystatin C and creatinine GFR equations in children validated with inulin clearance. Pediatr. Nephrol. 30, 1317–1326 (2015).

85. McCown, P. J. et al. A Human Glomerular Disease Atlas defines the APOL1-JAK-STAT feed forward loop in focal segmental glomerulosclerosis. 2025.09.12.25335572 Preprint at 10.1101/2025.09.12.25335572 (2025).

86. Heumos, L. et al. Best practices for single-cell analysis across modalities. Nat. Rev. Genet. 24, 550–572 (2023).

87. Le, D. et al. Plasma Biomarkers and Incident CKD Among Individuals Without Diabetes. Kidney Med. 5, 100719 (2023).

88. Schrauben, S. J. et al. Urine Biomarkers for Diabetic Kidney Disease Progression in Participants of the Chronic Renal Insufficiency Cohort Study. Clin. J. Am. Soc. Nephrol. 20, 958 (2025).

89. Limonte, C. P. & Schaub, J. A. Redefining kidney disease – Clinico-histopathological and molecular findings from the Kidney Precision Medicine Project. Preprint at (2025).

90. Barisoni, L. et al. Reproducibility of the NEPTUNE descriptor-based scoring system on whole-slide images and histologic and ultrastructural digital images. Mod. Pathol. 29, 671–684 (2016).

91. Dimitrov, D. et al. LIANA+ provides an all-in-one framework for cell–cell communication inference. Nat. Cell Biol. 26, 1613–1622 (2024).

92. Badia-i-Mompel, P., et al. decoupleR: ensemble of computational methods to infer biological activities from omics data. Bioinforma. Adv. 2, vbac016 (2022).

93. Argelaguet, R. et al. MOFA+: a statistical framework for comprehensive integration of multi-modal single-cell data. Genome Biol. 21, 111 (2020).

94. Bredikhin, D., Kats, I. & Stegle, O. MUON: multimodal omics analysis framework. Genome Biol. 23, 42 (2022).

95. Seabold, S. & Perktold, J. Statsmodels: Econometric and Statistical Modeling with Python. scipy https://doi.org/10.25080/Majora-92bf1922-011 (2010) doi:10.25080/Majora-92bf1922-011.

96. Liberzon, A. et al. The Molecular Signatures Database (MSigDB) hallmark gene set collection. Cell Syst. 1, 417–425 (2015).

97. Wolf, F., Angerer, P. & Theis, F. SCANPY: large-scale single-cell gene expression data analysis. Genome Biol. 19, 15 (2018).

98. Rideout, J. R., et al. biocore/scikit-bio: scikit-bio 0.5.9: Maintenance release. Zenodo 10.5281/zenodo.8209901 (2023).

99. Bates, D., Mächler, M., Bolker, B. & Walker, S. Fitting Linear Mixed-Effects Models Using lme4. J. Stat. Softw. 67, 1–48 (2015).

100. Jaeger, B. C., Edwards, L. J., Das, K. & Sen, P. K. An R2 statistic for fixed effects in the generalized linear mixed model. J. Appl. Stat. https://www.tandfonline.com/doi/abs/10.1080/02664763.2016.1193725 (2017).

101. Schaub, J. A. et al. SGLT2 inhibitors mitigate kidney tubular metabolic and mTORC1 perturbations in youth-onset type 2 diabetes. J. Clin. Invest. 133, (2023).

102. Neusser, M. A. et al. Human Nephrosclerosis Triggers a Hypoxia-Related Glomerulopathy. Am. J. Pathol. 176, 594–607 (2010).

103. Woroniecka, K. I. et al. Transcriptome Analysis of Human Diabetic Kidney Disease. Diabetes 60, 2354–2369 (2011).

104. Liu, P. et al. Transcriptomic and Proteomic Profiling Provides Insight into Mesangial Cell Function in IgA Nephropathy. J. Am. Soc. Nephrol. JASN 28, 2961–2972 (2017).

105. Fan, Y. et al. Comparison of Kidney Transcriptomic Profiles of Early and Advanced Diabetic Nephropathy Reveals Potential New Mechanisms for Disease Progression. Diabetes 68, 2301–2314 (2019).

106. Park, S. et al. RNA-Seq profiling of microdissected glomeruli identifies potential biomarkers for human IgA nephropathy. Am. J. Physiol.-Ren. Physiol. 10.1152/ajprenal.00037.2020 (2020) doi:10.1152/ajprenal.00037.2020.

107. Zhao, T. et al. Transcriptomics-proteomics Integration reveals alternative polyadenylation driving inflammation-related protein translation in patients with diabetic nephropathy. J. Transl. Med. 21, 86 (2023).

108. Virtanen, P. et al. SciPy 1.0: fundamental algorithms for scientific computing in Python. Nat. Methods 17, 261–272 (2020).

