## Supplemental Materials for "Shared multicellular injury programs of acute and chronic kidney disease enable mechanistic patient stratification"

### Supplementary Figures and Tables

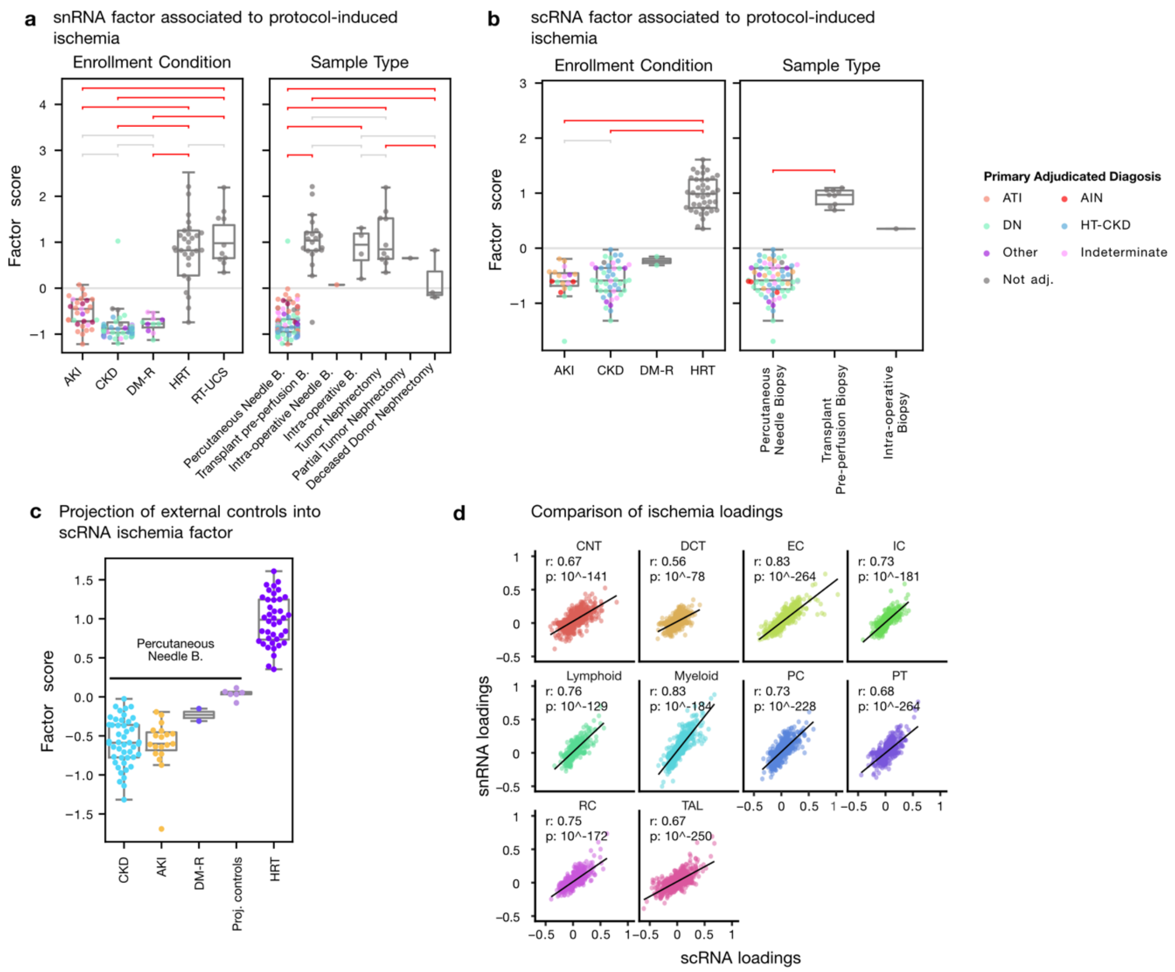

**Supplementary Figure 1:** Factors capture ischemic signature due to procurement

*a./b. Technical factors in snRNA and scRNA separate samples based on enrollment conditions, and is confounded with sample procurement method c. Projection of healthy reference samples from the CROCODILE study (percutaneous biopsies) onto scRNA factor capturing ischemia d. Correlation between factor loadings from snRNA and scRNA for factors relating to ischemia signature*

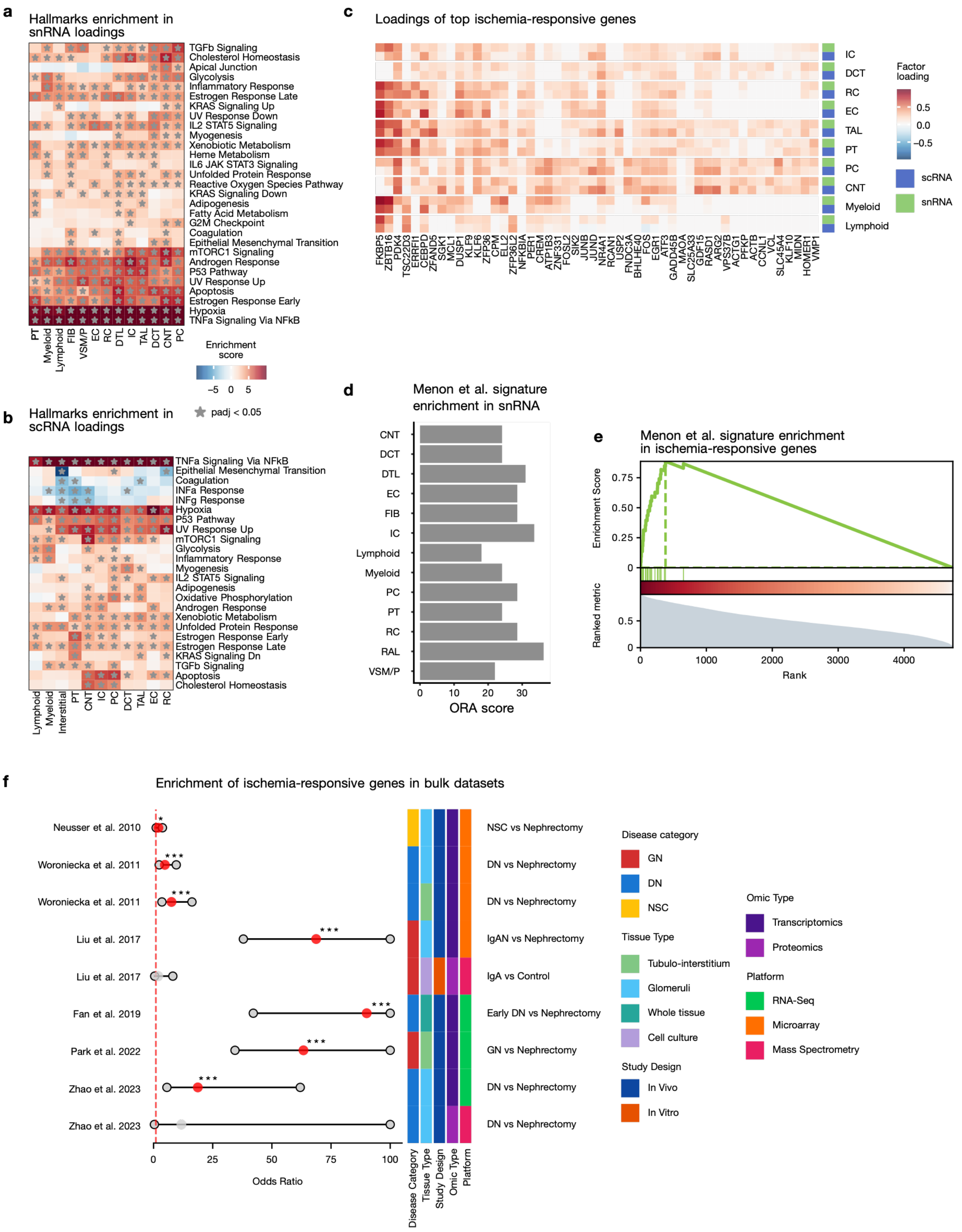

**Supplementary Figure 2:** Ischemic signature is captured by factors in snRNA and scRNA

*a./b. MSigDB hallmark enrichment in factor shows increased inflammatory signaling, hypoxia and apoptosis in snRNA and scRNA respectively c. Ischemia-induced gene signature based on consensus between snRNA and scRNA factor loadings across cell types d. Enrichment of gene signature from Menon et al.[28] (derived from scRNA) in the factor loadings of snRNA e. GSEA running score in the ischemia ranking shown in c. of the signature from Menon et al. f. Odds ratio of finding ischemia signature genes (from c) in genes more highly expressed in controls from bulk RNA and protein studies which use nephrectomy samples as control.*

**Supplementary Table S1:** scRNA cohort characteristics

| | | **AKI** | **CKD** | **DM-R** |
|---|---|---|---|---|
| | **N** | **19** | **46** | **2** |
| **General demographics** | Age | 55.00 [35.50; 62.00] | 62.50 [54.50; 66.75] | 44.50 [38.25; 50.75] |
| | BMI | 25.61 [23.55; 29.45] | 30.86 [28.27; 36.22] | 35.45 [29.32; 41.57] |
| | Female | 7 (36.8%) | 23 (50.0%) | 1 (50.0%) |
| | Male | 12 (63.2%) | 23 (50.0%) | 1 (50.0%) |
| | Has Diabetes | 8 (42.1%) | 39 (84.8%) | 2 (100.0%) |
| | Has Hypertension | 7 (36.8%) | 46 (100.0%) | 1 (50.0%) |
| **Primary Adjudicated Category** | AIN | 3 (15.8%) | 0 (0.0%) | 0 (0.0%) |
| | ATI | 8 (42.1%) | 0 (0.0%) | 0 (0.0%) |
| | DN | 4 (21.1%) | 19 (41.3%) | 2 (100.0%) |
| | HT-CKD | 0 (0.0%) | 14 (30.4%) | 0 (0.0%) |
| | Indeterminate | 3 (15.8%) | 8 (17.4%) | 0 (0.0%) |
| | Other | 1 (5.3%) | 4 (8.7%) | 0 (0.0%) |
| | Not Adj. | 0 (0.0%) | 1 (2.2%) | 0 (0.0%) |
| **Lab values** | Most Recent eGFR | 51.43 [36.53; 79.06] | 45.61 [38.18; 58.18] | 114.29 [109.56; 119.02] |
| | Urinary ACR | 2.73 [2.40; 3.02] | 2.60 [1.51; 3.39] | 0.95 [0.89; 1.01] |
| | Urinary PCR | 2.74 [2.41; 3.29] | 3.11 [2.31; 3.42] | N/A |
| | Most Recent HbA1c | 6.00 [5.50; 8.05] | 7.10 [6.30; 7.95] | 8.25 [8.12; 8.38] |
| **Treatments** | On SGLT2 Inhibitor | 1 (5.3%) | 13 (28.3%) | 0 (0.0%) |
| | On Insulin | 2 (10.5%) | 22 (47.8%) | 2 (100.0%) |
| | On RAAS Inhibitor | 3 (15.8%) | 39 (84.8%) | 1 (50.0%) |
| | On Diuretics | 4 (21.1%) | 22 (47.8%) | 0 (0.0%) |
| | On ACEi/ARB | 3 (15.8%) | 37 (80.4%) | 1 (50.0%) |
| | On Metformin | 2 (10.5%) | 15 (32.6%) | 0 (0.0%) |
| **TIV scoring** | Interstitial Fibrosis (%) | 0.30 [0.07; 0.55] | 0.28 [0.10; 0.40] | 0.01 [0.01; 0.01] |
| | Tubular Atrophy (%) | 0.15 [0.04; 0.53] | 0.20 [0.10; 0.31] | 0.01 [0.00; 0.01] |
| | Tubular Injury (%) | 0.20 [0.10; 0.40] | 0.20 [0.10; 0.30] | 0.25 [0.18; 0.33] |
| **All adjudication findings** | Acute Tubular Injury | 15.0 (78.9%) | 18.0 (39.1%) | 0.0 (0.0%) |
| | Acute Interstitial Nephritis | 5.0 (26.3%) | 0.0 (0.0%) | 0.0 (0.0%) |
| | Diabetic Kidney Disease | 5.0 (26.3%) | 28.0 (60.9%) | 2.0 (100.0%) |
| | Hypertensive Kidney Disease | 0.0 (0.0%) | 20.0 (43.5%) | 0.0 (0.0%) |
| | Diabetic Glomerulopathy | 5 (26.3%) | 28 (60.9%) | 2 (100.0%) |
| | Global Glomerulosclerosis | 4.0 (21.1%) | 31.0 (67.4%) | 0.0 (0.0%) |
| | Segmental Sclerosis | 5.0 (26.3%) | 14.0 (30.4%) | 0.0 (0.0%) |

*Clinical, adjudication and histology description of samples in scRNA. Continuous variables are shown as median [IQR], categorical variables as count (percentage). ACR: log2 albumin-to-creatinine ratio, PCR: log2 protein-to-creatinine ratio, eGFR: estimated glomerular filtration rate in ml/min/1.73m2. Primary adjudicated categories are mutually exclusive, whereas individual adjudication findings are not. Adjudication and TIV scoring was performed on a separate biopsy core than the one used for snRNA or scRNA. N/A: not available*

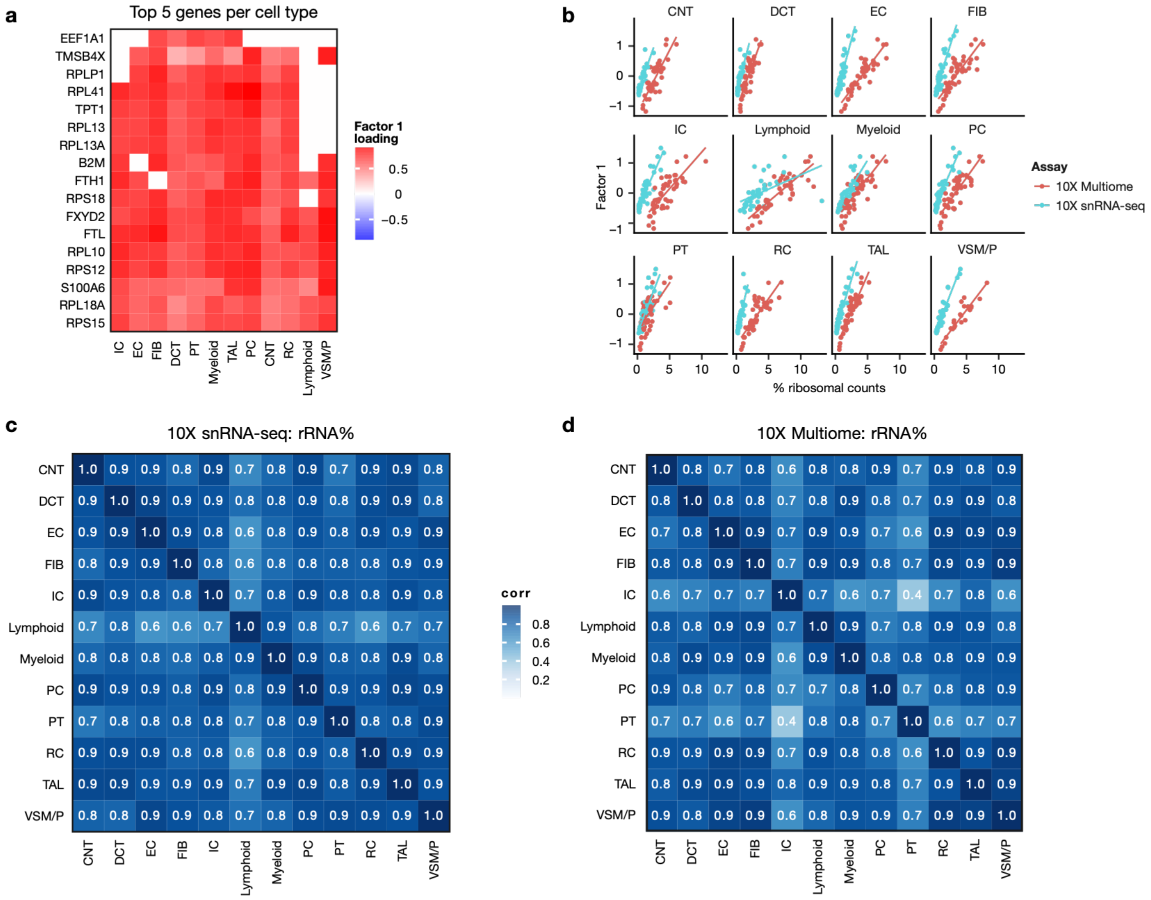

**Supplementary Figure 3:** Factor 1 explains ribosomal content differences between samples in snRNA

*a. Top 5 Factor 1 gene loadings per cell type from snRNA. b. snRNA factor 1 scores relative to percentage of ribosomal counts c./d. Pearson correlation of ribosomal content across cell types pseudobulks in 10X snRNA-seq and 10X Multiome samples*

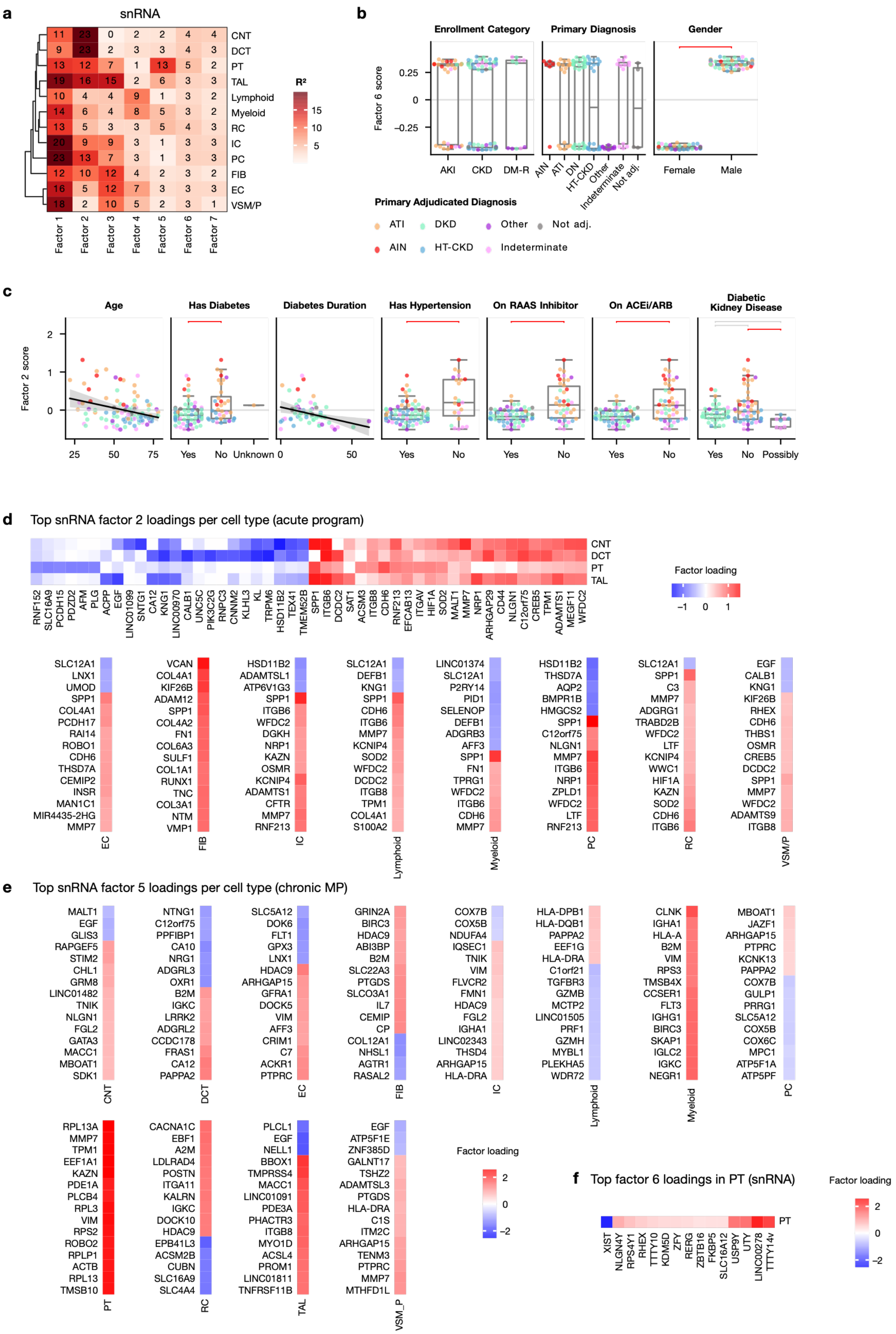

a
snRNA
CNT
DCT
PT
TAL
Lymphoid
Myeloid
RC
IC
PC
FIB
EC
VSM/P
R²
15
10
5
Factor 1
Factor 2
Factor 3
Factor 4
Factor 5
Factor 6
Factor 7
b
Enrollment Category
Primary Diagnosis
Gender
Factor 6 score
0.25
0
−0.25
AKI
CKD
DM-R
AIN
ATI
DN
HT-CKD
Other
Indeterminate
Not adj.
Female
Male
Primary Adjudicated Diagnosis
ATI
DKD
Other
Not adj.
AIN
HT-CKD
Indeterminate
c
Age
Has Diabetes
Diabetes Duration
Has Hypertension
On RAAS Inhibitor
On ACEi/ARB
Diabetic Kidney Disease
Factor 2 score
2
1
0
25
50
75
Yes
No
Unknown
0
50
Possibly
d
Top snRNA factor 2 loadings per cell type (acute program)
Factor loading
−1
0
1
EC
FIB
IC
Lymphoid
Myeloid
PC
RC
VSM/P
e
Top snRNA factor 5 loadings per cell type (chronic MP)
CNT
DCT
EC
FIB
IC
Lymphoid
Myeloid
PC
PT
RC
TAL
VSM_P
Factor loading
2
0
−2
f
Top factor 6 loadings in PT (snRNA)
Factor loading
PT
XIST
NLGN4Y
RPS4Y1
RHEX
TTTY10
KDM5D
ZFY
RERG
ZBTB16
FKBP5
SLC16A12
USP9Y
UTY
LINC00278
TTTY14

### Supplementary Figure 4:

*a. Gene expression variance explained by MCFA model in snRNA per cell type b. Factor 6 captures sex differences c. additional metadata associations with Factor 2 of snRNA. Horizontal brackets show Tukey's HSD tests in red if p < 0.05 and grey if not significant. CBD: Cannot be Determined d. Top gene loadings in factor 2 (acute program) per cell type e. Top loadings in factor 5 (chronic program) per cell type f. Top loadings in factor 6 in PT*

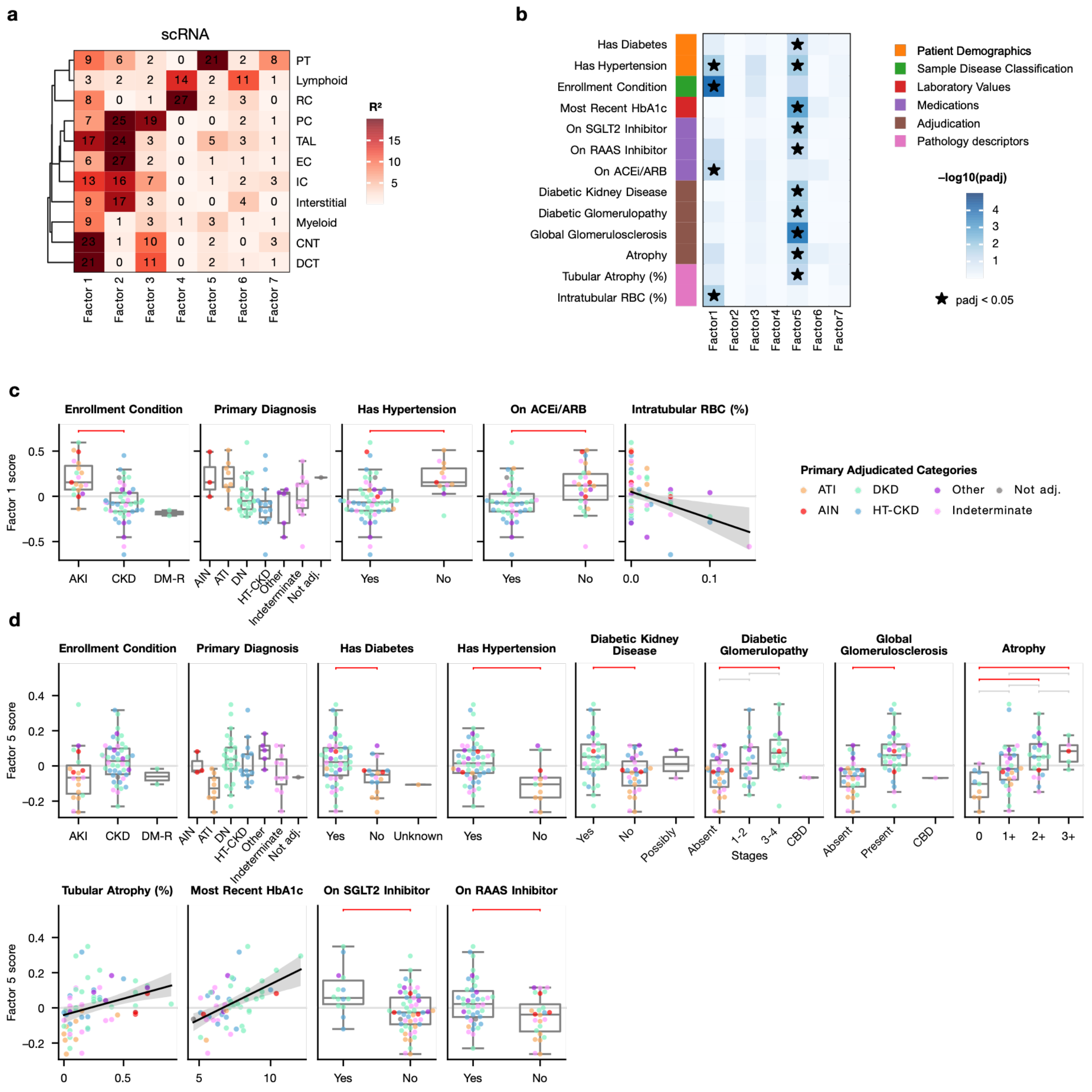

### Supplementary Figure 5:

*a. Gene expression variance explained by MCFA model in scRNA per cell type b. Statistical significance of the association between sample metadata and the MCFA factor scores in scRNA; * marks significant associations (adj. p. < 0.05). c./d. Metadata associations with Factor 1 and 5 respectively (acute, chronic) of scRNA. Horizontal brackets show Tukey's HSD tests in red if p < 0.05 and grey if not significant. CBD: Cannot be Determined*

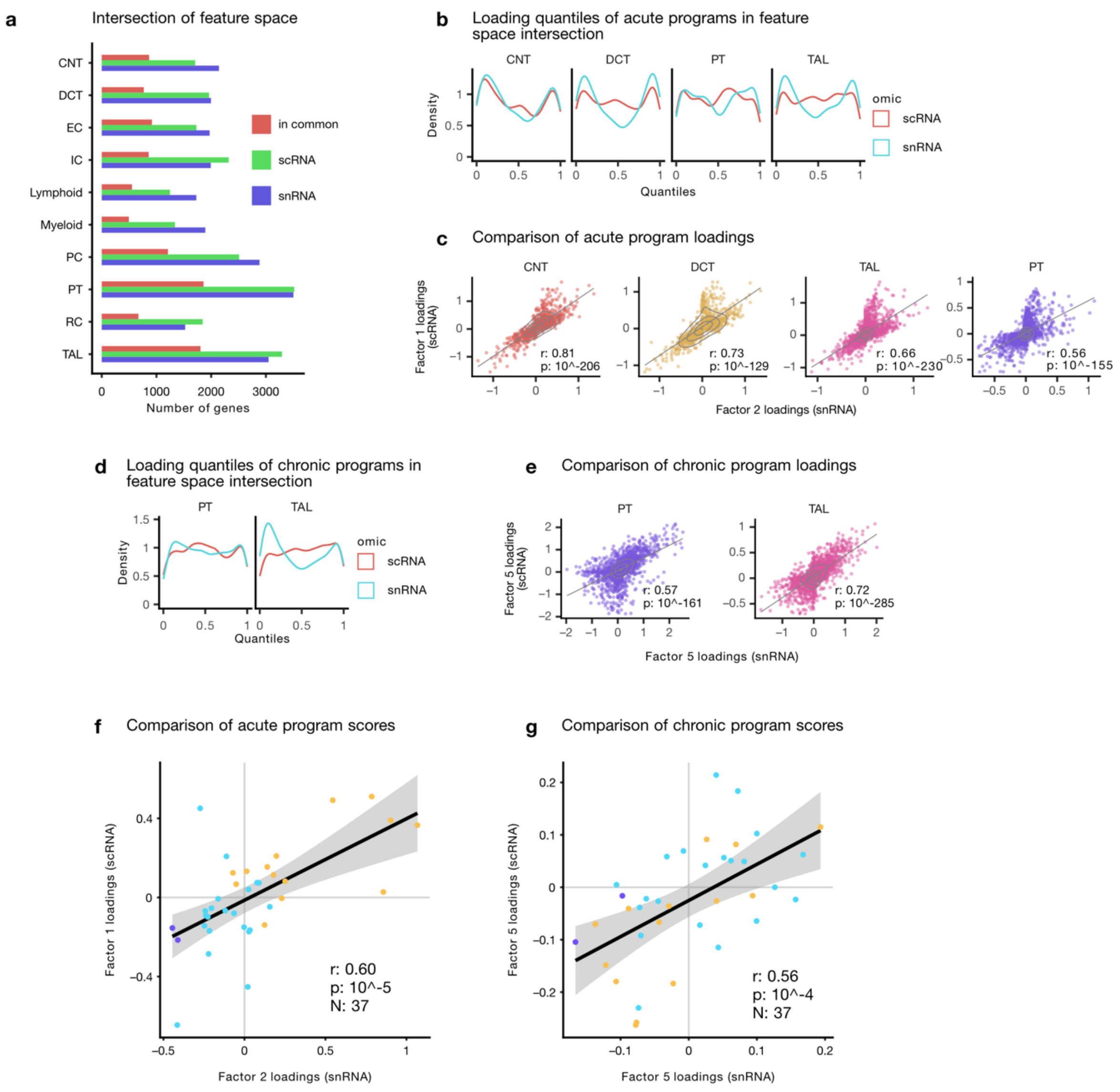

**Supplementary Figure 6:** Comparison of factors from snRNA and scRNA

*a. Gene overlap between snRNA and scRNA MCFA models, b. quantiles of gene loadings for genes included in the intersection of genes between snRNA factor 2 and scRNA factor 1 (acute programs) for best explained cell types, c. Pearson correlation of the gene loadings between snRNA factor 2 and scRNA factor 1 (acute programs), d. as in b. for factors 5 (chronic programs) from snRNA/scRNA, e. as in c. for factors 5 (chronic programs) from snRNA/scRNA, f. Pearson correlation of snRNA factor 2 and scRNA factor 1 scores (acute programs) for patients with both snRNA and scRNA data , g. as in f. for factors 5 (chronic programs) from snRNA/scRNA*

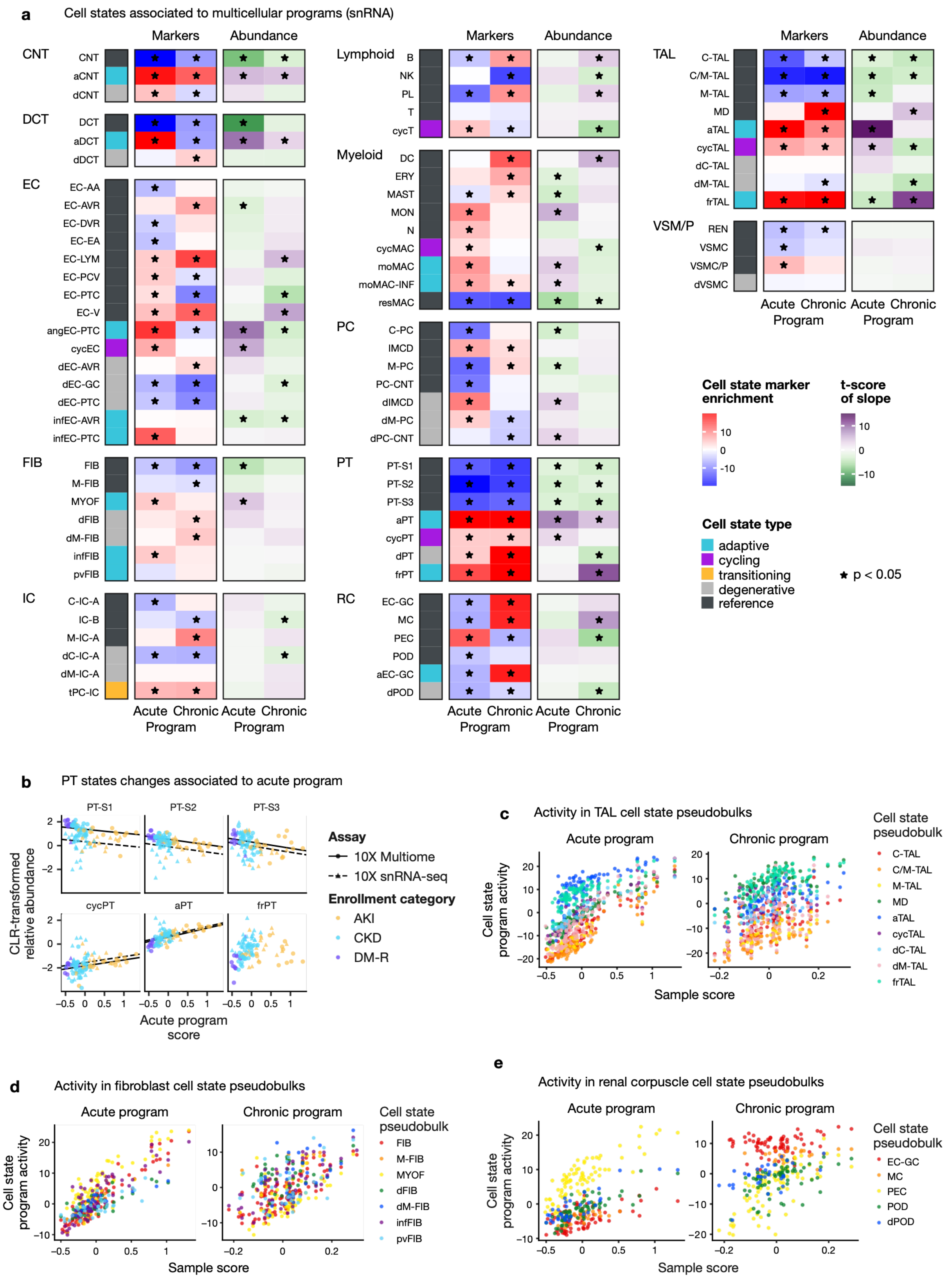

**Supplementary Figure 7:** Compositional and molecular shifts captured by acute and chronic programs in snRNA

*a. Cell state marker enrichment scores (red-blue heatmaps) in program loadings and t-scores of the slopes associating (e.g., in b) within-cell type relative cell state abundance to sample program scores (purple-green heatmaps) for major cell types in snRNA. * marks enrichments or slopes with padj < 0.05 b. CLR-transformed cell state proportion within PT relative to the acute sample program score. Regression lines show slope of the associated mixed-effect model, c-e acute and chronic program activity in PT, TAL and renal corpuscle cell state-level pseudobulks by sample program score in snRNA*

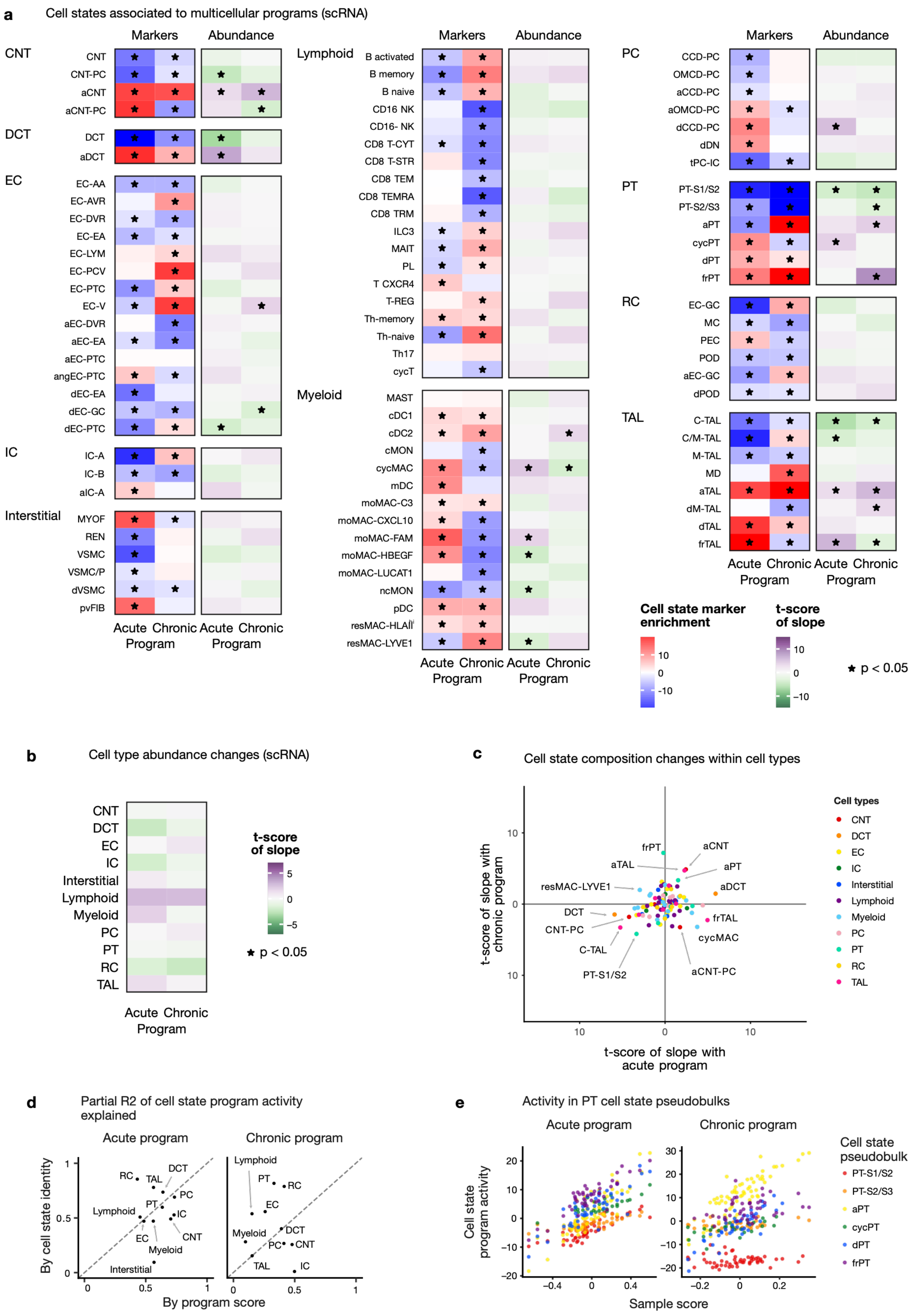

a
Cell states associated to multicellular programs (scRNA)
Markers
Abundance
CNT
CNT
CNT-PC
aCNT
aCNT-PC
DCT
DCT
aDCT
EC
EC-AA
EC-AVR
EC-DVR
EC-EA
EC-LYM
EC-PCV
EC-PTC
EC-V
aEC-DVR
aEC-EA
aEC-PTC
angEC-PTC
dEC-EA
dEC-GC
dEC-PTC
IC
IC-A
IC-B
aIC-A
Interstitial
MYOF
REN
VSMC
VSMC/P
dVSMC
pvFIB
Acute Chronic Acute Chronic
Program Program
Lymphoid
B activated
B memory
B naive
CD16 NK
CD16- NK
CD8 T-CYT
CD8 T-STR
CD8 TEM
CD8 TEMRA
CD8 TRM
ILC3
MAIT
PL
T CXCR4
T-REG
Th-memory
Th-naive
Th17
cycT
Myeloid
MAST
cDC1
cDC2
cMON
cycMAC
mDC
moMAC-C3
moMAC-CXCL10
moMAC-FAM
moMAC-HBEGF
moMAC-LUCAT1
ncMON
pDC
resMAC-HLAII
resMAC-LYVE1
Acute Chronic Acute Chronic
Program Program
PC
CCD-PC
OMCD-PC
aCCD-PC
aOMCD-PC
dCCD-PC
dDN
tPC-IC
PT
PT-S1/S2
PT-S2/S3
aPT
cycPT
dPT
frPT
RC
EC-GC
MC
PEC
POD
aEC-GC
dPOD
TAL
C-TAL
C/M-TAL
M-TAL
MD
aTAL
dM-TAL
dTAL
frTAL
Acute Chronic Acute Chronic
Program Program
Cell state marker enrichment
10
0
-10
t-score of slope
10
0
-10
★ p < 0.05
b
Cell type abundance changes (scRNA)
CNT
DCT
EC
IC
Interstitial
Lymphoid
Myeloid
PC
PT
RC
TAL
Acute Chronic
Program
t-score of slope
5
0
-5
★ p < 0.05
c
Cell state composition changes within cell types
t-score of slope with chronic program
t-score of slope with acute program
10
0
10
frPT
aCNT
aTAL
aPT
resMAC-LYVE1
aDCT
DCT
CNT-PC
frTAL
cycMAC
C-TAL
PT-S1/S2
aCNT-PC
Cell types
CNT
DCT
EC
IC
Interstitial
Lymphoid
Myeloid
PC
PT
RC
TAL
d
Partial R2 of cell state program activity explained
Acute program
Chronic program
By cell state identity
By program score
0
0.5
1
RC
TAL
DCT
PC
PT
IC
Lymphoid
EC
CNT
Myeloid
Interstitial
Lymphoid
PT
RC
EC
Myeloid
DCT
PC
CNT
TAL
IC
e
Activity in PT cell state pseudobulks
Acute program
Chronic program
Cell state program activity
Sample score
20
10
0
-10
-20
30
-0.4
0
0.4
-0.2
0.2
Cell state pseudobulk
PT-S1/S2
PT-S2/S3
aPT
cycPT
dPT
frPT

**Supplementary Figure 8:** Compositional and molecular shifts captured by acute and chronic programs in scRNA

*a. Cell state marker enrichment scores (red-blue heatmaps) in program loadings and t-scores of the slopes associating within-cell type relative cell state abundance to sample program scores (purple-green heatmaps) for major cell types in scRNA. * marks enrichments or slopes with padj < 0.05 b. Associations of acute and chronic program scores to overall shifts in relative cell type abundance in scRNA * mark significant associations (adj. p. < 0.05) c. t-scores of the slopes from mixed-effect model associating sample level acute (x-axis) and chronic (y-axis) program scores with relative cell state abundance within the respective cell types d. Partial R2 contribution of cell state identity and sample program score to the prediction of program activity in cell state pseudobulks in scRNA (e.g., show in e) e. Acute and chronic program activity in PT cell state-level pseudobulks by sample program score in scRNA*

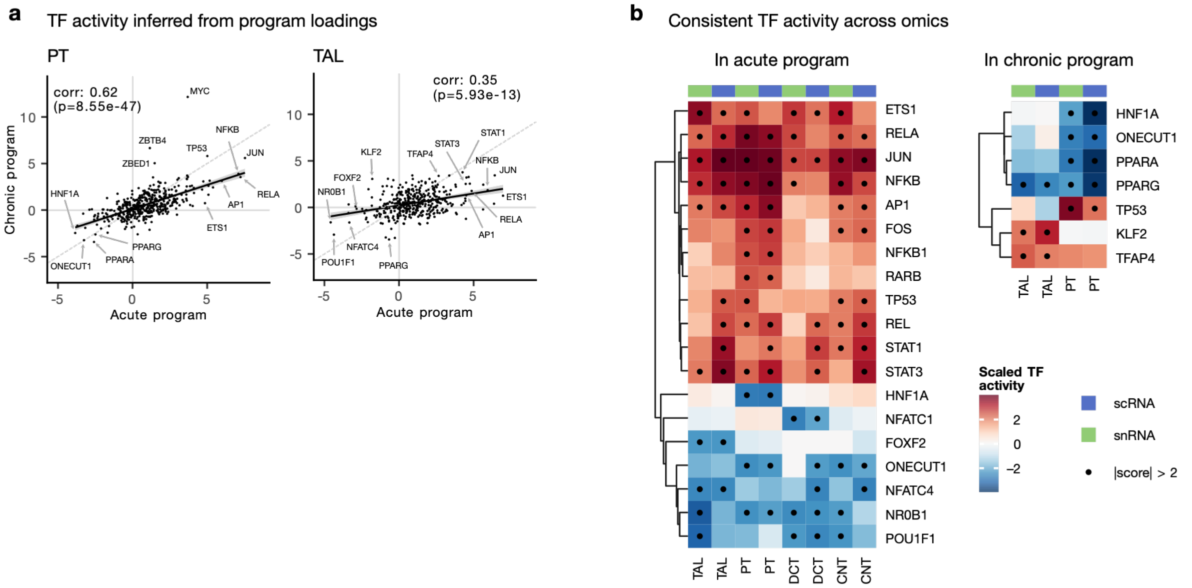

**Supplementary Figure 9:**

*a. Correlation between transcription factor activities in the loadings of the acute (left) and chronic (right) programs for PT and TAL b. Top most deregulated transcription factors (absolute z-standardized activity > 2) in common between snRNA and scRNA for the acute (left) and chronic (right) program*

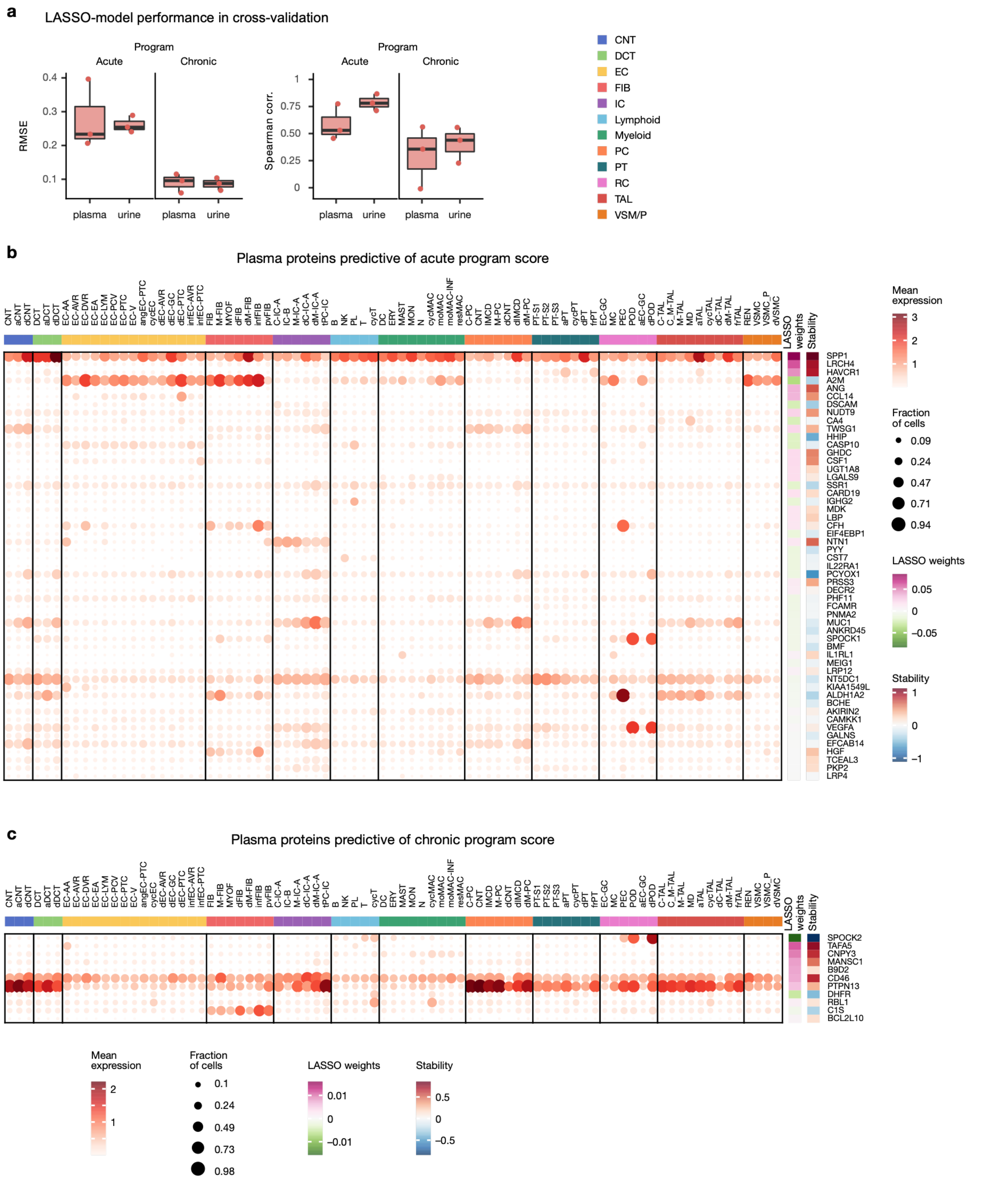

**Supplementary Figure 10:** Cell type and cell state specificity of expression of plasma proteins predictive of acute and chronic program scores

*a. LASSO-model performance in cross-validation in terms of root mean-square error (RMSE) and spearman correlation to predict acute/chronic program scores of samples from plasma or urine somascan proteins b./c. gene expression of plasma proteins that are cell type or cell state markers (dotplot), which were selected by the LASSO model to predict the acute and chronic program scores, respectively. The LASSO weights are in SD units for each protein. The stability is based on the mean/sd LASSO weight in bootstrap resampling times the inclusion rate (higher absolute value shows better numerical stability)*

**Supplementary Table S2:** Somascan data characteristics and models

| | **Urine** | **Plasma** |
|---|---|---|
| **Features** | | |
| Total measured | 7404 | 7596 |
| After QC | 5722 | 5431 |
| Acute model | 20 | 61 |
| Chronic model | 9 | 13 |
| **Samples** | | |
| Total measured | 137 | 137 |
| After QC | 137 | 121 |
| Acute and chronic models | 69 | 62 |
| Without snRNA | 68 | 59 |

*Total number of features in the somascan data, after filtering, and finally included in the acute and chronic LASSO models. Total number of samples with somascan data, that pass QC and which are finally used in the LASSO models (i.e. samples with both somascan and snRNA data). Samples without snRNA were used as a validation group within KPMP.*

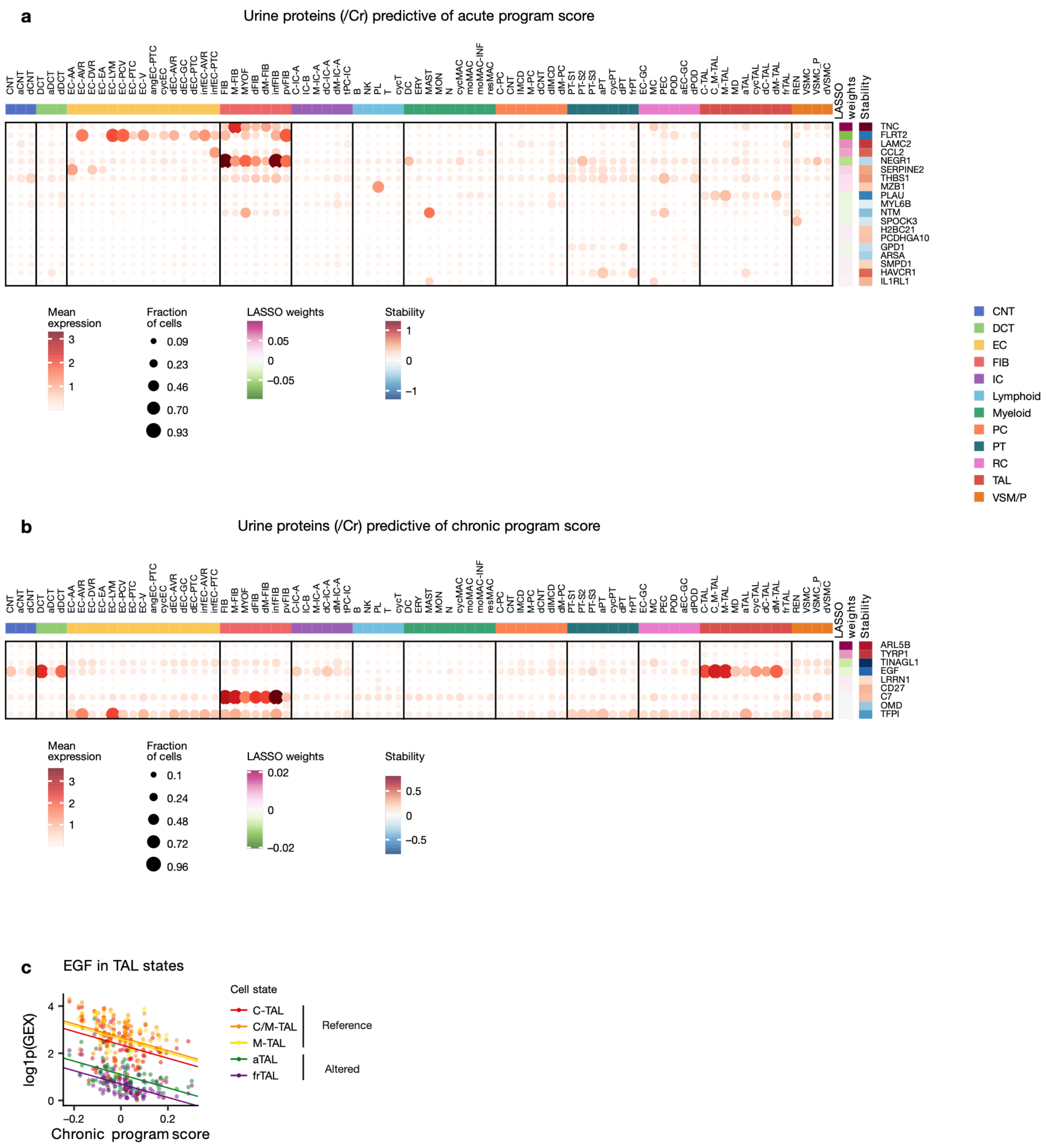

**Supplementary Figure 11:** Cell type and cell state specificity of expression of urine proteins predictive of acute and chronic program scores

*a./b. Gene expression of urinary proteins (normalized against urinary creatinine) that are cell type or cell state markers (dotplot), which were selected by the LASSO model to predict the acute and chronic program scores, respectively. The LASSO weights are in SD units for each protein. The stability metric is the mean/sd LASSO weight in bootstrap resampling times the inclusion rate (higher absolute value shows better numerical stability) c. EGF expression in TAL cell state pseudobulks relative to chronic program score. Slopes show the relation between gene expression and chronic program score with an intercept for each cell state, as determined by a mixed effect linear model.*

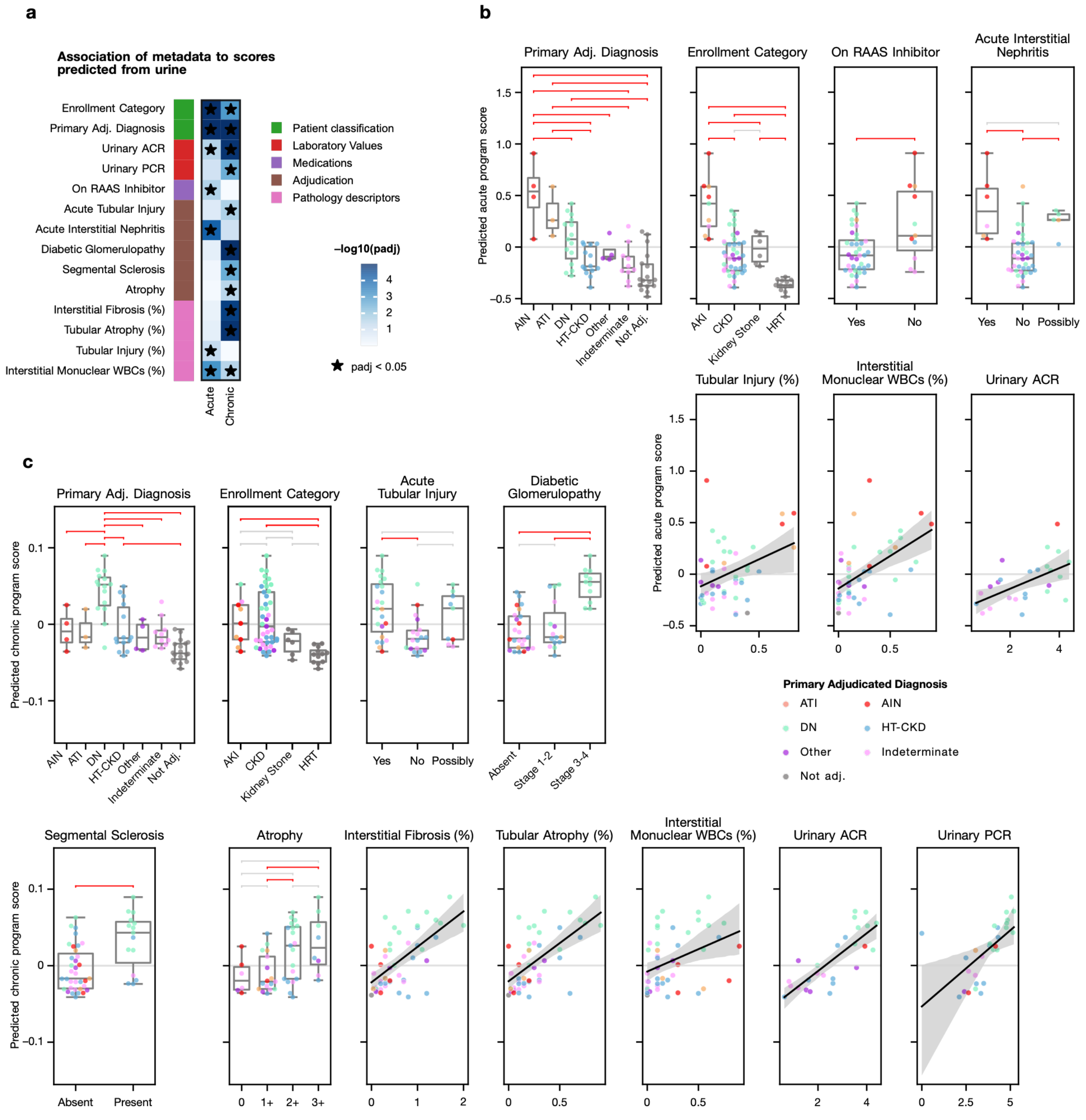

**Supplementary Figure 12:** Association of acute and chronic scores predicted from urinary protein abundances

*a. Significant associations between predicted scores and patient metadata. Adjusted p-values are shown based on analysis of variance or linear regression for categorical and continuous variables respectively. * marks significant* *associations (adj. p. < 0.05) b./c. Sample metadata associated with predicted acute and chronic scores. Horizontal brackets show Tukey's HSD tests in red if p < 0.05 and grey if not significant. CBD: cannot be determined.*

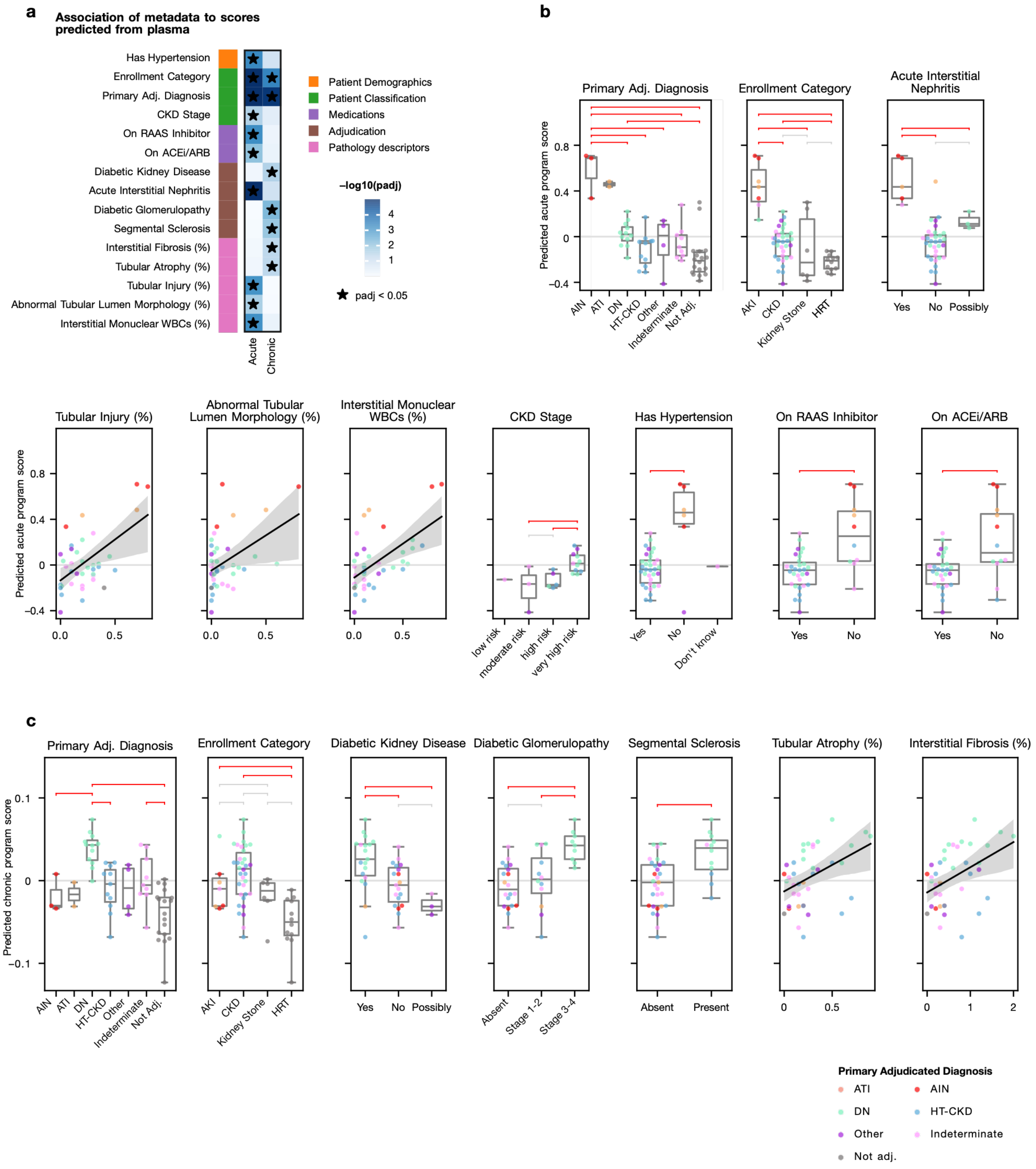

**Supplementary Figure 13:** Association of acute and chronic scores predicted from plasma protein abundances

*a. Significant associations between predicted scores and patient metadata. Adjusted p-values are shown based on analysis of variance or linear regression for categorical and continuous variables respectively. * marks significant* *associations (adj. p. < 0.05) b./c. Sample metadata associated with predicted acute and chronic scores. Horizontal brackets show Tukey's HSD tests in red if p < 0.05 and grey if not significant. CBD: cannot be determined.*

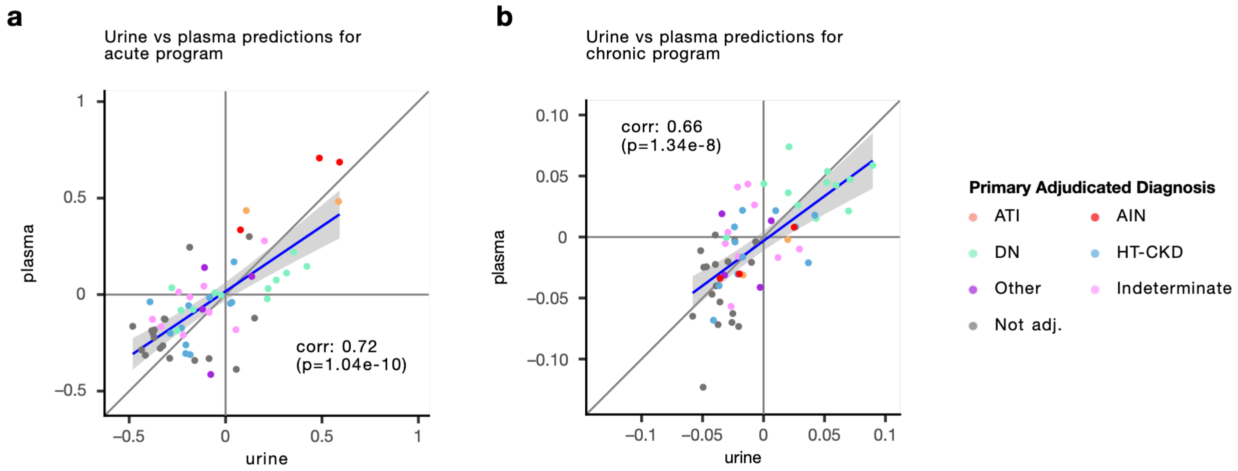

**Supplementary Figure 14:** Concordance between predicted acute and chronic scores

*a./b. Acute and chronic scores, respectively, predicted from urine and plasma proteins*

**Supplementary Table S3:** Baseline Characteristics of UK Biobank Participants by CKD Status.

| | CKD Status at Baseline | | | |
|---|---|---|---|---|
| **Characteristic** | **Overall** N = 45167[1] | **No CKD** N = 44250[1] | **CKD** N = 917[1] | **P-value**[2] |
| Age (years) | 56.8 (8.2) | 56.7 (8.2) | 62.9 (6.1) | <0.001 |
| Sex | | | | 0.8 |
| Female | 24,416 (54%) | 23,924 (54%) | 492 (54%) | |
| Male | 20,751 (46%) | 20,326 (46%) | 425 (46%) | |
| Body mass index (kg/m$^2$) | 27.5 (4.8) | 27.4 (4.8) | 29.4 (5.5) | <0.001 |
| Smoking status | | | | <0.001 |
| Never | 24,414 (54%) | 24,009 (55%) | 405 (44%) | |
| Current/Former | 20,545 (46%) | 20,038 (45%) | 507 (56%) | |
| Townsend deprivation index | -1.2 (3.2) | -1.2 (3.2) | -0.9 (3.3) | 0.007 |
| Serum creatinine (μmol/L) | 72.6 (17.2) | 71.4 (13.8) | 127.6 (47.2) | <0.001 |
| Serum cystatin C (mg/L) | 0.9 (0.2) | 0.9 (0.1) | 1.5 (0.5) | <0.001 |
| eGFR cr-based (mL/min/1.73m$^2$) | 94.2 (13.6) | 95.1 (12.1) | 50.1 (9.6) | <0.001 |
| eGFR cr-cys-based (mL/min/1.73m$^2$) | 94.3 (15.1) | 95.2 (13.8) | 51.4 (13.7) | <0.001 |
| RAS inhibitor use | 5,659 (13%) | 5,246 (12%) | 413 (45%) | <0.001 |
| Hypertension | 12,695 (28%) | 12,111 (27%) | 584 (64%) | <0.001 |
| Diabetes mellitus | 2,538 (6%) | 2,363 (5%) | 175 (19%) | <0.001 |
| Heart failure | 475 (1%) | 402 (1%) | 73 (8%) | <0.001 |
| Myocardial infarction | 1,184 (3%) | 1,080 (2%) | 104 (11%) | <0.001 |
| Ischemic heart disease | 2,511 (6%) | 2,331 (5%) | 180 (20%) | <0.001 |

| | | | | |
|---|---|---|---|---|
| Atherosclerosis | 72 (0%) | 53 (0%) | 19 (2%) | <0.001 |
| Peripheral vascular disease | 643 (1%) | 594 (1%) | 49 (5%) | <0.001 |
| Atrial fibrillation | 962 (2%) | 875 (2%) | 87 (9%) | <0.001 |
| Stroke | 162 (0%) | 146 (0%) | 16 (2%) | <0.001 |
| COPD | 1,073 (2%) | 1,010 (2%) | 63 (7%) | <0.001 |
| Asthma | 5,380 (12%) | 5,262 (12%) | 118 (13%) | 0.4 |
| Liver disease | 304 (1%) | 285 (1%) | 19 (2%) | <0.001 |
| Rheumatoid arthritis | 662 (1%) | 624 (1%) | 38 (4%) | <0.001 |
| Systemic lupus erythematosus | 371 (1%) | 349 (1%) | 22 (2%) | <0.001 |

[1]Mean (SD) or n (%)

[2]Welch Two Sample t-test; Pearson's Chi-squared test

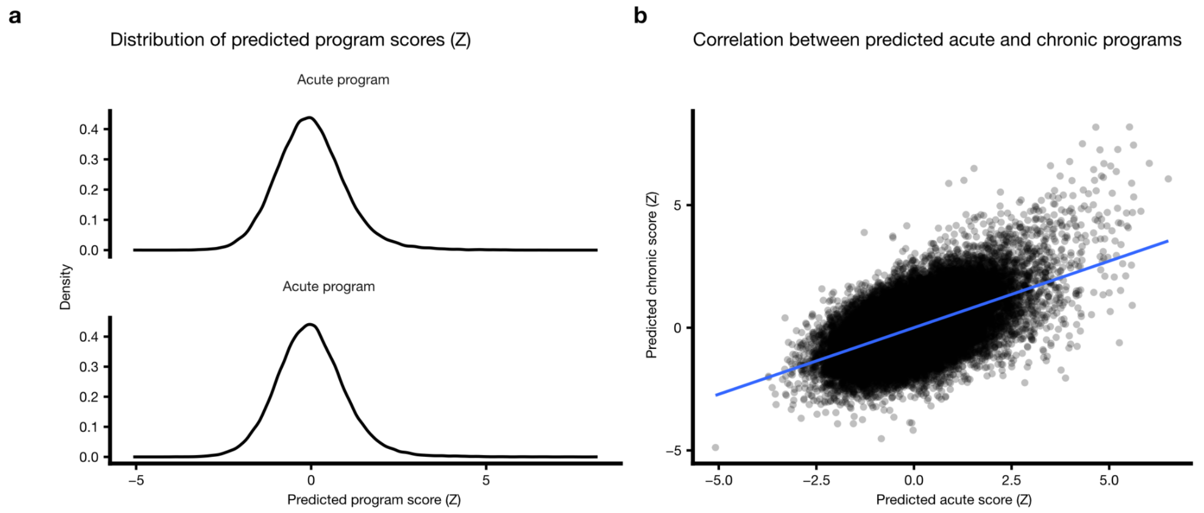

**Supplementary Figure 15:** Acute and chronic scores in the UK biobank

*a. Distribution of acute and chronic scores (z-transformed) predicted from plasma Olink data b. Relationship between predicted acute and chronic scores in the UK biobank*

**Supplementary Table S4:** Adjusted association between predicted program scores and eGFR.

| Program | Model | N | β (95% CI) | SE | P-value |
|---|---|---|---|---|---|
| Acute | Minimal adjustment | 45167 | -3.10 (-3.22 to -2.99) | 0.06 | <0.001 |
| Acute | Full adjustment | 44749 | -2.92 (-3.04 to -2.80) | 0.06 | <0.001 |
| Acute | No CKD stratum | 43843 | -1.64 (-1.76 to -1.53) | 0.06 | <0.001 |
| Acute | CKD stratum | 906 | -3.35 (-3.73 to -2.97) | 0.19 | <0.001 |
| Acute | Main effect | 44749 | -1.68 (-1.79 to -1.56) | 0.06 | <0.001 |
| Acute | Interaction term | 44749 | -1.61 (-2.09 to -1.12) | 0.25 | <0.001 |
| Chronic | Minimal adjustment | 45167 | -5.02 (-5.12 to -4.91) | 0.05 | <0.001 |
| Chronic | Full adjustment | 44749 | -4.88 (-4.98 to -4.77) | 0.05 | <0.001 |
| Chronic | No CKD stratum | 43843 | -3.58 (-3.68 to -3.47) | 0.05 | <0.001 |
| Chronic | CKD stratum | 906 | -3.60 (-3.92 to -3.27) | 0.17 | <0.001 |
| Chronic | Main effect | 44749 | -3.59 (-3.70 to -3.49) | 0.05 | <0.001 |
| Chronic | Interaction term | 44749 | -0.16 (-0.58 to 0.26) | 0.22 | 0.46 |

**Supplementary Table S5:** Hazard ratio of incident AKI based on acute or chronic plasma signature in UK biobank

| **Incident AKI (n = 2391/44250)** | | | | | | |
|---|---|---|---|---|---|---|
| Time Interval | Acute HR | Acute 95% CI (Lower) | Acute 95% CI (Upper) | Chronic HR | Chronic 95% CI (Lower) | Chronic 95% CI (Upper) |
| 0–3 months | 2.14 | 1.31 | 3.49 | 1.49 | 0.82 | 2.72 |
| 3–6 months | 2.13 | 1.42 | 3.2 | 2.15 | 1.39 | 3.31 |
| 6–12 months | 1.82 | 1.23 | 2.68 | 1.59 | 1.04 | 2.44 |
| >12 months | 1.54 | 1.48 | 1.61 | 1.35 | 1.29 | 1.41 |

**Supplementary Table S6:** Hazard ratio of incident CKD based on acute or chronic plasma signature in UK biobank

| **Incident CKD (n = 2315/44250)** | | | | | | |
|---|---|---|---|---|---|---|
| Time Interval | Acute HR | Acute 95% CI (Lower) | Acute 95% CI (Upper) | Chronic HR | Chronic 95% CI (Lower) | Chronic 95% CI (Upper) |
| 0–1 year | 1.5 | 1.29 | 1.75 | 1.45 | 1.24 | 1.7 |
| 1–3 years | 1.65 | 1.49 | 1.82 | 1.48 | 1.33 | 1.65 |
| 3–5 years | 1.59 | 1.44 | 1.77 | 1.52 | 1.36 | 1.69 |
| >5 years | 1.43 | 1.36 | 1.5 | 1.32 | 1.26 | 1.4 |

**Supplementary Table S7:** Hazard ratio of incident AKI or CKD based on combination of acute and chronic plasma signature in UK biobank

| **Phenotype Group** | **Incident AKI HR (95% CI)** | **Incident CKD HR (95% CI)** |
|---|---|---|
| Reference (low acute, low chronic) | 1.00 (reference) | 1.00 (reference) |
| High Acute Only | 1.48 (1.28–1.71) | 1.41 (1.19–1.66) |
| High Chronic Only | 1.07 (0.91–1.26) | 1.31 (1.10–1.55) |
| Double Hit (high acute, high chronic) | 2.13 (1.89–2.41) | 2.23 (1.94–2.56) |
